# Triglyceride-rich lipoproteins, low-density lipoproteins, and risk of abdominal aortic aneurysm

**DOI:** 10.64898/2026.02.22.26346555

**Authors:** Shuai Yuan, Elias Björnson, Gabrielle Shakt, Tia Dinatale, Julie A. Lynch, Ryan E. Temel, Hong S. Lu, Alan Daugherty, VA Million Veteran Program, Kyong-Mi Chang, Philip Tsao, Shaunak Adkar, Michael G. Levin, Scott M. Damrauer, Nicholas J. Leeper

**Affiliations:** Corporal Michael J. Crescenz VA Medical Center, Philadelphia, PA, USA; Department of Surgery, University of Pennsylvania Perelman School of Medicine, Philadelphia, PA, USA; Unit of Cardiovascular and Nutritional Epidemiology, Institute of Environmental Medicine, Karolinska Institutet, Stockholm, Sweden; Department of Medical Epidemiology and Biostatistics, Karolinska Institutet, Stockholm, Sweden; Department of Molecular and Clinical Medicine, Institute of Medicine, University of Gothenburg, Gothenburg, Sweden; VA Informatics and Computing Infrastructure (VINCI), VA Salt Lake City Heath Care System, Salt Lake City, Utah, USA; Division of Epidemiology, University of Utah School of Medicine, Salt Lake City, Utah, USA; Saha Cardiovascular Research Center, Saha Aortic Center, Department of Physiology, College of Medicine, University of Kentucky, Lexington, KY, USA; Department of Medicine, Perelman School of Medicine, University of Pennsylvania, Philadelphia, PA, USA; VA Palo Alto Healthcare System, Palo Alto, CA, USA; Department of Medicine, Division of Cardiovascular Medicine, Stanford University School of Medicine, Stanford, CA, USA; Stanford Cardiovascular Institute, Stanford University School of Medicine, Stanford, CA, USA; Department of Surgery, Division of Vascular Surgery, Stanford University School of Medicine, Stanford, CA, USA; Division of Cardiovascular Medicine, Department of Medicine, University of Pennsylvania, Perelman School of Medicine, Philadelphia, PA, USA; Department of Genetics, University of Pennsylvania Perelman School of Medicine, Philadelphia, PA, USA

**Keywords:** abdominal aortic aneurysm, aneurysmogenicity, lipids, triglyceride-rich lipoprotein cholesterol

## Abstract

**Background:** The comparative roles of triglyceride-rich lipoproteins (TRLs) and low-density lipoproteins (LDLs) in abdominal aortic aneurysm (AAA) pathogenesis are unclear.

**Objectives:** To evaluate the putative causal role of TRLs in AAA, quantify the relative effect on AAA risk (“aneurysmogenicity”) of TRL vs LDL particles, and prioritize lipid-lowering drug targets for AAA prevention and treatment.

**Methods:** We performed summary-level and individual-level Mendelian randomization (MR) analyses. Genetic variants were selected from 383,983 UK Biobank participants and ranked into 10 sets of variants where set 1 predominantly affected LDL cholesterol (LDL-C) and set 10 predominantly affected TRL cholesterol (TRL-C; and with mixed effects for intermediate variant sets). AAA outcome data were obtained from AAAgen (37,214 cases), FinnGen (4,439 cases), and the VA Million Veteran Program (MVP; 23,848 cases). Multivariable MR was used to assess the independent roles of LDL-C and TRL-C in AAA. For each set of variants, MR or logistic regression was used to estimate AAA odds ratios (ORs) per 10 mg/dL higher apolipoprotein B (apoB). Interaction analyses were conducted between a statin-like LDL-C-lowering variant set (set 3) and a TRL-C-lowering variant set (set 10). Drug-target MR was performed to evaluate lipid-lowering targets relevant to LDL-C- and TRL-C-lowering.

**Results:** Genetically predicted LDL-C and TRL-C concentrations were each associated independently with genetic liability for AAA after mutual adjustment, with 3.0 to 5.5 times stronger associations for TRL-C compared to LDL-C on a per-cholesterol basis. In AAAgen, the AAA OR per 10 mg/dL increased apoB concentrations were 1.10 (95% CI, 1.05–1.14) for variant set 1 (LDL-C-predominant) and 1.89 (95% CI, 1.69–2.11) for variant set 10 (TRL-C-predominant). Using the ratio of log(OR) per 10 mg/dL apoB for set 10 versus set 1 as a conservative estimate of relative aneurysmogenicity, TRLs were approximately 3.2 to 6.9 times more aneurysmogenic than LDLs across the three studies. No evidence of interaction was observed between LDLs and TRLs, indicating additive contribution to AAA risk. Drug-target MR supported strong protective associations for genetically proxied inhibition of TRL-pathway targets, particularly *APOC3* and *LPL*, with AAA risk.

**Conclusions:** TRLs are at least threefold more aneurysmogenic than LDLs on a per-particle basis. Therapeutic strategies targeting TRL-C —especially via *APOC3* and *LPL*—should be prioritized for AAA prevention and treatment.

## Introduction

Abdominal aortic aneurysm (AAA) is a common and potentially fatal vascular disease, and current management relies largely on surveillance and surgical repair. There is no approved drug therapy that prevents aneurysm development or slows progression (1), highlighting a need to identify actionable causal pathways for prevention and treatment.

Lipid dysregulation is a causal contributor to AAA pathogenesis (2–4). AAA frequently co-occurs with atherosclerotic cardiovascular disease, reflecting shared vascular risk biology that includes inflammatory activation, endothelial injury, and extracellular matrix remodeling (5). Consistent with this overlap, human genetic evidence supports a causal role of low-density lipoproteins (LDL) in AAA, and LDL-lowering pathways appear therapeutically actionable, with prior studies implicating PCSK9 as a potential target (4). In parallel, triglyceride-rich lipoproteins (TRLs)—including chylomicrons and very-low-density lipoproteins—and their cholesterol-enriched remnant particles are increasingly recognized as highly atherogenic. Genetic and clinical evidence across coronary artery disease and related phenotypes suggests that TRLs contribute risk through mechanisms that are at least partly independent of LDL (6–10). Because these pathways plausibly intersect with core AAA biology, we hypothesize that TRL metabolism is causally involved in AAA and may represent complementary targets beyond LDL lowering, particularly along the lipoprotein lipase (LPL)–mediated lipolysis and remnant clearance axis, including APOC3-mediated regulation (11).

Recent AAA-specific data extend this framework. In an integrative Mendelian randomization (MR) analyses, elevated triglyceride-rich lipoproteins and triglyceride metabolism–related proteins/metabolites showed causal associations with AAA risk (12). Animal-based studies further support a causal, dose-dependent role of triglyceride-rich lipoproteins in AAA, as hypertriglyceridemic models (e.g., Lpl-or Apoa5-deficient mice) show more severe disease and higher rupture risk, while pharmacologic triglyceride lowering via Angptl3 antisense attenuates AAA progression (12). However, human data directly interrogating TRL-specific measures (e.g., TRL remnants or TRL particle metrics) in AAA remain limited. Clarifying whether TRLs are causally related to AAA could therefore sharpen mechanistic understanding and help prioritize TRL-directed therapies as potential pharmacologic strategies for AAA.

MR leverages germline genetic variants as instrumental variables to strengthen causal inference in the presence of confounding and reverse causation, under assumptions regarding instrument relevance, independence, and exclusion restriction; these assumptions can be partially interrogated using established sensitivity analyses (13). Here, we conducted comprehensive MR analyses aiming to: (1) assess the putative causal role of TRLs in AAA risk; (2) compare the relative “aneurysmogenicity” of TRLs to LDLs in AAA, both on a per-cholesterol and per-apoB basis; and (3) evaluate whether genetic proxies of LDL- and TRL-lowering drug targets support application for AAA therapy.

## Methods

### Study design

This study combined summary-level and individual-level data to evaluate the role of TRL in AAA and to identify potential therapeutic targets for AAA therapy prevention. **Figure 1** shows the study design overview. First, we performed multivariable MR analysis to quantify the effect of LDL and TRL in AAA (i.e. on a per-cholesterol basis). We then performed variant-set based analyses to estimate the relative per-particle (per-apoB) effects of LDL and TRL in AAA. Lastly, we assessed candidate drug targets that predominantly reduced LDL-C or TRL-C concentrations via key lipid-metabolism genes and estimating their associations with AAA risk.

**Figure 1.**
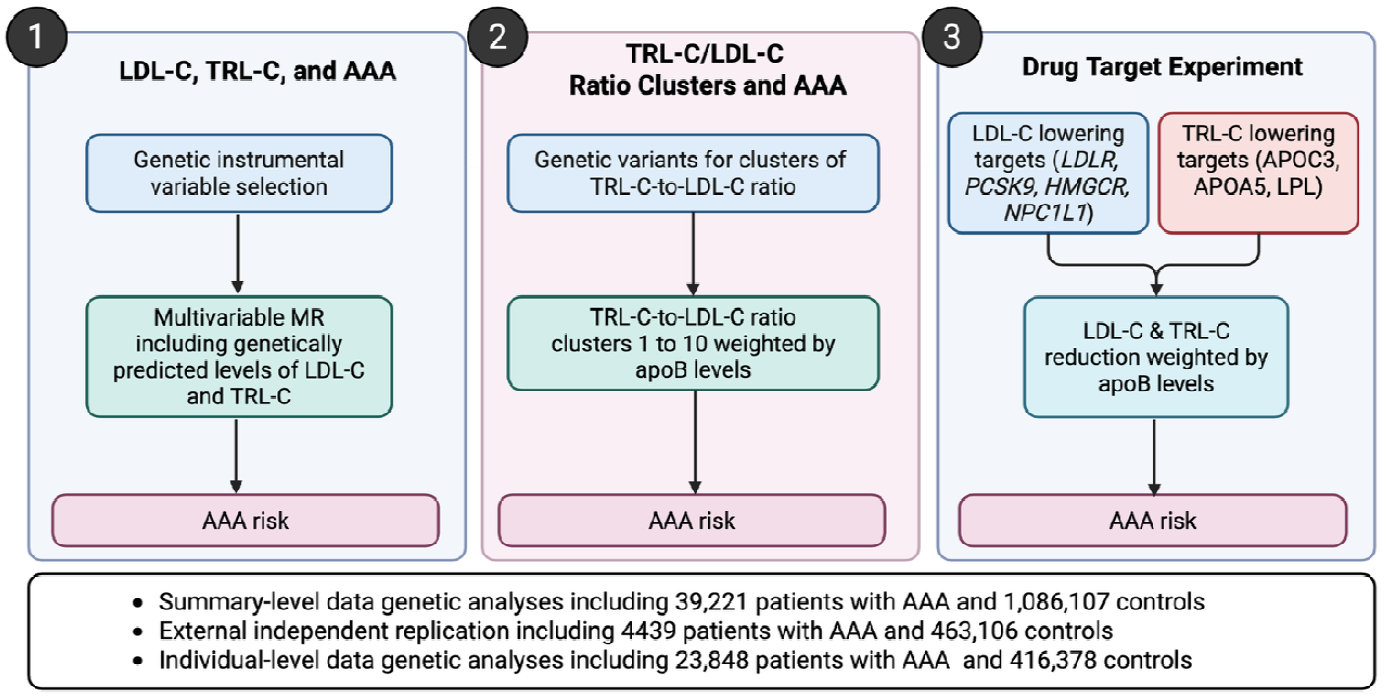
Study design overview. AAA, abdominal aortic aneurysm; apoB, apolipoprotein B; LDL-C, low-density lipoprotein cholesterol; TRL-C, triglyceride-rich lipoprotein cholesterol.

### Lipid measurements

Lipid measurements were derived from the UK Biobank resource, including over 502,000 UK residents of mainly European ancestry (14). LDL-C was directly measured using a Beckman Coulter assay (Brea, CA). Plasma non–high-density lipoprotein cholesterol (non–HDL-C) concentrations were calculated as total cholesterol minus HDL-C concentrations. TRL was calculated as non–HDL-C minus directly measured LDL-C. Because TRL concentrations were derived from measured components, it was considered as an indirectly measured concentration.

### Genetic instrumental variable (IV) selection

Variant selection has been described previously (6). Briefly: to obtain genetic instruments, we conducted a genome-wide association study (GWAS) for plasma triglycerides concentrations (n=383,983), TRL-C (n=350,797), and LDL-C (n=383,566) among UK Biobank participants with both genetic and lipid data who were not using lipid-lowering therapy at the time of lipid measurement. GWAS models were adjusted for age, sex, and genetic principal components 1– 5. Variants reaching genome-wide significance (*P* < 5 × 10^−^□) were pruned for linkage disequilibrium (*r*^2^ < 0.1 within a 20-Mb window) and filtered by minor allele frequency (MAF > 0.01). When variants were in linkage disequilibrium, we retained the variant with the largest combined effect magnitude across LDL-C and TRL-C, defined as 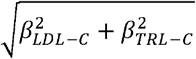. The resulting variant set was further filtered to remove variants associated with lipoprotein(a) after Bonferroni correction (*P* < 0.05), excluding 28 variants. This procedure yielded a final instrument set of 1,357 variants (**Table S1**) (6).

#### IVs for variant sets

Selected variants were ranked by the ratio of their effect sizes (β coefficients) for TRL-C relative to LDL-C. The ranked variants were then partitioned into 10 sets of variants, with each set containing an equal number of variants. Genetically predicted TRL-C-to-LDL-C ratio ranged from <0.2 in variant set 1 to >3 in variant set 10 meaning that genetic instruments in set 1 predominantly affected LDL-C with minor effects on TRL-C and genetic instruments in set 10 primarily affected TRL-C with minor effects on LDL-C (and with mixed effects for intermediate variant sets). Based on each variant set’s effects on LDL-C and TRL-C levels, set 3 was considered a proxy for statin-like effects (statins reduce LDL-C and TRL-C in a similar manner as set 3, and several *PCSK9* and *LDLR* variants are found in this set), whereas set 10 was considered a proxy for TRL-lowering treatment (near exclusive effect on TRL-C without major LDL-effect, and *LPL* and *APOC3* variants are contained in this set).

#### IVs for drug targets

Among the 1,357 variants, we selected variants in the cis regions of key genes encoding lipid metabolism–related proteins (e.g., receptors, enzymes, inhibitors, activators) as genetic instruments for lipid-lowering drug targets. These included *PCSK9, HMGCR, LDLR*, and *NPC1L1*, which primarily act to affect LDL-C, and *APOC3, APOA5*, and *LPL*, which primarily act to affect TRL-C.

### Summary-level genetic analyses

Two-sample MR analyses using summary-level data were conducted with AAA GWAS results from the AAAgen consortium including 14 studies (4). UK Biobank contributed a small proportion of cases (∼3.3%); given the strong instruments, any bias from sample overlap is unlikely to be material (15). The FinnGen R12 AAA GWAS was used as the independent replication data source (16).

### Individual-level genetic analyses

We conducted individual-level polygenic risk score (PRS) analyses in the VA Million Veteran Program (MVP), a large national cohort of U.S. Veterans established by the Department of Veterans Affairs to investigate genetic and environmental determinants of human disease (17). PRSs for each of the 10 variant sets and the lipid-lowering drug target variants were calculated using PLINK2 (18) for all individuals using a weighted sum (weighted by beta coefficients for apoB) and scaled so that 1 polygenic score unit represented a concentration of 10 mg/dL apoB. AAA cases were defined as participants with at least two occurrences of relevant diagnostic codes in the electronic health record (ICD-9: 441.3 or 441.4; ICD-10: I71.3 or I71.4) (14). Comparator participants without AAA had no such codes and were additionally excluded if they carried any related vascular disease codes (ICD-9: 440–448; ICD-10: I71–I75, I77–I79, K55). Genotyping was performed using the custom ThermoFisher Axiom MVP 1.0 array, and imputation was conducted using the TOPMed reference panel (19). The present analysis was restricted to participants genetically similar to the 1000 Genomes European (EUR) superpopulation (20), as determined using PLINK2.

### Statistical analyses

Multivariable MR analyses were used to compare effects of LDL-C and TRL-C on AAA with mutual adjustment. In MR of TRL-C/LDL-C ratio variant set, the inverse-variance weighted (IVW) method under a multiplicative random-effects model was used as the primary analysis and supplemented by the weighted median (21), MR-Egger (22), and MR-PRESSO (23) to evaluate robustness and potential horizontal pleiotropy. For individual-level analyses, PRSs were tested for association with AAA by logistic regression with adjustment for age, sex, and top 10 genetic principal components. Multiplicative and additive interactions between PRSs were assessed. All statistical analyses were conducted in R (version 4.2.2).

## Results

The summary-level analyses included 37,214 participants with AAA and more than one million participants without an AAA diagnosis from the AAAgen consortium, and 4,439 participants with an AAA and 463,106 without a clinically diagnosed AAA from FinnGen release R12. The individual-level analysis was conducted among 23,848 participants with AAA and 416,378 without a clinically diagnosed AAA from the VA MVP cohort.

Multivariable MR analysis using all 1,357 variants demonstrated that genetically predicted LDL-C and TRL-C concentrations were each independently associated with AAA after mutual adjustment. However, cholesterol carried in TRL particles showed a stronger association with genetically predicted AAA risk than cholesterol carried in LDL particles (**Figure 2A**). On a per-cholesterol basis TRL-C were found to be approximately 5.47 (95% CI, 3.18-7.76) times more aneurysmogenic compared to LDL-C in AAAgen, 3.56 (95% CI, 1.57-5.54) in FinnGen, and 2.98 (95% CI, 2.82-3.16) in MVP.

**Figure 2.**
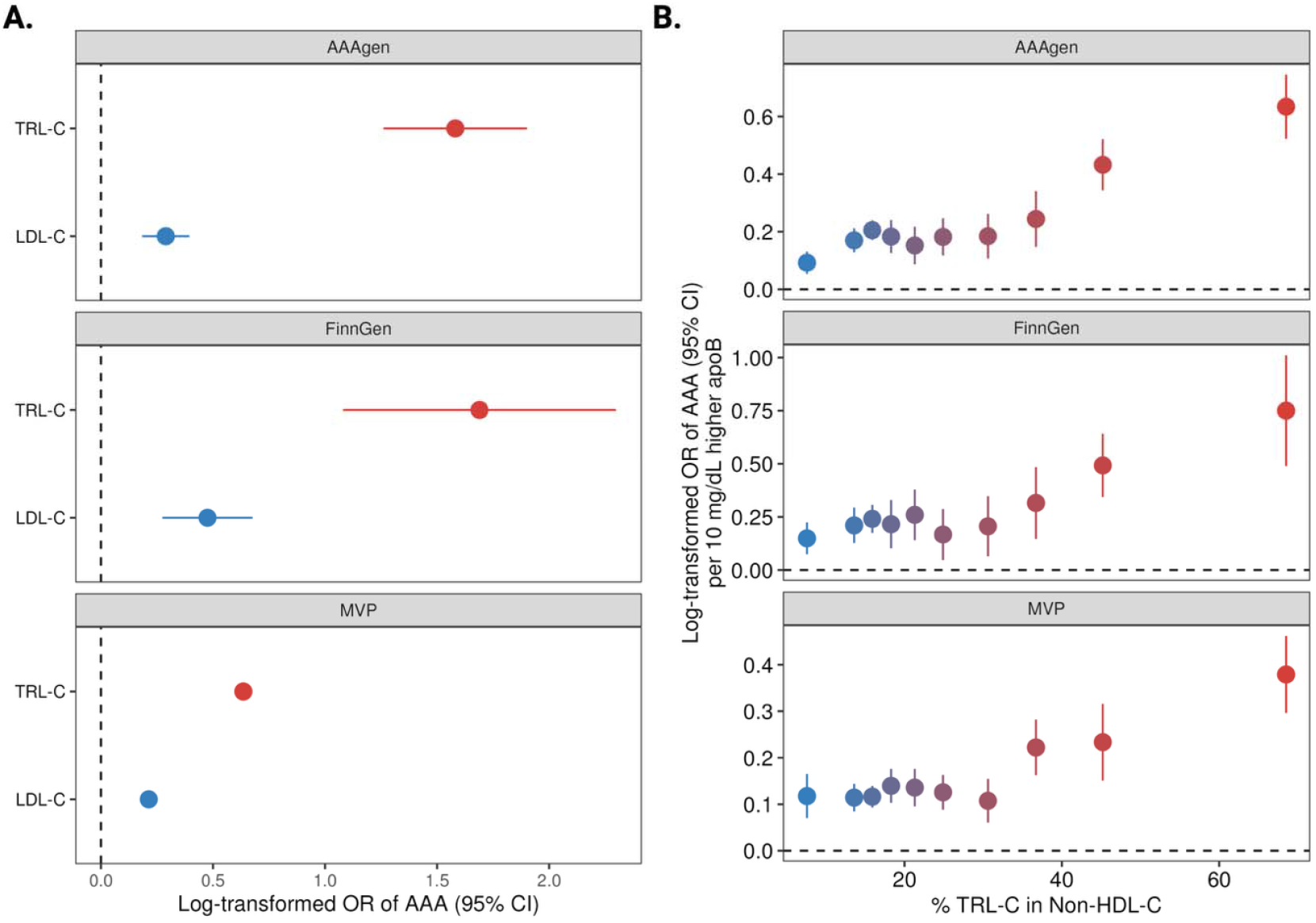
Aneurysmogenicity of triglyceride-rich lipoproteins cholesterol (TRL-C) vs low-density lipoprotein cholesterol (LDL-C). AAA, abdominal aortic aneurysm; MVP, Million Veteran Program. Panel A. multivariable Mendelian randomization of TRL-C and LDL-C in relation to AAA in AAAgen and FinnGen studies. Panel B. Mendelian randomization analysis of genetically proxied variant sets of TRL-C–to–LDL-C ratios in relation to AAA risk. The associations were scaled to per 10 mg/dL increase in apolipoprotein B. The x axis shows the average variant %TRL-C in non-high-density lipoprotein cholesterol among each variant set.

Per 10 mg/dL increased apoB, the more TRL-raising-effect a variant set had, the higher the AAA risk was observed (**Figure 2B**). In the AAAgen consortium, the odds ratio (OR) was 1.10 (95% CI, 1.05–1.14) for the LDL-specific variant set (set 1) and 1.89 (95% CI, 1.69–2.11) for the TRL-specific variant set (set 10). These findings were consistent in the summary-level FinnGen replication, in the individual-level VA MVP cohort (**Figure 2B**), and in MR sensitivity analyses (**Table S2**). Using a conservative metric of relative aneurysmogenicity—defined as the ratio of the AAA log-odds ratio per 10 mg/dL apoB for variant set 10 to the corresponding value for set 1—the estimated relative aneurysmogenicity was 3.23 (95% CI, 1.74–4.71) in MVP, 5.03 (95% CI, 1.95–8.11) in FinnGen, and 6.87 (95% CI, 3.73–10.0) in AAAgen.

No multiplicative interaction was observed between the statin-like PRS (variant set 3) and the TRL-related PRS (variant set 10; *P* for interaction = 0.298). The odds ratio for AAA per 10 mg/dL higher apoB associated with the TRL-related PRS was similar at the 75th percentile compared with the mean of the LDL-related PRS (OR 1.42 vs 1.46). On the additive scale, the interaction contrast was close to zero (IC = −0.002; 95% bootstrap CI −0.010 to 0.005), and the absolute risk difference per 10 mg/dL higher apoB was similar at the 25th and 75th percentiles of the LDL-related PRS (RD 0.023 vs 0.021).

Genetically proxied lower levels of apoB via LDL gene targets (*LDLR* and *PCSK9*) and TRL gene targets (*APOC3, LPL*, and *APOA5)* were associated with a lower risk of AAA in AAAgen (**Figure 3**). Associations for *LDLR, PCSK9, APOC3*, and *LPL* were consistent in both FinnGen and MVP. Of note, genetically proxied lower levels of apoB through TRL drug targets showed a stronger protective association with AAA (**Figure 3**). Genetically proxied inhibition of the LDL-target set and the TRL-target set was associated with AAA odds ratios of 0.76 (95% CI, 0.73–0.79) and 0.53 (95% CI, 0.47–0.59) per 10 mg/dL lower apoB-equivalent, respectively, in AAAgen. Per unit cholesterol lowering, TRL drug-target variants showed a 2.82-fold (95% CI, 2.17-3.48) larger AAA-risk reduction per 1 mmol/L TRL-C reduction compared to 1 mmol/L LDL-C reduction using LDL drug-target variants.

**Figure 3.**
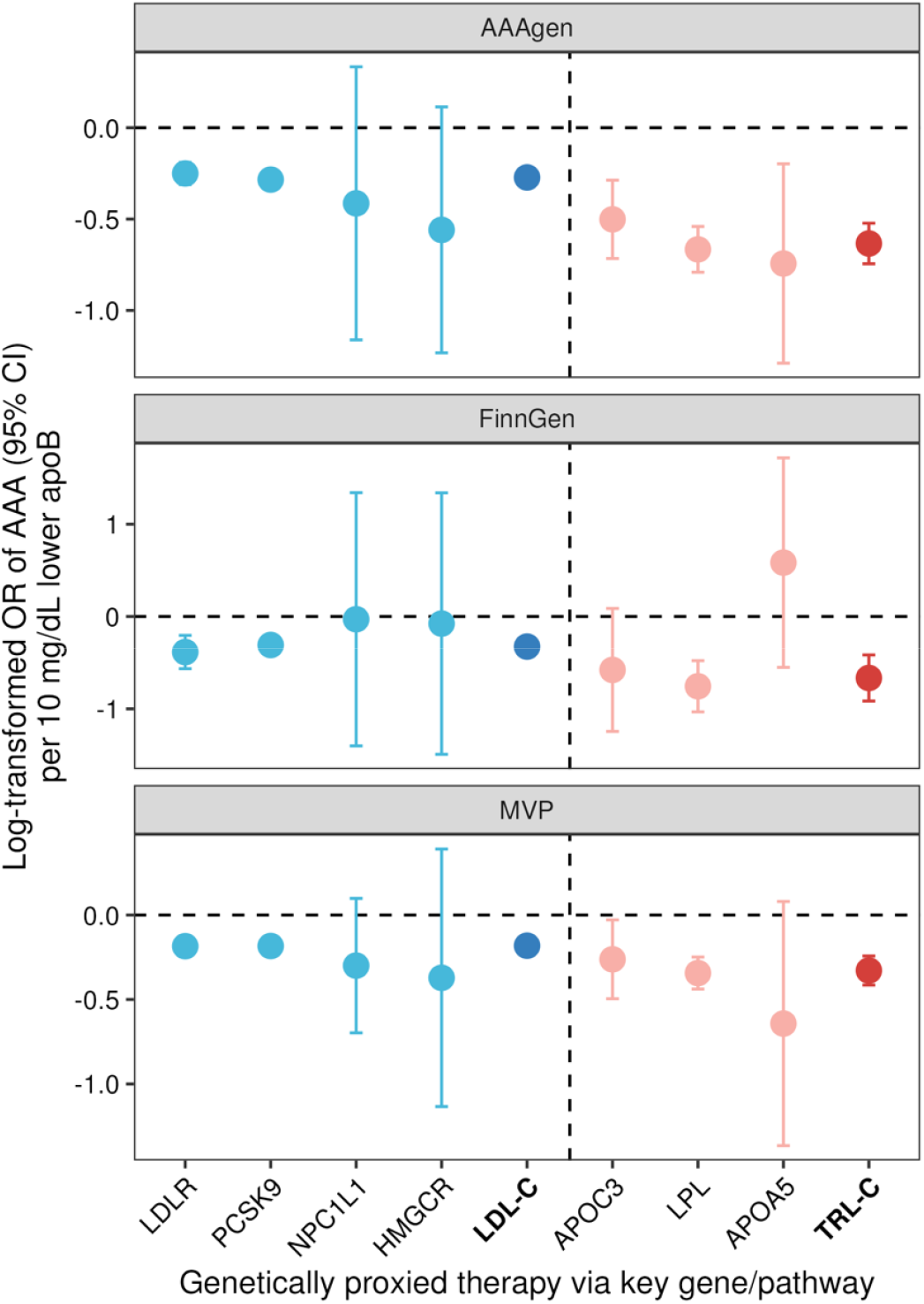
Genetic inhibition of triglyceride-rich lipoproteins cholesterol (TRL-C) and low-density lipoprotein cholesterol (LDL-C)-related targets in relation to abdominal aortic aneurysm risk. AAA, abdominal aortic aneurysm. MVP, Million Veteran Program. The associations were scaled to per 10 mg/dL decrease in apolipoprotein B. TRL-C included APOC3, LPL, and APOA5 targets. LDL-C included LDLR, PCSK9, NPC1L1, and HMGCR targets.

## Discussion

Given the lack of approved pharmacologic therapies for AAA, we conducted MR-based investigations using data from three large-scale cohorts to evaluate the causal role of TRL-C in AAA and to prioritize therapeutic targets. We determined that both LDLs and TRLs contribute causally to AAA risk; however, per unit apoB, TRLs were at least ∼3-fold more aneurysmogenic than LDLs. Interaction analyses provided no meaningful evidence of effect modification, supporting predominately independent contributions of the LDL- and TRL- pathways to AAA. These findings suggest that drug targets that lower apoB-containing lipoproteins—particularly those acting on the TRL metabolism axis, such as *APOC3* and *LPL*—should be prioritized for AAA prevention and treatment.

Prior evidence has most consistently implicated LDL-related pathways in AAA (4). Observationally, AAA co-segregates with atherosclerotic disease and is enriched among individuals with dyslipidemia. Several studies have reported associations between statin use and slower AAA growth and improved outcomes, although confounding by indication and healthy-user bias remain concerns (24–26). More compellingly, human genetics has supported LDL-pathway causality: the large AAAgen GWAS meta-analysis identified *PCSK9* as a therapeutic target, strengthening the argument that LDL-related mechanisms contribute to AAA risk (4). Taken together, these data provide a strong rationale to expect that LDL-lowering could be beneficial for AAA, even though definitive randomized evidence using AAA-specific endpoints remains limited.

In contrast, the AAA literature has historically provided less direct evidence for TRLs than for LDL-related pathways. While triglycerides and remnant cholesterol have been linked to vascular risk across multiple settings (7–10), AAA-focused observational findings have often been heterogeneous and difficult to interpret because TRL-related biomarkers correlate strongly with obesity, insulin resistance, and smoking. Recent reviews and emerging genetic work have begun to suggest a role for TRLs in aortic disease (27). This landscape has shifted with the recent study by Liu et al, which integrated genetic, proteomic, and metabolomic MR analyses to implicate elevated TRLs and triglyceride metabolism–related proteins/metabolites in AAA risk (12). Consistent with a causal, dose-dependent effect, Liu et al. further demonstrated markedly worsened AAA phenotypes in hypertriglyceridemic mouse models—including rupture in Lpl-deficient mice and accelerated aneurysm development in Apoa5-deficient mice—and showed that lowering triglycerides via an Angptl3-targeting antisense oligonucleotide attenuated AAA progression in hypertriglyceridemic settings (12). Even with these advances, the field has lacked a clear, head-to-head quantification of TRL versus LDL contributions to AAA risk in humans and a systematic prioritization of actionable TRL targets for AAA. Our results address this gap by demonstrating independent associations for LDL-C and TRL-C, substantially greater aneurysmogenicity for TRLs on a per-particle basis, and particularly strong drug-target MR support for the *LPL* axis (notably APOC3 and LPL).

Why TRLs appear more aneurysmogenic than LDLs is uncertain, but several non-mutually exclusive mechanisms are plausible. TRLs and their remnants may disproportionately provoke vascular inflammation and endothelial activation relative to LDL particles, including upregulation of adhesion molecules and cytokine responses triggered by TRLs and their lipolysis products (28). Given that AAA is characterized by chronic inflammation, extracellular matrix degradation, and smooth muscle cell loss, a TRL-linked inflammatory milieu could plausibly amplify aneurysmal remodeling beyond what would be expected from cholesterol transport alone. These hypotheses are consistent with the pronounced per-ApoB effects observed here but require direct testing in mechanistic and translational studies focused on aneurysm biology.

No cardiovascular outcomes trial (CVOT) has yet reported effects of APOC3 inhibition on AAA; however, our genetic findings allow a rough, genetically informed estimation of the magnitude of benefit that might reasonably be expected. First, we noted that the absolute risk estimates for both LDLs and TRLs in relation to AAA observed in this investigation were remarkably similar to those observed with coronary heart disease as outcome using the same genetic instruments (6). This concordance suggests that, as a first approximation, a given absolute reduction in atherogenic lipoprotein burden may translate into a comparable risk reduction for AAA as has been robustly demonstrated for coronary heart disease. Empirically, randomized trials and meta-analyses indicate that a 1 mmol/L reduction in LDL-C concentrations yields approximately a 22–24% relative reduction in coronary heart disease risk (29). Our genetic estimates imply a similar order of magnitude for AAA. Second, based on our drug-target MR results, cholesterol carried in TRL particles was at least two-fold more strongly associated with AAA risk than cholesterol carried in LDL particles. Conservatively, this would suggest that a reduction of ∼0.5 mmol/L (or around 20 mg/dL) in TRL-C concentrations would confer a risk reduction similar to that achieved by lowering LDL-C by ∼1.0 mmol/L (39 mg/dL). Under this assumption, a reduction in TRL-C concentrations of ∼0.5 mmol/L—which is within the range observed with APOC3 inhibition—would be expected to translate into an approximately 20–25% relative reduction in AAA risk, assuming causal effects and proportionality analogous to those observed for LDL-C and coronary heart disease.

Several limitations should be considered. MR estimates reflect lifelong exposure differences and may overstate short-term benefits achievable with late-life pharmacologic intervention. TRL-C concentrations in UK Biobank were derived as non–HDL-C minus directly measured LDL-C, which may introduce measurement error and attenuate instrument strength. As with all MR analyses, residual pleiotropy cannot be fully excluded, particularly for targets with potential lipid-independent effects such as *PCSK9* or *APOC3*. Finally, AAA phenotyping across cohorts and healthcare systems may vary, and these findings may not generalize to all ancestries or to outcomes such as aneurysm growth rate versus incident AAA.

In conclusion, our genetic data support a causal role for both LDLs and TRLs in AAA, indicate substantially greater aneurysmogenicity of TRL particles compared to LDL particles, and prioritize APOC3-inhibition as a promising AAA therapeutic strategy. Given the absence of approved pharmacologic therapies for AAA, incorporation of AAA-related endpoints into ongoing and future TRL- and/or LDL-lowering trials may be justified.

## Acknowledgments

This research is based on data from the Million Veteran Program, Office of Research and Development, and Veterans Health Administration, with support from MVP000, VA Merit Award #I01-BX003362 (Chang/Tsao) and the Department of Veterans Affairs (VA) Informatics and Computing Infrastructure (VINCI), including data analytics conducted by its Precision Medicine research team, which is funded under the research priority to Put VA Data to Work for Veterans (VA ORD 24-D4V-02). This research used resources of the Knowledge Discovery Infrastructure (KDI) at the Oak Ridge National Laboratory, which is supported by the Office of Science of the U.S. Department of Energy under Contract No. DE-AC05-00OR22725. This publication does not represent the views of the Department of Veterans Affairs or the United States Government. We gratefully acknowledge the Veterans who participated in the VA’s Million Veteran Program. This work was also supported by the National Heart Lung and Blood Institute grants R01HL166991 and R01HL171587 (to S.M.D) and R35HL176060 (to N.J.L). S.Y. was supported by an Award from the American Heart Association (Award ID: 24POST1189614) and the VIVA Foundation. The content in this manuscript is solely the responsibility of the authors and does not necessarily represent the official views of the National Institutes of Health. E.B. received support from Swedish Heart-Lung Foundation (grant nr 2024-1262). M.G.L. was supported by Doris Duke Foundation (Award 2023-0224) and US Department of Veterans Affairs Biomedical Research and Development Award IK2-BX006551.

## Authors’ contributions

S.Y., E.B., S.M.D., and N.J.L. conceived and designed the study. S.Y., E.B., G.S., T.D., J.A.L., K.C., P.T., S.M.D. contributed to data collection. S.Y. and E.B. undertook the statistical analyses. S.Y. and E.B. created the data visualizations and authored the initial draft of the manuscript. S.Y., E.B., G.S., T.D., J.A.L., R.E.T., H.S.L., A.D., K.C., P.T., S.A., M.G.L., S.M.D. and N.J.L. contributed to data interpretation, offered significant intellectual insights to the manuscript, and approved its final version.

## Data availability

The MVP individual-level data are open to VA-affiliated researchers via application (https://www.mvp.va.gov/pwa/mvp-data-available-research).

