## Supplementary Tables for "Triglyceride-rich lipoproteins, low-density lipoproteins, and risk of abdominal aortic aneurysm"

### **Table S1. Genetic instrumental variables used to proxy triglyceride-rich lipoproteins cholesterol, low-density lipoprotein cholesterol, and their ratio clusters**

| **SNP** | **ALLELE0** | **ALLELE1** | **Gene** | **Cluster** | **BETA_APOB** | **BETA_REMNCHOL** | **BETA_LDL** | **SE_APOB** | **SE_REMNCHOL** | **SE_LDL** | **P_APOB** | **P_REMNCHOL** | **P_LDL** |
| --- | --- | --- | --- | --- | --- | --- | --- | --- | --- | --- | --- | --- | --- |
| rs1002935 | A | G | HBP1 | 1 | 0.004 | 0.001 | 0.005 | 0.001 | 0.001 | 0.002 | 7.96E-14 | 2.81E-01 | 1.36E-02 |
| rs1029388 | C | T | ATXN2 | 1 | -0.004 | -0.001 | -0.019 | 0.001 | 0.001 | 0.002 | 4.75E-11 | 9.29E-02 | 3.19E-19 |
| rs10419669 | A | G | CBLC | 1 | -0.008 | 0.000 | -0.019 | 0.001 | 0.001 | 0.003 | 2.63E-18 | 8.24E-01 | 1.13E-08 |
| rs1081101 | T | C | APOE_TOMM40 | 1 | 0.048 | -0.006 | 0.107 | 0.008 | 0.010 | 0.028 | 1.76E-10 | 5.78E-01 | 1.34E-04 |
| rs108499 | T | C | MYRF | 1 | 0.013 | 0.000 | 0.025 | 0.000 | 0.001 | 0.002 | 6.50E-150 | 6.51E-01 | 5.37E-42 |
| rs10881645 | G | A | KIF20B | 1 | -0.003 | -0.001 | -0.007 | 0.000 | 0.001 | 0.002 | 3.18E-09 | 3.31E-02 | 9.94E-05 |
| rs11038642 | T | C | TRIM5 | 1 | -0.005 | 0.000 | -0.010 | 0.001 | 0.001 | 0.002 | 1.91E-14 | 7.95E-01 | 2.91E-05 |
| rs11080055 | C | A | TMEM97 | 1 | -0.004 | 0.000 | -0.012 | 0.000 | 0.001 | 0.002 | 1.56E-19 | 9.46E-01 | 1.04E-12 |
| rs11083761 | A | G | NKPD1_MARK4 | 1 | 0.081 | 0.017 | 0.193 | 0.003 | 0.005 | 0.012 | 1.50E-127 | 2.82E-04 | 4.57E-54 |
| rs111282584 | C | T | SPC24 | 1 | -0.013 | -0.004 | -0.042 | 0.002 | 0.003 | 0.008 | 1.49E-09 | 1.95E-01 | 1.76E-07 |
| rs112354933 | C | G | RAB3D | 1 | 0.010 | 0.003 | 0.031 | 0.001 | 0.002 | 0.005 | 9.32E-14 | 6.74E-02 | 1.03E-10 |
| rs11244084 | T | C | SURF6_ABO | 1 | -0.010 | -0.003 | -0.034 | 0.001 | 0.001 | 0.003 | 1.16E-30 | 1.93E-02 | 8.82E-25 |
| rs1133190 | G | A | CKM | 1 | 0.004 | 0.001 | 0.010 | 0.000 | 0.001 | 0.002 | 8.58E-13 | 6.14E-02 | 1.54E-08 |
| rs114036675 | A | G | BCL3 | 1 | -0.032 | -0.011 | -0.069 | 0.005 | 0.006 | 0.016 | 1.48E-12 | 7.78E-02 | 2.09E-05 |
| rs115357389 | T | C | GDF7_HS1BP3 | 1 | 0.014 | 0.004 | 0.031 | 0.002 | 0.003 | 0.008 | 8.28E-11 | 1.36E-01 | 1.23E-04 |
| rs115394954 | C | T | PPAP2B_MIR4422 | 1 | 0.009 | 0.003 | 0.025 | 0.002 | 0.002 | 0.006 | 7.87E-09 | 1.86E-01 | 1.43E-05 |
| rs11554038 | C | A | TMEM97 | 1 | 0.006 | 0.002 | 0.016 | 0.001 | 0.001 | 0.003 | 8.94E-13 | 4.88E-02 | 7.70E-07 |
| rs11555891 | A | G | IRGC | 1 | 0.012 | 0.001 | 0.023 | 0.002 | 0.002 | 0.006 | 2.43E-13 | 7.18E-01 | 1.31E-04 |
| rs11645898 | C | T | PMFBP1_ZFHX3 | 1 | -0.003 | -0.001 | -0.013 | 0.001 | 0.001 | 0.002 | 5.93E-06 | 9.63E-02 | 2.07E-08 |
| rs117142879 | T | C | CBLC_BCAM | 1 | -0.012 | -0.003 | -0.034 | 0.002 | 0.002 | 0.006 | 1.11E-11 | 2.31E-01 | 1.04E-07 |
| rs117490455 | G | A | SMARCA4 | 1 | -0.014 | -0.003 | -0.036 | 0.002 | 0.002 | 0.006 | 1.37E-15 | 1.51E-01 | 8.10E-09 |
| rs11768465 | T | C | FBXO24_PCOLCE-AS1 | 1 | 0.003 | 0.001 | 0.011 | 0.001 | 0.001 | 0.002 | 2.18E-10 | 2.99E-01 | 5.67E-08 |
| rs1229984 | C | T | ADH1B | 1 | -0.009 | -0.001 | -0.041 | 0.001 | 0.002 | 0.005 | 3.05E-11 | 5.80E-01 | 1.24E-15 |
| rs12526480 | G | T | LRRC16A | 1 | 0.003 | 0.001 | 0.010 | 0.000 | 0.001 | 0.002 | 1.46E-08 | 1.88E-01 | 8.69E-08 |
| rs12609269 | C | T | CEACAM20 | 1 | 0.015 | 0.002 | 0.034 | 0.001 | 0.001 | 0.003 | 1.03E-65 | 4.56E-02 | 1.33E-25 |
| rs12710745 | G | A | APOB_C2orf43 | 1 | 0.009 | 0.002 | 0.028 | 0.000 | 0.001 | 0.002 | 1.99E-85 | 6.43E-03 | 2.79E-57 |
| rs12721109 | A | G | APOC2_APOC4 | 1 | 0.119 | 0.035 | 0.290 | 0.002 | 0.002 | 0.006 | 0.00E+00 | 7.06E-64 | 0.00E+00 |
| rs12976464 | A | G | GIPR_EML2 | 1 | 0.012 | 0.002 | 0.033 | 0.001 | 0.001 | 0.004 | 3.36E-33 | 1.91E-01 | 1.72E-18 |
| rs13301187 | A | C | OSTF1_MIR548H3 | 1 | -0.004 | -0.001 | -0.012 | 0.001 | 0.001 | 0.002 | 1.47E-10 | 1.52E-01 | 6.82E-08 |
| rs1363907 | A | G | ERAP2 | 1 | -0.002 | 0.000 | -0.007 | 0.000 | 0.001 | 0.002 | 2.10E-07 | 4.79E-01 | 1.06E-04 |
| rs1389742 | G | A | MARC1 | 1 | -0.004 | -0.001 | -0.012 | 0.000 | 0.001 | 0.002 | 3.90E-14 | 4.07E-01 | 6.42E-12 |
| rs139659653 | A | G | NPC1L1 | 1 | 0.029 | 0.012 | 0.102 | 0.007 | 0.009 | 0.026 | 1.07E-05 | 2.14E-01 | 6.41E-05 |
| rs139745869 | A | G | PHLDB3 | 1 | 0.014 | 0.003 | 0.036 | 0.002 | 0.003 | 0.007 | 1.40E-12 | 2.20E-01 | 3.51E-07 |
| rs1434579 | T | C | ZNF229 | 1 | -0.005 | 0.000 | -0.011 | 0.001 | 0.001 | 0.002 | 1.37E-24 | 5.47E-01 | 1.65E-09 |
| rs148790687 | T | C | DNM2 | 1 | 0.025 | 0.005 | 0.084 | 0.005 | 0.007 | 0.018 | 2.51E-07 | 4.59E-01 | 2.87E-06 |
| rs1594895 | C | T | ZFP112_ZNF285 | 1 | 0.010 | 0.002 | 0.024 | 0.001 | 0.001 | 0.002 | 1.23E-58 | 1.19E-02 | 4.07E-28 |
| rs1661169 | C | T | CEACAM22P | 1 | -0.009 | -0.003 | -0.022 | 0.001 | 0.001 | 0.003 | 2.57E-40 | 6.18E-03 | 1.10E-18 |
| rs1692120 | A | G | RPLP0P2_DAGLA | 1 | -0.003 | 0.000 | -0.006 | 0.000 | 0.001 | 0.002 | 7.73E-12 | 4.83E-01 | 1.68E-04 |
| rs16980741 | T | C | SLC1A5_SNAR-E | 1 | 0.007 | 0.001 | 0.012 | 0.001 | 0.002 | 0.005 | 4.09E-08 | 6.88E-01 | 9.67E-03 |
| rs17031779 | C | T | LRPPRC | 1 | 0.008 | 0.001 | 0.024 | 0.001 | 0.002 | 0.006 | 2.39E-08 | 7.26E-01 | 1.38E-05 |
| rs17111479 | C | G | PCSK9_BSND | 1 | -0.004 | -0.002 | -0.013 | 0.001 | 0.001 | 0.002 | 2.02E-14 | 4.65E-02 | 2.75E-10 |
| rs17199964 | A | G | BANK1_FLJ20021 | 1 | 0.004 | 0.001 | 0.013 | 0.001 | 0.001 | 0.003 | 1.20E-05 | 3.42E-01 | 5.11E-05 |
| rs17200067 | T | C | HCP5_MICA | 1 | -0.007 | -0.004 | -0.030 | 0.001 | 0.002 | 0.005 | 6.41E-06 | 3.85E-02 | 2.90E-08 |
| rs1727731 | C | T | CEACAM20_CEACAM22P | 1 | 0.006 | 0.000 | 0.014 | 0.001 | 0.001 | 0.003 | 1.10E-11 | 9.12E-01 | 1.18E-05 |
| rs17406264 | T | C | THADA | 1 | 0.011 | 0.003 | 0.034 | 0.002 | 0.002 | 0.006 | 1.03E-11 | 2.46E-01 | 1.42E-07 |
| rs17496332 | G | A | PRMT6_LOC100129138 | 1 | -0.003 | -0.002 | -0.010 | 0.000 | 0.001 | 0.002 | 1.25E-07 | 2.43E-02 | 1.09E-07 |
| rs17565182 | C | T | POC5_SV2C | 1 | -0.006 | -0.002 | -0.016 | 0.001 | 0.001 | 0.003 | 2.01E-14 | 5.72E-02 | 3.41E-08 |
| rs17585355 | C | A | SORT1 | 1 | -0.008 | -0.001 | -0.021 | 0.001 | 0.001 | 0.004 | 8.98E-18 | 4.20E-01 | 1.18E-08 |
| rs1799943 | A | G | BRCA2 | 1 | 0.004 | 0.001 | 0.009 | 0.001 | 0.001 | 0.002 | 9.65E-15 | 1.87E-01 | 3.06E-06 |
| rs1799945 | G | C | HFE | 1 | 0.005 | 0.001 | 0.020 | 0.001 | 0.001 | 0.002 | 2.34E-16 | 1.11E-01 | 7.90E-16 |
| rs1800961 | T | C | HNF4A | 1 | 0.011 | 0.001 | 0.051 | 0.001 | 0.002 | 0.005 | 2.09E-16 | 5.60E-01 | 1.63E-24 |
| rs198472 | A | G | MYRF | 1 | -0.003 | 0.000 | -0.005 | 0.001 | 0.001 | 0.002 | 8.47E-07 | 5.10E-01 | 7.75E-03 |
| rs1998925 | C | T | SLC17A8 | 1 | 0.003 | 0.001 | 0.009 | 0.000 | 0.001 | 0.002 | 3.51E-09 | 7.72E-02 | 3.41E-07 |
| rs2027086 | A | G | LAMC1 | 1 | -0.002 | -0.001 | -0.008 | 0.000 | 0.001 | 0.002 | 3.16E-07 | 1.51E-01 | 1.08E-05 |
| rs2275492 | A | G | EYA3 | 1 | 0.002 | 0.000 | 0.006 | 0.001 | 0.001 | 0.002 | 6.84E-05 | 9.68E-01 | 1.28E-03 |
| rs2285673 | T | C | C5orf56 | 1 | -0.003 | -0.001 | -0.010 | 0.001 | 0.001 | 0.002 | 1.29E-08 | 4.04E-01 | 5.90E-06 |
| rs2535323 | G | A | IER3_DDR1 | 1 | -0.003 | -0.002 | -0.015 | 0.001 | 0.001 | 0.002 | 4.45E-05 | 7.45E-03 | 2.60E-10 |
| rs2618566 | T | G | SNX5_BANF2 | 1 | 0.010 | 0.003 | 0.024 | 0.000 | 0.001 | 0.002 | 1.51E-95 | 1.62E-06 | 1.71E-40 |
| rs270607 | G | A | SLC22A4_LOC553103 | 1 | -0.003 | 0.000 | -0.008 | 0.000 | 0.001 | 0.002 | 4.92E-12 | 5.96E-01 | 4.02E-05 |
| rs2736601 | T | C | BEST1_RAB3IL1 | 1 | 0.007 | 0.001 | 0.015 | 0.001 | 0.001 | 0.003 | 2.18E-13 | 2.79E-01 | 4.74E-06 |
| rs2885404 | C | T | WIPI1_PRKAR1A | 1 | -0.004 | -0.001 | -0.010 | 0.001 | 0.001 | 0.002 | 2.87E-13 | 3.83E-01 | 4.52E-07 |
| rs2965156 | C | G | CEACAM16_CEACAM19 | 1 | 0.012 | 0.004 | 0.029 | 0.000 | 0.001 | 0.002 | 1.44E-145 | 6.46E-10 | 1.28E-59 |
| rs2965174 | A | G | BCL3_CEACAM16 | 1 | 0.011 | 0.003 | 0.026 | 0.000 | 0.001 | 0.002 | 6.24E-108 | 1.87E-05 | 6.80E-50 |
| rs31670 | A | G | ABCB4 | 1 | -0.003 | 0.001 | -0.010 | 0.001 | 0.001 | 0.002 | 6.19E-09 | 4.49E-01 | 1.41E-06 |
| rs3219484 | T | C | MUTYH | 1 | 0.005 | 0.002 | 0.016 | 0.001 | 0.001 | 0.003 | 1.12E-09 | 1.89E-01 | 1.30E-06 |
| rs34924007 | C | T | S1PR5_ATG4D | 1 | 0.003 | 0.001 | 0.010 | 0.001 | 0.001 | 0.002 | 2.07E-08 | 2.98E-01 | 1.95E-06 |
| rs35350976 | G | A | SLC27A5 | 1 | -0.003 | -0.002 | -0.011 | 0.001 | 0.001 | 0.002 | 7.32E-08 | 7.10E-02 | 9.45E-07 |
| rs35891370 | A | G | RELB | 1 | -0.008 | -0.001 | -0.018 | 0.001 | 0.001 | 0.002 | 2.48E-60 | 1.05E-01 | 4.71E-23 |
| rs36043205 | A | G | KRT78_KRT8 | 1 | -0.003 | -0.001 | -0.006 | 0.000 | 0.001 | 0.002 | 2.33E-10 | 4.05E-01 | 1.23E-03 |
| rs360789 | T | C | EHBP1 | 1 | 0.005 | 0.002 | 0.014 | 0.001 | 0.001 | 0.002 | 2.86E-17 | 6.55E-02 | 4.70E-10 |
| rs364585 | G | A | SPTLC3_BTBD3 | 1 | -0.003 | -0.001 | -0.011 | 0.000 | 0.001 | 0.002 | 4.37E-10 | 4.97E-02 | 1.04E-09 |
| rs3750320 | C | T | LRRC8A | 1 | 0.004 | 0.001 | 0.013 | 0.001 | 0.001 | 0.002 | 2.91E-10 | 5.42E-01 | 1.55E-08 |
| rs3786507 | A | G | CLASRP | 1 | -0.005 | 0.000 | -0.012 | 0.000 | 0.001 | 0.002 | 2.38E-22 | 8.74E-01 | 7.13E-12 |
| rs3809775 | G | C | HOXB8_HOXB9 | 1 | -0.003 | -0.001 | -0.011 | 0.000 | 0.001 | 0.002 | 3.53E-14 | 5.20E-02 | 5.53E-11 |
| rs405509 | G | T | APOE_TOMM40 | 1 | 0.031 | 0.005 | 0.068 | 0.000 | 0.001 | 0.002 | 0.00E+00 | 5.80E-17 | 0.00E+00 |
| rs412334 | T | C | FEN1_TMEM258 | 1 | -0.007 | -0.001 | -0.017 | 0.001 | 0.001 | 0.002 | 6.57E-31 | 4.51E-01 | 4.93E-13 |
| rs41290122 | A | G | PVRL2 | 1 | -0.013 | -0.005 | -0.029 | 0.002 | 0.003 | 0.008 | 4.43E-09 | 8.99E-02 | 3.34E-04 |
| rs41302083 | T | C | PSRC1 | 1 | 0.023 | 0.004 | 0.048 | 0.003 | 0.004 | 0.010 | 1.64E-19 | 3.25E-01 | 7.63E-07 |
| rs4461181 | G | C | MARK4_EXOC3L2 | 1 | -0.004 | -0.001 | -0.010 | 0.001 | 0.001 | 0.003 | 4.11E-09 | 5.28E-01 | 6.46E-05 |
| rs4681675 | T | G | RPP14_ABHD6 | 1 | 0.003 | 0.001 | 0.009 | 0.000 | 0.001 | 0.002 | 2.78E-12 | 6.26E-02 | 2.65E-07 |
| rs4802260 | T | C | VASP | 1 | 0.005 | 0.002 | 0.015 | 0.001 | 0.001 | 0.002 | 3.50E-20 | 3.74E-03 | 7.95E-14 |
| rs4850047 | C | T | COLEC11_RPS7 | 1 | 0.005 | -0.001 | 0.015 | 0.001 | 0.001 | 0.003 | 1.49E-13 | 5.19E-01 | 4.85E-09 |
| rs505151 | A | G | PCSK9 | 1 | 0.022 | 0.008 | 0.062 | 0.001 | 0.002 | 0.005 | 3.09E-68 | 1.06E-05 | 1.06E-40 |
| rs56109496 | A | G | CARM1 | 1 | -0.006 | -0.001 | -0.013 | 0.001 | 0.001 | 0.004 | 6.75E-10 | 5.72E-01 | 5.47E-04 |
| rs56315738 | T | C | SMARCA4_LDLR | 1 | 0.024 | 0.012 | 0.080 | 0.002 | 0.003 | 0.009 | 5.91E-24 | 8.09E-05 | 1.40E-20 |
| rs57579470 | G | T | PPM1N_VASP | 1 | 0.007 | 0.000 | 0.015 | 0.001 | 0.001 | 0.003 | 2.83E-21 | 9.87E-01 | 3.74E-08 |
| rs57830821 | T | C | MLEC_CABP1 | 1 | -0.004 | -0.002 | -0.010 | 0.001 | 0.001 | 0.002 | 6.91E-09 | 5.59E-02 | 3.23E-05 |
| rs581107 | T | C | ABO | 1 | -0.003 | -0.001 | -0.014 | 0.000 | 0.001 | 0.002 | 3.59E-13 | 7.10E-02 | 4.46E-16 |
| rs6072355 | C | G | EMILIN3_CHD6 | 1 | -0.004 | -0.001 | -0.011 | 0.001 | 0.001 | 0.002 | 5.20E-15 | 1.42E-01 | 9.39E-09 |
| rs6111683 | G | C | BANF2_SNX5 | 1 | -0.004 | -0.002 | -0.013 | 0.001 | 0.001 | 0.002 | 3.91E-14 | 9.83E-03 | 1.68E-09 |
| rs61993987 | G | A | PPP2R5C | 1 | -0.003 | -0.001 | -0.012 | 0.001 | 0.001 | 0.003 | 4.63E-07 | 1.20E-01 | 2.52E-06 |
| rs62116303 | G | A | ZNF229_ZNF180 | 1 | 0.016 | 0.004 | 0.037 | 0.001 | 0.001 | 0.003 | 4.38E-119 | 1.22E-04 | 4.36E-47 |
| rs62119261 | C | A | IGSF23_CEACAM22P | 1 | 0.043 | 0.009 | 0.101 | 0.001 | 0.002 | 0.004 | 1.95E-303 | 1.35E-09 | 1.00E-127 |
| rs62119318 | A | G | PVR_CEACAM19 | 1 | -0.012 | -0.003 | -0.029 | 0.001 | 0.001 | 0.004 | 4.37E-30 | 4.89E-02 | 3.57E-14 |
| rs62401414 | G | A | OR2B2_HIST1H2BO | 1 | -0.009 | -0.006 | -0.029 | 0.002 | 0.003 | 0.007 | 7.84E-06 | 3.03E-02 | 9.28E-05 |
| rs62640397 | C | T | FDX1L | 1 | 0.006 | 0.004 | 0.024 | 0.001 | 0.002 | 0.004 | 2.15E-07 | 1.85E-02 | 1.44E-07 |
| rs6495122 | C | A | CPLX3_ULK3 | 1 | -0.002 | -0.001 | -0.013 | 0.000 | 0.001 | 0.002 | 4.61E-07 | 2.57E-01 | 5.96E-13 |
| rs6662108 | A | C | RPRD2 | 1 | 0.003 | 0.000 | 0.010 | 0.000 | 0.001 | 0.002 | 1.35E-11 | 8.34E-01 | 5.72E-08 |
| rs6750700 | A | C | PRKCE | 1 | 0.003 | 0.000 | 0.007 | 0.000 | 0.001 | 0.002 | 3.64E-08 | 6.81E-01 | 2.88E-04 |
| rs6960320 | G | A | MIR148A_RNU6-16P | 1 | -0.005 | 0.000 | -0.011 | 0.001 | 0.001 | 0.003 | 4.36E-12 | 9.37E-01 | 1.04E-04 |
| rs7221192 | T | C | HOXB1_SKAP1 | 1 | 0.007 | 0.000 | 0.021 | 0.001 | 0.001 | 0.003 | 1.19E-14 | 7.81E-01 | 1.31E-10 |
| rs7255060 | T | C | PPP1R37_MARK4 | 1 | -0.006 | -0.003 | -0.017 | 0.001 | 0.001 | 0.003 | 1.59E-11 | 1.81E-02 | 2.83E-08 |
| rs72703257 | T | C | PSMA5 | 1 | -0.011 | -0.001 | -0.018 | 0.001 | 0.002 | 0.005 | 1.30E-14 | 6.18E-01 | 4.96E-04 |
| rs72749953 | T | C | IRF2BP2_LOC100506795 | 1 | -0.004 | -0.002 | -0.012 | 0.001 | 0.001 | 0.002 | 2.20E-11 | 2.99E-02 | 8.85E-08 |
| rs73572039 | T | C | PVRL2_BCAM | 1 | -0.011 | -0.003 | -0.029 | 0.001 | 0.001 | 0.003 | 2.18E-47 | 5.91E-03 | 1.33E-24 |
| rs74327448 | G | A | SET | 1 | 0.007 | 0.002 | 0.025 | 0.001 | 0.002 | 0.005 | 5.32E-07 | 2.58E-01 | 6.82E-07 |
| rs74544477 | C | T | ZNF229 | 1 | -0.007 | -0.002 | -0.020 | 0.001 | 0.002 | 0.004 | 3.19E-09 | 2.15E-01 | 2.52E-06 |
| rs7481842 | T | C | FADS2_FADS3 | 1 | -0.007 | -0.001 | -0.014 | 0.001 | 0.001 | 0.003 | 1.10E-20 | 1.81E-01 | 1.34E-07 |
| rs74852793 | G | A | LOC100507562_APOB | 1 | 0.010 | 0.003 | 0.022 | 0.001 | 0.001 | 0.004 | 3.63E-21 | 5.84E-02 | 4.73E-09 |
| rs75816342 | A | G | MARK4 | 1 | -0.008 | -0.003 | -0.023 | 0.001 | 0.001 | 0.004 | 5.14E-14 | 1.89E-02 | 1.41E-09 |
| rs75902389 | T | C | USP24 | 1 | 0.008 | 0.001 | 0.019 | 0.001 | 0.001 | 0.004 | 2.61E-13 | 3.63E-01 | 1.27E-06 |
| rs76240114 | C | A | UQCRHL_FBLIM1 | 1 | -0.004 | -0.001 | -0.010 | 0.001 | 0.001 | 0.002 | 3.16E-12 | 1.37E-01 | 7.86E-06 |
| rs7631792 | T | C | SMC4 | 1 | 0.003 | 0.001 | 0.007 | 0.000 | 0.001 | 0.002 | 8.73E-09 | 3.65E-01 | 6.66E-05 |
| rs76473094 | T | C | MIR148A_NFE2L3 | 1 | -0.007 | 0.004 | -0.019 | 0.002 | 0.002 | 0.006 | 6.69E-06 | 3.57E-02 | 1.05E-03 |
| rs76560105 | T | G | CBLC | 1 | 0.078 | 0.017 | 0.186 | 0.001 | 0.002 | 0.005 | 0.00E+00 | 1.63E-21 | 0.00E+00 |
| rs76797241 | C | G | ZNF223_ZNF222 | 1 | 0.018 | 0.002 | 0.051 | 0.002 | 0.003 | 0.008 | 2.99E-17 | 3.86E-01 | 6.06E-11 |
| rs7718341 | G | A | ANKRD31_GCNT4 | 1 | -0.005 | -0.002 | -0.016 | 0.001 | 0.001 | 0.002 | 4.21E-22 | 2.08E-02 | 2.51E-16 |
| rs77196615 | C | T | PVRL2 | 1 | -0.013 | -0.004 | -0.035 | 0.002 | 0.003 | 0.007 | 6.32E-12 | 1.29E-01 | 5.04E-07 |
| rs77231091 | A | G | MAP2K6_KCNJ16 | 1 | -0.014 | -0.005 | -0.041 | 0.001 | 0.002 | 0.005 | 8.47E-26 | 8.13E-03 | 1.97E-16 |
| rs77400396 | A | G | HMBOX1 | 1 | 0.008 | 0.005 | 0.030 | 0.001 | 0.002 | 0.005 | 2.23E-09 | 1.07E-02 | 1.48E-08 |
| rs77704739 | C | T | ISL1_PELO | 1 | 0.015 | 0.003 | 0.038 | 0.001 | 0.002 | 0.004 | 1.03E-40 | 4.82E-02 | 4.71E-18 |
| rs7857390 | G | A | ABO_OBP2B | 1 | -0.004 | -0.001 | -0.016 | 0.000 | 0.001 | 0.002 | 8.94E-14 | 8.09E-02 | 3.47E-18 |
| rs7868232 | C | T | GBGT1_OBP2B | 1 | -0.005 | -0.001 | -0.017 | 0.001 | 0.001 | 0.002 | 1.06E-17 | 1.68E-01 | 2.85E-17 |
| rs78755596 | A | T | ABO_OBP2B | 1 | -0.011 | -0.003 | -0.041 | 0.001 | 0.002 | 0.005 | 1.10E-13 | 1.17E-01 | 7.01E-14 |
| rs78852738 | C | A | CELSR2 | 1 | -0.009 | -0.005 | -0.030 | 0.001 | 0.002 | 0.005 | 8.08E-12 | 9.02E-03 | 4.92E-09 |
| rs79106033 | T | C | APOB_C2orf43 | 1 | 0.011 | 0.002 | 0.034 | 0.001 | 0.002 | 0.005 | 9.04E-14 | 4.02E-01 | 4.64E-10 |
| rs79146351 | C | A | GDF7_HS1BP3 | 1 | -0.006 | 0.000 | -0.011 | 0.001 | 0.001 | 0.003 | 1.06E-11 | 8.99E-01 | 7.41E-04 |
| rs79187649 | C | T | SYN2 | 1 | 0.003 | 0.000 | 0.012 | 0.001 | 0.001 | 0.002 | 3.63E-08 | 8.26E-01 | 6.19E-09 |
| rs79327334 | G | A | FAM118B | 1 | 0.014 | 0.004 | 0.031 | 0.002 | 0.004 | 0.009 | 3.32E-08 | 2.78E-01 | 8.33E-04 |
| rs7935946 | T | C | FADS2 | 1 | 0.008 | -0.001 | 0.015 | 0.001 | 0.001 | 0.004 | 7.21E-16 | 7.30E-01 | 1.17E-04 |
| rs79385701 | C | A | CEACAM22P_IGSF23 | 1 | -0.009 | -0.003 | -0.020 | 0.001 | 0.002 | 0.005 | 1.10E-11 | 1.20E-01 | 3.02E-05 |
| rs79860339 | C | G | CEACAM20_CEACAM22P | 1 | 0.011 | 0.002 | 0.019 | 0.001 | 0.002 | 0.005 | 5.34E-15 | 2.79E-01 | 1.23E-04 |
| rs846866 | C | A | IGSF23 | 1 | -0.007 | 0.000 | -0.014 | 0.001 | 0.001 | 0.002 | 1.98E-24 | 6.93E-01 | 6.72E-09 |
| rs9306894 | G | A | C2orf43_GDF7 | 1 | 0.003 | 0.001 | 0.011 | 0.000 | 0.001 | 0.002 | 1.34E-09 | 3.01E-01 | 5.38E-09 |
| rs9396646 | A | G | GMPR_MYLIP | 1 | 0.004 | 0.000 | 0.010 | 0.001 | 0.001 | 0.002 | 8.32E-13 | 5.81E-01 | 5.47E-06 |
| rs9467715 | C | T | BTN3A2_HIST1H4H | 1 | 0.003 | 0.001 | 0.010 | 0.001 | 0.001 | 0.002 | 7.91E-08 | 2.81E-01 | 9.99E-08 |
| rs1006896 | C | A | CGGBP1 | 2 | -0.005 | -0.002 | -0.013 | 0.001 | 0.001 | 0.003 | 1.38E-10 | 4.66E-02 | 2.89E-06 |
| rs10166144 | G | A | APOB_LOC645949 | 2 | 0.027 | 0.012 | 0.068 | 0.001 | 0.001 | 0.003 | 0.00E+00 | 1.21E-34 | 2.74E-157 |
| rs10205003 | T | C | APOB_LOC645949 | 2 | 0.018 | 0.007 | 0.045 | 0.002 | 0.003 | 0.007 | 4.15E-19 | 1.52E-02 | 1.22E-09 |
| rs10415983 | T | C | MARK4_EXOC3L2 | 2 | -0.007 | -0.002 | -0.014 | 0.001 | 0.001 | 0.002 | 2.74E-31 | 1.49E-02 | 4.98E-10 |
| rs10462509 | G | T | FAM169A_GCNT4 | 2 | -0.007 | -0.004 | -0.023 | 0.001 | 0.002 | 0.004 | 1.52E-10 | 2.54E-02 | 1.89E-07 |
| rs10876168 | T | C | GALNT6 | 2 | 0.002 | 0.001 | 0.006 | 0.000 | 0.001 | 0.002 | 9.74E-08 | 5.19E-02 | 2.57E-04 |
| rs10888897 | C | T | PCSK9 | 2 | -0.009 | -0.004 | -0.026 | 0.000 | 0.001 | 0.002 | 3.07E-86 | 1.09E-11 | 2.97E-49 |
| rs10951261 | A | G | COX19_ADAP1 | 2 | 0.003 | 0.001 | 0.009 | 0.001 | 0.001 | 0.002 | 1.26E-10 | 1.18E-01 | 1.07E-06 |
| rs11038628 | T | C | TRIM5 | 2 | 0.009 | 0.003 | 0.020 | 0.001 | 0.001 | 0.003 | 2.70E-22 | 2.65E-02 | 4.81E-09 |
| rs11206510 | C | T | PCSK9_BSND | 2 | 0.015 | 0.007 | 0.039 | 0.001 | 0.001 | 0.002 | 1.12E-133 | 3.72E-16 | 3.63E-69 |
| rs11230735 | T | C | SYT7 | 2 | -0.004 | -0.001 | -0.010 | 0.001 | 0.001 | 0.002 | 3.65E-13 | 2.35E-01 | 9.59E-06 |
| rs113337987 | A | G | MTTP | 2 | 0.008 | 0.006 | 0.029 | 0.002 | 0.002 | 0.006 | 9.31E-08 | 5.47E-03 | 7.51E-07 |
| rs113408695 | C | T | ABCA6_ABCA9 | 2 | -0.022 | -0.010 | -0.069 | 0.001 | 0.002 | 0.005 | 7.56E-75 | 5.35E-09 | 8.18E-51 |
| rs114166723 | A | G | SARS | 2 | 0.011 | 0.006 | 0.032 | 0.002 | 0.002 | 0.006 | 3.46E-11 | 1.19E-02 | 9.64E-08 |
| rs114374491 | G | A | KLHL29_LOC645949 | 2 | 0.011 | 0.004 | 0.024 | 0.002 | 0.002 | 0.006 | 1.48E-12 | 7.42E-02 | 8.67E-05 |
| rs114533012 | C | T | FAM169A_GCNT4 | 2 | -0.009 | -0.006 | -0.027 | 0.001 | 0.002 | 0.005 | 2.17E-13 | 1.39E-03 | 1.66E-08 |
| rs115030251 | T | C | MYBPHL | 2 | 0.007 | 0.003 | 0.017 | 0.001 | 0.001 | 0.003 | 1.87E-24 | 3.75E-03 | 1.13E-11 |
| rs11545166 | G | T | KRI1 | 2 | -0.009 | -0.005 | -0.028 | 0.001 | 0.002 | 0.005 | 2.17E-11 | 9.84E-03 | 1.19E-08 |
| rs115458560 | C | T | AMIGO1_GPR61 | 2 | 0.016 | 0.007 | 0.041 | 0.002 | 0.002 | 0.006 | 4.54E-20 | 5.81E-03 | 1.70E-10 |
| rs11603023 | C | T | PHLDB1 | 2 | 0.003 | 0.001 | 0.009 | 0.000 | 0.001 | 0.002 | 2.09E-08 | 5.61E-02 | 4.06E-07 |
| rs116102380 | A | G | PPAP2B_MIR4422 | 2 | 0.010 | 0.004 | 0.028 | 0.002 | 0.002 | 0.006 | 5.95E-09 | 6.74E-02 | 1.80E-05 |
| rs11688414 | A | G | TMEM163 | 2 | 0.003 | 0.002 | 0.007 | 0.000 | 0.001 | 0.002 | 1.46E-07 | 2.34E-02 | 1.25E-04 |
| rs117110139 | A | G | TMEM258 | 2 | 0.009 | 0.001 | 0.020 | 0.001 | 0.002 | 0.005 | 7.77E-13 | 4.62E-01 | 9.79E-06 |
| rs11763759 | C | T | NPC1L1 | 2 | 0.003 | 0.003 | 0.013 | 0.001 | 0.001 | 0.002 | 8.41E-12 | 2.05E-04 | 4.79E-11 |
| rs117753658 | G | A | CHST4_ZNF19 | 2 | 0.013 | 0.007 | 0.043 | 0.002 | 0.002 | 0.007 | 5.81E-14 | 2.13E-03 | 5.44E-11 |
| rs117783250 | C | T | TXNL4B | 2 | 0.010 | 0.004 | 0.033 | 0.002 | 0.002 | 0.006 | 1.43E-09 | 6.75E-02 | 2.11E-07 |
| rs11810371 | A | G | PCSK9_BSND | 2 | 0.008 | 0.004 | 0.023 | 0.001 | 0.001 | 0.004 | 1.58E-14 | 1.28E-02 | 6.49E-09 |
| rs11849883 | G | A | ENTPD5 | 2 | -0.003 | -0.002 | -0.009 | 0.000 | 0.001 | 0.002 | 7.50E-12 | 7.31E-03 | 1.58E-06 |
| rs11879798 | A | G | KCNN4 | 2 | 0.004 | 0.001 | 0.008 | 0.000 | 0.001 | 0.002 | 4.24E-14 | 8.61E-02 | 3.55E-06 |
| rs11881756 | C | T | CEACAM16_BCL3 | 2 | 0.041 | 0.013 | 0.099 | 0.001 | 0.001 | 0.003 | 0.00E+00 | 3.47E-38 | 2.43E-284 |
| rs12031153 | A | G | USP24 | 2 | -0.008 | -0.003 | -0.018 | 0.001 | 0.001 | 0.004 | 7.94E-14 | 2.13E-02 | 1.67E-06 |
| rs12329383 | C | T | WDPCP | 2 | -0.003 | -0.001 | -0.008 | 0.000 | 0.001 | 0.002 | 1.40E-08 | 1.92E-01 | 6.23E-06 |
| rs12467951 | G | T | OTX1_DBIL5P2 | 2 | -0.004 | -0.001 | -0.009 | 0.001 | 0.001 | 0.002 | 1.96E-10 | 1.96E-01 | 3.01E-05 |
| rs12582170 | G | A | SP7_SP1 | 2 | 0.005 | 0.001 | 0.007 | 0.001 | 0.001 | 0.002 | 8.33E-13 | 1.87E-01 | 2.12E-03 |
| rs12611268 | T | C | ERCC1_FOSB | 2 | 0.004 | 0.002 | 0.011 | 0.001 | 0.001 | 0.002 | 4.56E-12 | 3.40E-02 | 8.52E-07 |
| rs12713559 | A | G | APOB | 2 | -0.088 | -0.035 | -0.252 | 0.007 | 0.010 | 0.026 | 1.08E-34 | 2.71E-04 | 1.40E-21 |
| rs12732125 | T | C | BSND | 2 | 0.029 | 0.016 | 0.087 | 0.002 | 0.002 | 0.006 | 1.88E-64 | 4.30E-11 | 7.95E-43 |
| rs12803635 | A | G | CABP4 | 2 | 0.007 | 0.002 | 0.014 | 0.001 | 0.001 | 0.004 | 1.56E-11 | 1.22E-01 | 5.07E-04 |
| rs12983573 | C | T | SLC44A2_AP1M2 | 2 | -0.005 | -0.003 | -0.014 | 0.001 | 0.001 | 0.002 | 8.08E-19 | 1.80E-04 | 5.81E-14 |
| rs13301006 | T | C | ABCA1 | 2 | 0.002 | 0.001 | 0.014 | 0.001 | 0.001 | 0.002 | 1.13E-02 | 9.56E-02 | 6.23E-09 |
| rs142501705 | T | A | TRAPPC6A_MARK4 | 2 | -0.035 | -0.009 | -0.058 | 0.004 | 0.006 | 0.015 | 9.27E-16 | 1.04E-01 | 1.72E-04 |
| rs144787122 | G | A | SNX8 | 2 | -0.030 | -0.016 | -0.106 | 0.004 | 0.005 | 0.014 | 9.19E-16 | 2.42E-03 | 1.14E-13 |
| rs16894128 | G | A | GPX6_ZSCAN23 | 2 | 0.004 | 0.001 | 0.015 | 0.001 | 0.001 | 0.003 | 6.75E-10 | 1.70E-01 | 1.88E-08 |
| rs1689800 | G | A | ZNF648_GLUL | 2 | -0.003 | -0.002 | -0.010 | 0.000 | 0.001 | 0.002 | 5.88E-13 | 7.57E-03 | 3.08E-08 |
| rs16988864 | A | T | MAFB_LOC339568 | 2 | -0.004 | -0.003 | -0.013 | 0.001 | 0.001 | 0.003 | 5.92E-08 | 4.80E-03 | 5.16E-07 |
| rs17111792 | G | A | PPAP2B_MIR4422 | 2 | -0.007 | -0.003 | -0.019 | 0.001 | 0.001 | 0.004 | 5.11E-11 | 5.79E-02 | 8.79E-07 |
| rs17242346 | A | G | LDLR_SMARCA4 | 2 | -0.013 | -0.006 | -0.039 | 0.001 | 0.001 | 0.004 | 1.07E-39 | 8.29E-06 | 1.29E-26 |
| rs17508548 | G | T | GABBR1_OR2H2 | 2 | -0.005 | -0.003 | -0.020 | 0.001 | 0.001 | 0.003 | 4.53E-13 | 3.65E-04 | 2.32E-15 |
| rs17660635 | G | A | LOC157273_TNKS | 2 | 0.004 | 0.002 | 0.014 | 0.001 | 0.001 | 0.002 | 5.46E-12 | 1.94E-02 | 1.04E-10 |
| rs1805738 | G | A | PHC1 | 2 | -0.004 | -0.003 | -0.015 | 0.001 | 0.001 | 0.002 | 1.49E-14 | 4.10E-04 | 1.94E-12 |
| rs181860403 | T | G | RHCE | 2 | 0.007 | 0.004 | 0.022 | 0.001 | 0.001 | 0.002 | 3.20E-49 | 3.51E-08 | 1.13E-31 |
| rs2066714 | C | T | ABCA1 | 2 | -0.003 | -0.003 | -0.017 | 0.001 | 0.001 | 0.003 | 7.97E-05 | 2.16E-03 | 8.86E-12 |
| rs2239620 | G | A | LOC730101_TRAM2-AS1 | 2 | -0.003 | -0.001 | -0.007 | 0.000 | 0.001 | 0.002 | 6.67E-12 | 2.03E-01 | 1.96E-04 |
| rs2275166 | G | A | CLCNKB | 2 | -0.003 | -0.002 | -0.007 | 0.000 | 0.001 | 0.002 | 1.58E-10 | 1.80E-02 | 3.03E-04 |
| rs2294264 | T | C | MYLIP_DTNBP1 | 2 | 0.005 | 0.002 | 0.014 | 0.000 | 0.001 | 0.002 | 1.75E-30 | 2.03E-03 | 7.04E-16 |
| rs2303291 | T | C | ITSN2 | 2 | -0.003 | -0.001 | -0.009 | 0.001 | 0.001 | 0.002 | 2.63E-10 | 5.16E-02 | 6.26E-06 |
| rs2317677 | A | G | ITGB3_EFCAB13 | 2 | -0.006 | -0.002 | -0.016 | 0.000 | 0.001 | 0.002 | 3.18E-36 | 4.65E-04 | 5.15E-20 |
| rs2618582 | G | A | SNX5_BANF2 | 2 | -0.007 | -0.003 | -0.016 | 0.001 | 0.001 | 0.003 | 2.18E-13 | 5.15E-02 | 3.88E-06 |
| rs267733 | G | A | ANXA9 | 2 | 0.005 | 0.002 | 0.012 | 0.001 | 0.001 | 0.002 | 2.50E-14 | 6.04E-02 | 7.17E-07 |
| rs2682563 | C | T | ZNF575_ETHE1 | 2 | 0.003 | 0.002 | 0.009 | 0.000 | 0.001 | 0.002 | 1.13E-11 | 1.75E-02 | 1.35E-06 |
| rs2738458 | C | T | LDLR | 2 | 0.007 | 0.002 | 0.019 | 0.001 | 0.001 | 0.002 | 2.58E-36 | 3.23E-03 | 2.33E-23 |
| rs2760058 | T | C | RABGAP1L | 2 | -0.003 | -0.001 | -0.010 | 0.001 | 0.001 | 0.002 | 8.27E-09 | 6.29E-02 | 1.46E-06 |
| rs2807835 | T | G | MARC1 | 2 | -0.003 | -0.001 | -0.010 | 0.001 | 0.001 | 0.002 | 1.24E-10 | 4.91E-02 | 4.77E-07 |
| rs28678581 | C | T | IRF2BP1_FOXA3 | 2 | 0.004 | 0.002 | 0.015 | 0.001 | 0.001 | 0.002 | 2.64E-10 | 3.04E-03 | 1.43E-11 |
| rs2965109 | T | C | CEACAM16_BCL3 | 2 | 0.018 | 0.006 | 0.041 | 0.000 | 0.001 | 0.002 | 2.37E-278 | 1.24E-17 | 2.48E-111 |
| rs2972557 | A | G | PVRL2 | 2 | 0.010 | 0.003 | 0.019 | 0.001 | 0.001 | 0.003 | 2.28E-37 | 4.87E-03 | 6.40E-12 |
| rs3094246 | G | A | POLK | 2 | 0.008 | 0.005 | 0.025 | 0.001 | 0.002 | 0.004 | 3.45E-12 | 1.89E-03 | 3.18E-09 |
| rs3129736 | T | C | HLA-DQB1_HLA-DQA2 | 2 | -0.003 | -0.002 | -0.011 | 0.001 | 0.001 | 0.002 | 9.89E-09 | 3.50E-02 | 4.04E-08 |
| rs314675 | C | T | PRKD2 | 2 | -0.004 | -0.002 | -0.011 | 0.001 | 0.001 | 0.003 | 1.83E-07 | 5.60E-02 | 1.52E-05 |
| rs34243815 | T | C | DOCK6 | 2 | 0.006 | 0.002 | 0.009 | 0.001 | 0.001 | 0.003 | 2.74E-10 | 1.36E-01 | 6.50E-03 |
| rs34353535 | A | G | CEACAM20_CEACAM22P | 2 | 0.008 | 0.003 | 0.018 | 0.001 | 0.001 | 0.003 | 1.58E-17 | 3.91E-02 | 3.38E-08 |
| rs34777587 | G | A | PVRL2 | 2 | 0.043 | 0.019 | 0.137 | 0.007 | 0.009 | 0.026 | 1.05E-09 | 4.66E-02 | 1.35E-07 |
| rs349030 | A | G | ZNF283 | 2 | -0.003 | -0.001 | -0.007 | 0.000 | 0.001 | 0.002 | 4.34E-11 | 4.08E-01 | 1.20E-04 |
| rs35127135 | T | C | APOB_LOC645949 | 2 | -0.007 | -0.003 | -0.017 | 0.001 | 0.001 | 0.003 | 3.20E-26 | 8.85E-03 | 6.76E-11 |
| rs35803101 | A | G | NPC1L1 | 2 | 0.023 | 0.011 | 0.067 | 0.003 | 0.005 | 0.013 | 1.56E-11 | 2.82E-02 | 5.28E-07 |
| rs3772102 | G | T | ST3GAL6 | 2 | 0.003 | 0.002 | 0.007 | 0.000 | 0.001 | 0.002 | 1.00E-12 | 8.32E-03 | 4.66E-05 |
| rs3805939 | G | C | PTK7 | 2 | 0.003 | 0.000 | 0.004 | 0.000 | 0.001 | 0.002 | 2.05E-09 | 9.22E-01 | 2.32E-02 |
| rs3852856 | A | G | PVRL2 | 2 | -0.017 | -0.006 | -0.045 | 0.001 | 0.001 | 0.002 | 4.42E-153 | 1.12E-12 | 2.64E-83 |
| rs4468734 | A | G | CEACAM22P_IGSF23 | 2 | -0.003 | -0.001 | -0.008 | 0.000 | 0.001 | 0.002 | 8.56E-12 | 5.38E-02 | 2.19E-06 |
| rs45564645 | G | T | ZNF563 | 2 | -0.012 | -0.002 | -0.029 | 0.002 | 0.002 | 0.007 | 2.63E-10 | 4.61E-01 | 2.18E-05 |
| rs471705 | G | T | PCSK9 | 2 | -0.012 | -0.006 | -0.036 | 0.000 | 0.001 | 0.002 | 6.71E-135 | 1.69E-17 | 1.09E-86 |
| rs4788815 | T | A | LOC100132529_TAT | 2 | -0.005 | -0.003 | -0.015 | 0.000 | 0.001 | 0.002 | 2.96E-23 | 1.53E-04 | 8.57E-16 |
| rs4804564 | T | C | SMARCA4 | 2 | -0.008 | -0.003 | -0.024 | 0.001 | 0.001 | 0.003 | 3.79E-17 | 6.04E-03 | 2.70E-12 |
| rs4899343 | G | A | COX16_ADAM21P1 | 2 | -0.004 | -0.002 | -0.008 | 0.001 | 0.001 | 0.002 | 3.60E-15 | 2.15E-02 | 1.24E-05 |
| rs4942486 | C | T | BRCA2 | 2 | 0.007 | 0.003 | 0.015 | 0.000 | 0.001 | 0.002 | 9.11E-57 | 7.50E-05 | 8.49E-19 |
| rs526936 | A | G | LINC00184_LOC100506810 | 2 | -0.010 | -0.005 | -0.030 | 0.000 | 0.001 | 0.002 | 6.08E-106 | 3.47E-16 | 8.29E-65 |
| rs55660224 | T | C | MYBPHL | 2 | 0.027 | 0.011 | 0.065 | 0.001 | 0.001 | 0.003 | 5.66E-228 | 1.02E-19 | 4.99E-96 |
| rs56029521 | C | T | DNAH11 | 2 | -0.005 | -0.002 | -0.012 | 0.000 | 0.001 | 0.002 | 3.17E-26 | 3.38E-03 | 1.21E-11 |
| rs61872264 | T | G | FLJ46361 | 2 | -0.003 | -0.001 | -0.008 | 0.000 | 0.001 | 0.002 | 2.48E-08 | 9.67E-02 | 7.11E-06 |
| rs62116988 | A | G | KCNN4_LYPD5 | 2 | 0.020 | 0.008 | 0.049 | 0.001 | 0.002 | 0.005 | 6.13E-52 | 9.81E-06 | 6.84E-24 |
| rs62124178 | T | C | HS1BP3_RHOB | 2 | -0.006 | -0.003 | -0.017 | 0.001 | 0.001 | 0.004 | 3.18E-08 | 7.80E-02 | 1.11E-05 |
| rs62366598 | T | C | COL4A3BP | 2 | 0.007 | 0.004 | 0.023 | 0.001 | 0.001 | 0.003 | 1.17E-15 | 3.74E-03 | 1.42E-11 |
| rs62513000 | G | T | TRPS1 | 2 | 0.005 | 0.002 | 0.015 | 0.001 | 0.001 | 0.003 | 9.88E-11 | 4.33E-02 | 3.73E-07 |
| rs6511721 | A | G | LDLR | 2 | 0.014 | 0.007 | 0.042 | 0.000 | 0.001 | 0.002 | 6.30E-190 | 1.28E-24 | 1.52E-123 |
| rs6531257 | A | G | APOB_C2orf43 | 2 | 0.012 | 0.005 | 0.029 | 0.001 | 0.001 | 0.003 | 7.24E-56 | 1.47E-06 | 5.63E-25 |
| rs657361 | A | G | FAM118B | 2 | 0.003 | 0.001 | 0.008 | 0.001 | 0.001 | 0.002 | 7.80E-11 | 7.55E-02 | 5.94E-05 |
| rs6867855 | G | T | CSNK1G3 | 2 | -0.004 | -0.002 | -0.013 | 0.001 | 0.001 | 0.002 | 1.16E-13 | 3.60E-02 | 1.94E-10 |
| rs687416 | T | C | PDS5B | 2 | 0.003 | 0.001 | 0.005 | 0.000 | 0.001 | 0.002 | 2.10E-10 | 3.36E-01 | 2.64E-03 |
| rs7136570 | G | A | CERS5_COX14 | 2 | -0.003 | -0.002 | -0.010 | 0.000 | 0.001 | 0.002 | 1.38E-08 | 5.77E-03 | 3.43E-08 |
| rs71435583 | G | A | C2orf43 | 2 | 0.016 | 0.006 | 0.041 | 0.001 | 0.001 | 0.004 | 1.53E-52 | 7.11E-06 | 1.71E-26 |
| rs7178424 | T | C | C2CD4B_C2CD4A | 2 | -0.003 | -0.002 | -0.009 | 0.000 | 0.001 | 0.002 | 1.09E-09 | 1.70E-02 | 2.99E-07 |
| rs7198936 | C | T | PKD1L3 | 2 | -0.003 | -0.002 | -0.008 | 0.000 | 0.001 | 0.002 | 6.32E-13 | 1.19E-02 | 2.58E-06 |
| rs72647804 | G | A | SORT1 | 2 | -0.009 | -0.004 | -0.022 | 0.001 | 0.001 | 0.004 | 2.16E-15 | 5.01E-03 | 3.92E-08 |
| rs72658867 | A | G | LDLR | 2 | 0.073 | 0.037 | 0.222 | 0.002 | 0.003 | 0.008 | 5.60E-251 | 1.21E-35 | 6.73E-168 |
| rs72701097 | T | C | SARS | 2 | -0.014 | -0.005 | -0.027 | 0.002 | 0.003 | 0.008 | 4.60E-10 | 1.10E-01 | 8.10E-04 |
| rs72702343 | T | C | PPP4R4_IFI27L2 | 2 | -0.008 | -0.004 | -0.022 | 0.001 | 0.002 | 0.004 | 1.71E-12 | 9.48E-03 | 9.07E-08 |
| rs72756554 | C | T | ITGA1 | 2 | 0.003 | 0.002 | 0.011 | 0.001 | 0.001 | 0.002 | 3.31E-09 | 3.63E-02 | 9.87E-08 |
| rs72796772 | T | C | ABCG8 | 2 | 0.009 | 0.006 | 0.031 | 0.001 | 0.002 | 0.005 | 7.85E-12 | 4.08E-04 | 5.87E-11 |
| rs72800933 | T | G | PPM1B_LRPPRC | 2 | -0.006 | -0.003 | -0.019 | 0.001 | 0.001 | 0.004 | 1.97E-09 | 1.99E-02 | 9.34E-08 |
| rs72801786 | T | C | IST1 | 2 | -0.006 | -0.003 | -0.013 | 0.001 | 0.001 | 0.003 | 8.74E-17 | 4.62E-03 | 7.02E-07 |
| rs72828723 | T | C | LRRC16A_CMAHP | 2 | 0.006 | 0.002 | 0.018 | 0.001 | 0.001 | 0.003 | 2.13E-12 | 6.10E-02 | 8.52E-09 |
| rs73015030 | A | G | LDLR | 2 | 0.031 | 0.016 | 0.099 | 0.001 | 0.002 | 0.005 | 1.17E-126 | 4.03E-20 | 9.63E-97 |
| rs73070437 | G | A | DNAH11 | 2 | -0.010 | -0.006 | -0.028 | 0.001 | 0.001 | 0.003 | 1.77E-27 | 1.16E-05 | 1.01E-15 |
| rs73496517 | T | C | ATG4D | 2 | -0.008 | -0.004 | -0.025 | 0.001 | 0.001 | 0.003 | 2.75E-19 | 2.19E-03 | 7.55E-16 |
| rs73936968 | A | G | TOMM40 | 2 | -0.013 | -0.006 | -0.038 | 0.002 | 0.002 | 0.006 | 7.27E-17 | 7.33E-03 | 4.92E-11 |
| rs7421942 | T | C | LRPPRC_PPM1B | 2 | -0.004 | -0.003 | -0.015 | 0.001 | 0.001 | 0.003 | 6.58E-08 | 7.45E-03 | 2.37E-08 |
| rs74257940 | G | T | LDLR | 2 | -0.019 | -0.007 | -0.048 | 0.002 | 0.003 | 0.008 | 8.69E-19 | 1.38E-02 | 2.94E-10 |
| rs74607435 | C | T | BCL3_CEACAM16 | 2 | 0.023 | 0.008 | 0.052 | 0.001 | 0.001 | 0.004 | 3.59E-97 | 3.75E-07 | 5.95E-37 |
| rs74942924 | A | G | SMARCA4_C19orf52 | 2 | 0.025 | 0.010 | 0.071 | 0.001 | 0.002 | 0.005 | 9.02E-82 | 1.03E-09 | 3.07E-51 |
| rs7523242 | T | C | PCSK9_BSND | 2 | -0.012 | -0.007 | -0.036 | 0.001 | 0.001 | 0.002 | 6.10E-92 | 1.48E-15 | 4.48E-60 |
| rs7534498 | A | G | KIAA1324 | 2 | 0.006 | 0.002 | 0.012 | 0.001 | 0.001 | 0.002 | 5.62E-22 | 3.60E-02 | 6.30E-08 |
| rs75452015 | T | C | PMFBP1_ZFHX3 | 2 | 0.006 | 0.003 | 0.017 | 0.001 | 0.001 | 0.002 | 3.59E-20 | 4.39E-04 | 1.42E-12 |
| rs769448 | T | C | APOE | 2 | -0.014 | -0.006 | -0.038 | 0.002 | 0.002 | 0.006 | 2.47E-16 | 9.18E-03 | 2.39E-10 |
| rs77908346 | T | C | MAFB_LOC339568 | 2 | 0.004 | 0.003 | 0.014 | 0.001 | 0.001 | 0.003 | 2.07E-10 | 7.47E-03 | 1.85E-07 |
| rs7862078 | C | T | MIR548H3 | 2 | -0.002 | -0.001 | -0.007 | 0.000 | 0.001 | 0.002 | 2.25E-06 | 2.47E-02 | 3.04E-05 |
| rs7941030 | C | T | UBASH3B_MIR100HG | 2 | -0.003 | -0.002 | -0.011 | 0.000 | 0.001 | 0.002 | 7.90E-09 | 1.00E-02 | 2.42E-10 |
| rs79765293 | A | G | CILP2 | 2 | -0.008 | -0.002 | -0.020 | 0.001 | 0.002 | 0.005 | 3.76E-10 | 2.36E-01 | 2.63E-05 |
| rs80341249 | C | T | LOC100507562_APOB | 2 | 0.020 | 0.006 | 0.035 | 0.002 | 0.003 | 0.009 | 8.59E-18 | 4.56E-02 | 4.70E-05 |
| rs8057016 | T | G | DHODH | 2 | 0.006 | 0.001 | 0.015 | 0.001 | 0.002 | 0.004 | 1.71E-08 | 3.42E-01 | 2.87E-04 |
| rs8614 | A | C | NUFIP2 | 2 | -0.004 | -0.002 | -0.013 | 0.001 | 0.001 | 0.002 | 2.25E-09 | 1.93E-02 | 1.51E-08 |
| rs928911 | T | C | PPP1R13L | 2 | -0.008 | -0.002 | -0.019 | 0.001 | 0.002 | 0.004 | 4.30E-13 | 1.41E-01 | 8.58E-06 |
| rs9330459 | G | T | TMEM8C_SLC2A6 | 2 | 0.007 | 0.002 | 0.028 | 0.001 | 0.002 | 0.004 | 1.51E-09 | 1.38E-01 | 5.98E-10 |
| rs936165 | T | G | DNAJC13 | 2 | -0.004 | -0.001 | -0.009 | 0.001 | 0.001 | 0.002 | 1.39E-13 | 1.36E-01 | 1.15E-06 |
| rs9616810 | T | C | ARSA_SHANK3 | 2 | -0.003 | -0.001 | -0.007 | 0.001 | 0.001 | 0.002 | 1.78E-09 | 3.10E-01 | 4.94E-04 |
| rs9697571 | G | A | TAF13_WDR47 | 2 | -0.006 | -0.002 | -0.013 | 0.000 | 0.001 | 0.002 | 2.19E-33 | 1.62E-03 | 5.24E-14 |
| rs9911208 | C | T | MAP2K6_KCNJ16 | 2 | -0.004 | -0.002 | -0.011 | 0.001 | 0.001 | 0.002 | 3.52E-13 | 3.69E-03 | 5.06E-08 |
| rs10420888 | G | A | EXOC3L2_MARK4 | 3 | -0.010 | -0.005 | -0.025 | 0.001 | 0.001 | 0.003 | 8.88E-33 | 1.68E-05 | 3.76E-17 |
| rs10474446 | T | C | POC5_SV2C | 3 | -0.013 | -0.010 | -0.046 | 0.001 | 0.002 | 0.005 | 8.18E-27 | 1.45E-08 | 7.55E-23 |
| rs10771185 | G | A | A2ML1_PHC1 | 3 | 0.003 | 0.003 | 0.013 | 0.001 | 0.001 | 0.002 | 6.42E-07 | 8.45E-05 | 2.75E-08 |
| rs10791660 | A | C | PDGFD | 3 | 0.004 | 0.002 | 0.014 | 0.001 | 0.001 | 0.002 | 2.37E-12 | 7.02E-03 | 6.95E-10 |
| rs10888908 | C | T | PPAP2B_MIR4422 | 3 | -0.011 | -0.006 | -0.033 | 0.001 | 0.001 | 0.003 | 3.17E-35 | 1.21E-06 | 2.90E-25 |
| rs11065079 | T | C | PLA2G1B_SIRT4 | 3 | -0.007 | -0.004 | -0.027 | 0.002 | 0.002 | 0.006 | 4.01E-05 | 7.56E-02 | 1.22E-05 |
| rs11101976 | C | T | AMPD2_GSTM4 | 3 | -0.004 | -0.002 | -0.009 | 0.000 | 0.001 | 0.002 | 2.29E-15 | 2.31E-03 | 1.47E-07 |
| rs11102001 | A | G | EPS8L3 | 3 | 0.011 | 0.005 | 0.025 | 0.001 | 0.001 | 0.004 | 6.41E-32 | 4.43E-05 | 1.88E-12 |
| rs11220462 | A | G | ST3GAL4 | 3 | -0.011 | -0.007 | -0.032 | 0.001 | 0.001 | 0.003 | 6.22E-60 | 4.59E-12 | 1.96E-36 |
| rs1127343 | A | G | FAM162A | 3 | 0.003 | 0.002 | 0.008 | 0.000 | 0.001 | 0.002 | 3.64E-10 | 6.47E-03 | 1.18E-06 |
| rs114185526 | T | C | APOB_LOC645949 | 3 | 0.032 | 0.015 | 0.083 | 0.002 | 0.003 | 0.009 | 3.70E-46 | 1.72E-06 | 3.73E-22 |
| rs114523982 | C | A | FAM169A_GCNT4 | 3 | -0.012 | -0.007 | -0.038 | 0.002 | 0.003 | 0.007 | 2.94E-11 | 1.24E-02 | 4.24E-08 |
| rs114770276 | G | C | THADA | 3 | 0.013 | 0.008 | 0.041 | 0.002 | 0.003 | 0.007 | 3.94E-12 | 2.88E-03 | 2.06E-08 |
| rs115035890 | T | C | DOCK6 | 3 | 0.040 | 0.024 | 0.131 | 0.005 | 0.007 | 0.020 | 3.46E-13 | 1.03E-03 | 8.25E-11 |
| rs11571788 | T | C | BRCA2 | 3 | 0.009 | 0.003 | 0.016 | 0.001 | 0.002 | 0.004 | 6.94E-15 | 7.09E-02 | 3.81E-04 |
| rs11591147 | T | G | PCSK9 | 3 | 0.099 | 0.054 | 0.303 | 0.002 | 0.002 | 0.007 | 0.00E+00 | 2.32E-102 | 0.00E+00 |
| rs11601507 | A | C | TRIM5 | 3 | -0.014 | -0.006 | -0.031 | 0.001 | 0.001 | 0.003 | 1.75E-57 | 1.13E-06 | 2.01E-20 |
| rs116378305 | T | G | PPAP2B_MIR4422 | 3 | -0.017 | -0.008 | -0.044 | 0.002 | 0.003 | 0.009 | 2.62E-11 | 1.34E-02 | 1.80E-06 |
| rs11668327 | C | G | TOMM40 | 3 | 0.051 | 0.022 | 0.127 | 0.001 | 0.001 | 0.002 | 0.00E+00 | 1.64E-132 | 0.00E+00 |
| rs116958755 | G | T | MARK4_EXOC3L2 | 3 | -0.015 | -0.008 | -0.037 | 0.001 | 0.002 | 0.005 | 5.01E-31 | 3.65E-06 | 3.08E-14 |
| rs117706747 | C | T | ZNF821 | 3 | 0.017 | 0.009 | 0.049 | 0.002 | 0.002 | 0.006 | 4.45E-23 | 1.66E-04 | 2.11E-14 |
| rs118163475 | T | C | YIPF2 | 3 | -0.020 | -0.013 | -0.056 | 0.002 | 0.003 | 0.009 | 4.93E-17 | 4.69E-05 | 8.11E-11 |
| rs11881101 | G | A | RELB | 3 | 0.007 | 0.004 | 0.018 | 0.001 | 0.001 | 0.002 | 8.29E-34 | 4.85E-07 | 1.47E-15 |
| rs11897825 | G | A | LOC100507562_APOB | 3 | -0.005 | -0.002 | -0.012 | 0.000 | 0.001 | 0.002 | 2.52E-29 | 3.22E-04 | 1.97E-12 |
| rs12052058 | T | G | SMARCA4 | 3 | 0.017 | 0.008 | 0.044 | 0.001 | 0.001 | 0.002 | 1.04E-196 | 1.77E-27 | 3.13E-104 |
| rs12143028 | C | G | USP24 | 3 | -0.007 | -0.003 | -0.019 | 0.001 | 0.001 | 0.003 | 4.13E-24 | 2.55E-04 | 1.37E-13 |
| rs12336893 | G | T | VLDLR_FLJ35024 | 3 | 0.007 | 0.004 | 0.022 | 0.001 | 0.001 | 0.004 | 2.08E-13 | 6.85E-03 | 1.72E-09 |
| rs12713843 | T | C | APOB | 3 | 0.070 | 0.029 | 0.162 | 0.003 | 0.005 | 0.013 | 4.02E-100 | 3.46E-10 | 1.50E-37 |
| rs12720358 | T | C | TYK2 | 3 | 0.017 | 0.008 | 0.048 | 0.002 | 0.002 | 0.006 | 1.38E-24 | 2.17E-04 | 6.53E-15 |
| rs12720359 | G | C | TYK2 | 3 | 0.016 | 0.011 | 0.043 | 0.001 | 0.002 | 0.005 | 7.56E-36 | 3.76E-10 | 6.16E-21 |
| rs12924903 | G | A | FBXL19-AS1_CTF1 | 3 | 0.003 | 0.002 | 0.008 | 0.000 | 0.001 | 0.002 | 6.65E-10 | 4.64E-03 | 8.61E-06 |
| rs13178545 | C | T | CSNK1G3_CEP120 | 3 | 0.004 | 0.002 | 0.010 | 0.001 | 0.001 | 0.002 | 2.76E-10 | 2.56E-02 | 3.35E-06 |
| rs13189797 | T | G | ANKRD31 | 3 | 0.006 | 0.003 | 0.017 | 0.001 | 0.001 | 0.004 | 5.49E-10 | 3.04E-02 | 6.62E-06 |
| rs13277838 | T | C | DMTN | 3 | -0.005 | -0.003 | -0.013 | 0.001 | 0.001 | 0.002 | 8.13E-21 | 1.79E-03 | 8.02E-09 |
| rs140717526 | A | G | SARS | 3 | 0.024 | 0.010 | 0.051 | 0.002 | 0.003 | 0.008 | 2.56E-32 | 5.11E-04 | 2.20E-11 |
| rs144130656 | A | G | DHX38 | 3 | 0.012 | 0.009 | 0.042 | 0.002 | 0.003 | 0.009 | 1.46E-07 | 7.81E-03 | 1.27E-06 |
| rs148657245 | A | G | CHST4_ZNF19 | 3 | 0.014 | 0.008 | 0.043 | 0.002 | 0.003 | 0.008 | 6.88E-11 | 7.12E-03 | 2.49E-07 |
| rs1529711 | T | C | CARM1 | 3 | -0.009 | -0.005 | -0.025 | 0.001 | 0.001 | 0.002 | 1.07E-37 | 2.54E-07 | 2.29E-25 |
| rs16872961 | C | T | POC5_SV2C | 3 | 0.004 | 0.002 | 0.013 | 0.001 | 0.001 | 0.002 | 1.40E-12 | 1.64E-02 | 3.23E-08 |
| rs16926246 | T | C | HK1 | 3 | 0.006 | 0.004 | 0.017 | 0.001 | 0.001 | 0.003 | 3.87E-21 | 9.25E-05 | 8.06E-12 |
| rs16937390 | C | T | DENND4C_PLIN2 | 3 | -0.006 | -0.003 | -0.014 | 0.001 | 0.001 | 0.003 | 4.55E-12 | 3.39E-02 | 9.94E-06 |
| rs17014532 | G | T | KIAA1324 | 3 | 0.007 | 0.003 | 0.018 | 0.001 | 0.001 | 0.002 | 1.25E-34 | 1.17E-05 | 3.94E-19 |
| rs17248748 | T | C | LDLR | 3 | 0.023 | 0.012 | 0.060 | 0.002 | 0.003 | 0.007 | 3.41E-31 | 7.69E-06 | 3.81E-17 |
| rs17336182 | C | T | PLEKHH2 | 3 | 0.009 | 0.005 | 0.027 | 0.001 | 0.001 | 0.003 | 3.38E-27 | 2.16E-04 | 1.76E-16 |
| rs1801698 | C | T | APOB | 3 | 0.034 | 0.018 | 0.077 | 0.004 | 0.005 | 0.014 | 1.70E-19 | 6.70E-04 | 1.02E-07 |
| rs1865452 | T | C | UBXN4 | 3 | -0.004 | -0.002 | -0.011 | 0.001 | 0.001 | 0.003 | 4.60E-10 | 1.46E-02 | 1.88E-05 |
| rs221798 | G | C | GIGYF1_POP7 | 3 | 0.005 | 0.003 | 0.017 | 0.001 | 0.001 | 0.003 | 3.44E-13 | 1.55E-03 | 1.91E-10 |
| rs2264779 | T | C | C12orf43 | 3 | -0.003 | -0.002 | -0.010 | 0.000 | 0.001 | 0.002 | 7.27E-09 | 1.69E-02 | 2.35E-07 |
| rs2287623 | A | G | ABCB11 | 3 | 0.005 | 0.004 | 0.019 | 0.000 | 0.001 | 0.002 | 5.88E-31 | 1.03E-08 | 4.48E-27 |
| rs2727261 | C | T | BEST1_RAB3IL1 | 3 | -0.006 | -0.001 | -0.010 | 0.001 | 0.001 | 0.003 | 2.49E-16 | 2.31E-01 | 3.16E-04 |
| rs28399637 | A | G | BCAM | 3 | -0.027 | -0.011 | -0.065 | 0.001 | 0.001 | 0.002 | 0.00E+00 | 1.15E-58 | 1.55E-258 |
| rs2972831 | T | C | SV2C | 3 | -0.004 | -0.003 | -0.012 | 0.001 | 0.001 | 0.003 | 5.24E-09 | 5.45E-03 | 7.76E-06 |
| rs3124747 | G | A | C9orf96 | 3 | 0.003 | 0.002 | 0.014 | 0.000 | 0.001 | 0.002 | 1.28E-12 | 1.90E-03 | 1.43E-13 |
| rs34457396 | G | T | SLC44A2 | 3 | -0.006 | -0.003 | -0.014 | 0.001 | 0.001 | 0.004 | 1.28E-07 | 5.31E-02 | 3.07E-04 |
| rs34495400 | T | C | KRI1 | 3 | 0.013 | 0.007 | 0.040 | 0.002 | 0.003 | 0.007 | 2.95E-11 | 6.74E-03 | 1.72E-08 |
| rs34605587 | G | T | PPAP2B_MIR4422 | 3 | 0.010 | 0.006 | 0.030 | 0.001 | 0.001 | 0.004 | 1.05E-25 | 9.25E-06 | 1.41E-15 |
| rs346764 | A | G | EXOC3L2_MARK4 | 3 | -0.006 | -0.003 | -0.017 | 0.001 | 0.001 | 0.003 | 1.95E-11 | 1.31E-02 | 4.42E-07 |
| rs35882350 | G | A | B4GALNT3 | 3 | -0.005 | -0.002 | -0.012 | 0.001 | 0.001 | 0.002 | 8.18E-20 | 8.95E-04 | 4.75E-10 |
| rs36005514 | A | G | LDLR_SMARCA4 | 3 | -0.013 | -0.007 | -0.038 | 0.001 | 0.001 | 0.003 | 4.43E-44 | 2.51E-07 | 2.74E-28 |
| rs36086613 | A | G | CALB2 | 3 | 0.004 | 0.002 | 0.013 | 0.000 | 0.001 | 0.002 | 2.62E-15 | 1.09E-03 | 3.40E-13 |
| rs379309 | T | C | KANK2 | 3 | 0.007 | 0.003 | 0.016 | 0.000 | 0.001 | 0.002 | 1.41E-43 | 4.45E-07 | 1.21E-20 |
| rs3826804 | G | A | DNM2 | 3 | 0.011 | 0.006 | 0.029 | 0.001 | 0.001 | 0.002 | 9.92E-92 | 1.59E-16 | 1.30E-52 |
| rs387976 | C | A | PVRL2 | 3 | 0.023 | 0.010 | 0.056 | 0.001 | 0.001 | 0.002 | 0.00E+00 | 3.75E-45 | 5.52E-186 |
| rs3916847 | C | T | ERCC2 | 3 | 0.011 | 0.006 | 0.031 | 0.001 | 0.002 | 0.005 | 3.00E-19 | 6.71E-04 | 7.28E-12 |
| rs41276062 | T | C | TINAG | 3 | 0.003 | 0.003 | 0.013 | 0.001 | 0.001 | 0.003 | 7.39E-07 | 1.68E-03 | 1.28E-06 |
| rs4131228 | G | A | ABCG5 | 3 | -0.009 | -0.005 | -0.026 | 0.002 | 0.002 | 0.006 | 1.86E-08 | 1.17E-02 | 3.22E-06 |
| rs4149268 | T | C | ABCA1 | 3 | 0.002 | 0.002 | 0.011 | 0.000 | 0.001 | 0.002 | 3.70E-05 | 1.55E-04 | 5.46E-10 |
| rs4302748 | A | G | EEPD1_SEPT7 | 3 | -0.003 | -0.003 | -0.013 | 0.001 | 0.001 | 0.002 | 2.98E-09 | 9.68E-04 | 5.07E-09 |
| rs4423040 | T | G | FNDC7 | 3 | 0.003 | 0.001 | 0.007 | 0.000 | 0.001 | 0.002 | 9.08E-14 | 2.38E-02 | 1.07E-04 |
| rs4661674 | G | A | ZBTB17 | 3 | 0.003 | 0.002 | 0.008 | 0.000 | 0.001 | 0.002 | 1.32E-11 | 7.45E-03 | 2.07E-05 |
| rs4689635 | C | T | SORCS2 | 3 | -0.003 | -0.002 | -0.009 | 0.000 | 0.001 | 0.002 | 3.45E-08 | 2.76E-03 | 1.32E-06 |
| rs4703645 | C | T | ANKRD31_GCNT4 | 3 | -0.016 | -0.010 | -0.049 | 0.001 | 0.001 | 0.002 | 1.52E-139 | 4.84E-28 | 1.14E-95 |
| rs4729561 | C | T | AZGP1P1_ZKSCAN1 | 3 | 0.003 | 0.001 | 0.008 | 0.000 | 0.001 | 0.002 | 5.58E-12 | 1.89E-01 | 2.74E-05 |
| rs4791641 | T | C | PFAS | 3 | 0.002 | 0.001 | 0.008 | 0.000 | 0.001 | 0.002 | 1.42E-06 | 1.42E-01 | 8.01E-06 |
| rs4804159 | G | A | TSPAN16_DOCK6 | 3 | -0.005 | -0.003 | -0.016 | 0.001 | 0.001 | 0.002 | 3.24E-20 | 5.63E-07 | 2.66E-17 |
| rs4808762 | C | T | PDE4C | 3 | -0.003 | -0.002 | -0.012 | 0.001 | 0.001 | 0.002 | 5.61E-07 | 6.49E-04 | 5.77E-10 |
| rs518076 | G | A | GNAI3 | 3 | 0.010 | 0.005 | 0.022 | 0.001 | 0.001 | 0.003 | 1.30E-41 | 8.55E-06 | 7.71E-16 |
| rs56041751 | T | C | ST3GAL2 | 3 | 0.007 | 0.006 | 0.025 | 0.001 | 0.002 | 0.005 | 8.12E-07 | 3.20E-03 | 3.71E-06 |
| rs56078519 | G | A | C2orf43 | 3 | -0.021 | -0.011 | -0.058 | 0.001 | 0.002 | 0.004 | 2.21E-69 | 1.24E-10 | 2.95E-38 |
| rs56245770 | C | A | FAM169A_GCNT4 | 3 | -0.010 | -0.007 | -0.031 | 0.002 | 0.002 | 0.006 | 4.80E-11 | 1.71E-03 | 4.78E-08 |
| rs599951 | T | C | FGF21_BCAT2 | 3 | 0.003 | 0.002 | 0.007 | 0.001 | 0.001 | 0.002 | 5.28E-07 | 3.05E-02 | 1.34E-03 |
| rs608511 | T | C | SARS | 3 | 0.007 | 0.003 | 0.015 | 0.001 | 0.001 | 0.002 | 6.34E-38 | 2.86E-04 | 5.81E-15 |
| rs60960031 | A | G | APOB_LOC645949 | 3 | 0.012 | 0.006 | 0.030 | 0.000 | 0.001 | 0.002 | 5.82E-143 | 1.71E-21 | 9.89E-64 |
| rs61797139 | T | C | SYPL2_PSMA5 | 3 | -0.008 | -0.004 | -0.020 | 0.001 | 0.001 | 0.003 | 8.02E-23 | 1.81E-04 | 3.18E-10 |
| rs62022813 | G | A | LITAF_LOC388210 | 3 | 0.003 | 0.001 | 0.010 | 0.000 | 0.001 | 0.002 | 6.58E-09 | 1.24E-01 | 2.53E-07 |
| rs62623708 | A | C | CELSR2 | 3 | 0.029 | 0.012 | 0.064 | 0.003 | 0.004 | 0.011 | 3.05E-22 | 5.01E-03 | 1.89E-08 |
| rs6537837 | T | C | GNAI3 | 3 | -0.005 | -0.003 | -0.014 | 0.001 | 0.001 | 0.002 | 2.35E-20 | 1.82E-04 | 7.48E-10 |
| rs655246 | G | A | PSRC1_MYBPHL | 3 | -0.013 | -0.005 | -0.030 | 0.000 | 0.001 | 0.002 | 2.85E-175 | 1.04E-16 | 2.46E-67 |
| rs6697526 | T | C | SCCPDH_CNST | 3 | -0.005 | -0.002 | -0.008 | 0.001 | 0.001 | 0.003 | 5.38E-09 | 8.43E-02 | 4.35E-03 |
| rs6947410 | G | A | SP4_RPL23P8 | 3 | -0.003 | -0.002 | -0.008 | 0.000 | 0.001 | 0.002 | 2.67E-13 | 1.15E-02 | 1.29E-06 |
| rs7013120 | A | G | TCEB1 | 3 | 0.003 | 0.001 | 0.009 | 0.000 | 0.001 | 0.002 | 9.13E-11 | 3.32E-02 | 7.42E-07 |
| rs7209433 | G | A | CACNG1_HELZ | 3 | -0.003 | -0.001 | -0.005 | 0.000 | 0.001 | 0.002 | 4.65E-14 | 2.24E-02 | 1.76E-03 |
| rs7247903 | G | A | CYP2A6_CYP2A7 | 3 | 0.006 | 0.003 | 0.015 | 0.001 | 0.001 | 0.003 | 1.23E-09 | 1.32E-02 | 1.54E-05 |
| rs72626215 | A | G | DMWD | 3 | -0.003 | -0.001 | -0.007 | 0.001 | 0.001 | 0.002 | 3.49E-08 | 4.79E-02 | 2.32E-04 |
| rs72703203 | A | G | CELSR2 | 3 | 0.037 | 0.015 | 0.084 | 0.001 | 0.002 | 0.004 | 1.06E-219 | 3.74E-21 | 4.11E-82 |
| rs72749998 | C | A | LOC148638_LINC00184 | 3 | 0.008 | 0.005 | 0.025 | 0.001 | 0.002 | 0.004 | 8.46E-11 | 5.72E-03 | 1.20E-08 |
| rs72785837 | C | T | APOB_LOC645949 | 3 | -0.010 | -0.005 | -0.031 | 0.001 | 0.001 | 0.004 | 4.85E-21 | 2.44E-04 | 2.06E-16 |
| rs72790077 | G | C | CAMKMT | 3 | 0.012 | 0.008 | 0.045 | 0.002 | 0.002 | 0.007 | 1.37E-12 | 5.60E-04 | 7.51E-12 |
| rs72798837 | T | G | ABCG8 | 3 | -0.010 | -0.006 | -0.031 | 0.001 | 0.001 | 0.004 | 6.02E-23 | 6.36E-05 | 4.42E-15 |
| rs72833416 | G | A | MYLIP_DTNBP1 | 3 | 0.005 | 0.005 | 0.019 | 0.001 | 0.001 | 0.004 | 4.21E-07 | 4.18E-04 | 5.67E-08 |
| rs72850341 | A | G | ABCA6_ABCA9 | 3 | 0.012 | 0.007 | 0.036 | 0.002 | 0.002 | 0.006 | 2.67E-12 | 2.72E-03 | 1.10E-08 |
| rs72902579 | C | T | APOB_LOC645949 | 3 | 0.033 | 0.015 | 0.072 | 0.001 | 0.002 | 0.004 | 1.31E-196 | 2.60E-21 | 9.27E-66 |
| rs73035978 | C | T | ZNF230_ZNF222 | 3 | -0.006 | -0.004 | -0.019 | 0.001 | 0.001 | 0.003 | 1.33E-15 | 2.98E-04 | 4.24E-11 |
| rs73111353 | A | G | MAFB_LOC339568 | 3 | -0.007 | -0.005 | -0.022 | 0.001 | 0.002 | 0.005 | 3.23E-08 | 2.24E-03 | 1.20E-06 |
| rs74758151 | G | A | KRAS_IFLTD1 | 3 | 0.007 | 0.004 | 0.014 | 0.001 | 0.001 | 0.003 | 1.33E-14 | 1.35E-03 | 5.40E-05 |
| rs747632 | T | C | TECPR1 | 3 | 0.003 | 0.001 | 0.006 | 0.000 | 0.001 | 0.002 | 9.40E-11 | 3.74E-02 | 2.69E-04 |
| rs74964185 | G | A | GCNT4_ANKRD31 | 3 | 0.006 | 0.004 | 0.020 | 0.001 | 0.001 | 0.003 | 3.03E-17 | 7.95E-06 | 4.21E-15 |
| rs75161053 | A | G | CEACAM22P | 3 | -0.012 | -0.005 | -0.030 | 0.001 | 0.002 | 0.004 | 2.12E-26 | 8.81E-04 | 5.88E-13 |
| rs75919641 | C | A | STXBP3 | 3 | 0.015 | 0.006 | 0.036 | 0.002 | 0.002 | 0.007 | 7.34E-17 | 1.50E-02 | 6.43E-08 |
| rs7616785 | G | A | CSTA_CCDC58 | 3 | -0.003 | -0.001 | -0.007 | 0.000 | 0.001 | 0.002 | 1.76E-12 | 2.34E-02 | 9.00E-05 |
| rs76218671 | C | T | SP4_RPL23P8 | 3 | -0.009 | -0.003 | -0.017 | 0.001 | 0.002 | 0.005 | 4.72E-13 | 1.42E-01 | 3.31E-04 |
| rs76267242 | T | G | HBS1L_MYB | 3 | -0.004 | -0.003 | -0.009 | 0.001 | 0.001 | 0.003 | 5.09E-07 | 1.31E-02 | 1.00E-03 |
| rs76810866 | T | C | ABCA9 | 3 | 0.006 | 0.005 | 0.022 | 0.001 | 0.001 | 0.004 | 5.77E-10 | 1.33E-03 | 1.93E-08 |
| rs76971643 | A | G | TRAPPC6A_MARK4 | 3 | 0.014 | 0.007 | 0.038 | 0.001 | 0.002 | 0.004 | 4.31E-37 | 9.82E-07 | 6.32E-20 |
| rs7707833 | C | T | FAM169A_GCNT4 | 3 | -0.004 | -0.002 | -0.013 | 0.000 | 0.001 | 0.002 | 1.12E-16 | 1.65E-04 | 6.97E-14 |
| rs7715739 | A | G | ANKDD1B | 3 | -0.009 | -0.005 | -0.028 | 0.001 | 0.001 | 0.003 | 8.45E-22 | 3.55E-04 | 1.57E-16 |
| rs77256534 | T | C | ABHD6_DNASE1L3 | 3 | 0.008 | 0.005 | 0.028 | 0.001 | 0.002 | 0.005 | 2.20E-09 | 1.74E-02 | 8.44E-08 |
| rs77335236 | G | C | FAM102B | 3 | 0.007 | 0.004 | 0.016 | 0.001 | 0.001 | 0.004 | 6.25E-11 | 1.16E-02 | 3.53E-05 |
| rs7751601 | G | A | FRK_HS3ST5 | 3 | 0.003 | 0.001 | 0.008 | 0.001 | 0.001 | 0.002 | 3.79E-11 | 6.15E-02 | 1.14E-05 |
| rs77579349 | A | C | PACSIN1 | 3 | 0.006 | 0.004 | 0.019 | 0.001 | 0.001 | 0.003 | 1.02E-12 | 6.56E-04 | 2.01E-09 |
| rs77838597 | G | A | ANKRD31 | 3 | 0.007 | 0.005 | 0.023 | 0.001 | 0.002 | 0.004 | 9.36E-09 | 3.77E-03 | 2.57E-07 |
| rs7788515 | A | G | SP4_DNAH11 | 3 | -0.005 | -0.003 | -0.014 | 0.000 | 0.001 | 0.002 | 1.54E-28 | 1.00E-05 | 3.24E-15 |
| rs78113009 | G | T | ZNF593_CNKSR1 | 3 | -0.007 | -0.002 | -0.013 | 0.001 | 0.001 | 0.004 | 5.81E-13 | 1.00E-01 | 5.77E-04 |
| rs78155240 | T | G | DNM2 | 3 | -0.015 | -0.008 | -0.040 | 0.002 | 0.003 | 0.008 | 2.86E-13 | 5.80E-03 | 1.77E-07 |
| rs7864821 | T | C | OBP2B_ABO | 3 | -0.005 | -0.002 | -0.014 | 0.001 | 0.001 | 0.002 | 7.21E-15 | 2.03E-02 | 4.54E-09 |
| rs79351558 | G | A | DNAH11 | 3 | -0.010 | -0.006 | -0.025 | 0.001 | 0.002 | 0.005 | 3.60E-12 | 3.03E-03 | 2.22E-06 |
| rs80000866 | T | C | CEACAM16_CEACAM19 | 3 | -0.008 | -0.004 | -0.024 | 0.001 | 0.001 | 0.004 | 6.12E-14 | 3.85E-03 | 5.60E-10 |
| rs820062 | A | G | ANKS1A | 3 | 0.003 | 0.002 | 0.013 | 0.001 | 0.001 | 0.002 | 1.77E-08 | 6.00E-03 | 7.83E-09 |
| rs888789 | G | A | POC5 | 3 | -0.008 | -0.006 | -0.028 | 0.000 | 0.001 | 0.002 | 1.34E-76 | 1.80E-18 | 6.82E-60 |
| rs890928 | C | A | CEP120 | 3 | -0.003 | -0.002 | -0.011 | 0.000 | 0.001 | 0.002 | 9.80E-11 | 4.41E-04 | 7.64E-09 |
| rs892010 | C | G | YIPF2 | 3 | -0.006 | -0.004 | -0.019 | 0.001 | 0.001 | 0.004 | 8.94E-10 | 4.24E-03 | 1.93E-07 |
| rs930461 | A | T | CEACAM22P | 3 | -0.004 | -0.001 | -0.008 | 0.001 | 0.001 | 0.002 | 2.71E-12 | 4.08E-02 | 6.61E-05 |
| rs9323533 | G | A | SLC8A3_ADAM21P1 | 3 | -0.005 | -0.002 | -0.012 | 0.000 | 0.001 | 0.002 | 2.25E-28 | 2.60E-03 | 1.84E-11 |
| rs9368571 | G | C | SCAND3_LOC401242 | 3 | -0.002 | -0.002 | -0.007 | 0.001 | 0.001 | 0.002 | 4.18E-05 | 1.97E-02 | 1.54E-03 |
| rs9393691 | C | T | HIST1H2BG_HIST1H2BI | 3 | -0.003 | -0.003 | -0.014 | 0.000 | 0.001 | 0.002 | 2.46E-11 | 8.07E-06 | 8.93E-16 |
| rs962899 | G | C | HCG18_HCG17 | 3 | -0.003 | -0.003 | -0.013 | 0.001 | 0.001 | 0.002 | 3.67E-09 | 5.74E-04 | 3.56E-10 |
| rs10102164 | A | G | SOX17_RP1 | 4 | -0.006 | -0.004 | -0.017 | 0.001 | 0.001 | 0.002 | 3.54E-29 | 3.39E-07 | 1.20E-16 |
| rs10184376 | T | C | ACVR1C | 4 | 0.006 | 0.005 | 0.019 | 0.001 | 0.001 | 0.003 | 9.89E-14 | 1.28E-04 | 1.10E-09 |
| rs10402457 | C | T | SPC24 | 4 | 0.010 | 0.007 | 0.027 | 0.001 | 0.001 | 0.002 | 6.14E-63 | 1.27E-17 | 4.28E-37 |
| rs10405086 | T | C | PPP1R37_MARK4 | 4 | -0.011 | -0.007 | -0.029 | 0.001 | 0.001 | 0.003 | 4.78E-40 | 1.31E-11 | 1.23E-21 |
| rs1052883 | T | C | PLXND1 | 4 | -0.005 | -0.003 | -0.015 | 0.001 | 0.001 | 0.003 | 4.57E-16 | 6.21E-04 | 1.23E-09 |
| rs10745351 | C | T | KIAA1324 | 4 | 0.010 | 0.005 | 0.020 | 0.001 | 0.001 | 0.003 | 2.18E-38 | 1.94E-06 | 1.18E-12 |
| rs10893483 | G | A | RPUSD4_CDON | 4 | -0.005 | -0.002 | -0.013 | 0.001 | 0.001 | 0.003 | 1.21E-10 | 2.03E-02 | 8.04E-07 |
| rs11038546 | C | A | TRIM5_HBG2 | 4 | 0.003 | 0.003 | 0.010 | 0.000 | 0.001 | 0.002 | 4.89E-13 | 6.86E-05 | 1.58E-09 |
| rs11158680 | T | C | NYNRIN | 4 | -0.004 | -0.002 | -0.012 | 0.000 | 0.001 | 0.002 | 4.40E-21 | 8.17E-04 | 1.21E-11 |
| rs112214887 | G | A | CYTH1_DNAH17 | 4 | 0.003 | 0.002 | 0.006 | 0.001 | 0.001 | 0.002 | 3.48E-06 | 4.68E-02 | 1.08E-02 |
| rs112987420 | C | G | SORT1 | 4 | 0.015 | 0.008 | 0.032 | 0.002 | 0.003 | 0.007 | 2.42E-16 | 2.85E-03 | 4.36E-06 |
| rs113039407 | C | A | APOB_C2orf43 | 4 | -0.024 | -0.013 | -0.051 | 0.002 | 0.002 | 0.006 | 1.74E-54 | 8.83E-10 | 1.05E-19 |
| rs1132054 | T | C | SULT2B1 | 4 | -0.006 | -0.005 | -0.017 | 0.000 | 0.001 | 0.002 | 1.15E-30 | 1.02E-12 | 1.59E-22 |
| rs114569472 | C | G | PARP9 | 4 | -0.007 | -0.005 | -0.017 | 0.001 | 0.002 | 0.005 | 1.56E-08 | 2.52E-03 | 2.49E-04 |
| rs115049444 | C | T | SYPL2 | 4 | 0.028 | 0.015 | 0.067 | 0.002 | 0.003 | 0.007 | 1.60E-50 | 7.27E-09 | 4.86E-21 |
| rs115329105 | T | C | KLHL29_LOC645949 | 4 | 0.009 | 0.007 | 0.022 | 0.001 | 0.002 | 0.006 | 6.77E-10 | 1.69E-03 | 1.07E-04 |
| rs11538262 | T | C | PRRC2A | 4 | -0.017 | -0.011 | -0.040 | 0.002 | 0.003 | 0.008 | 4.68E-16 | 2.55E-04 | 5.68E-07 |
| rs11554765 | T | C | DHX38 | 4 | -0.013 | -0.008 | -0.038 | 0.002 | 0.003 | 0.008 | 8.76E-10 | 8.73E-03 | 3.60E-06 |
| rs115817702 | T | C | PRKCSH | 4 | 0.007 | 0.005 | 0.019 | 0.001 | 0.002 | 0.005 | 6.33E-09 | 4.39E-03 | 4.86E-05 |
| rs116042783 | T | C | SLC17A4 | 4 | 0.015 | 0.011 | 0.056 | 0.002 | 0.003 | 0.007 | 4.63E-15 | 3.61E-05 | 6.94E-15 |
| rs116157399 | A | G | APOB_LOC645949 | 4 | -0.026 | -0.013 | -0.068 | 0.002 | 0.002 | 0.007 | 4.61E-48 | 1.14E-07 | 1.67E-24 |
| rs116276872 | T | C | AMIGO1_GPR61 | 4 | 0.012 | 0.006 | 0.025 | 0.002 | 0.002 | 0.006 | 1.33E-13 | 1.28E-02 | 3.69E-05 |
| rs116416309 | C | T | KIAA1324 | 4 | 0.010 | 0.006 | 0.019 | 0.001 | 0.001 | 0.003 | 1.39E-44 | 2.85E-08 | 1.53E-12 |
| rs11669683 | A | G | MYPOP_NANOS2 | 4 | -0.003 | -0.002 | -0.008 | 0.000 | 0.001 | 0.002 | 1.15E-08 | 2.14E-03 | 1.93E-06 |
| rs11683623 | G | A | PPM1B_LRPPRC | 4 | 0.003 | 0.002 | 0.010 | 0.001 | 0.001 | 0.002 | 5.12E-08 | 1.24E-02 | 1.18E-05 |
| rs116967764 | A | G | PVRL2 | 4 | -0.012 | -0.006 | -0.033 | 0.002 | 0.003 | 0.007 | 4.18E-10 | 3.24E-02 | 2.57E-06 |
| rs11772223 | G | A | NUDCD3_CAMK2B | 4 | -0.004 | -0.003 | -0.014 | 0.001 | 0.001 | 0.002 | 7.89E-13 | 3.84E-05 | 1.10E-10 |
| rs117983270 | C | T | FADS2 | 4 | 0.009 | 0.003 | 0.016 | 0.001 | 0.002 | 0.005 | 9.12E-13 | 4.67E-02 | 3.62E-04 |
| rs11800265 | A | G | PCSK9 | 4 | 0.010 | 0.007 | 0.027 | 0.001 | 0.002 | 0.004 | 6.43E-18 | 2.49E-05 | 2.36E-10 |
| rs118146420 | G | A | LOC157273_PPP1R3B | 4 | 0.006 | 0.005 | 0.019 | 0.001 | 0.002 | 0.004 | 2.32E-07 | 1.87E-03 | 2.74E-05 |
| rs1182864 | A | G | KCNK5_KCNK17 | 4 | 0.003 | 0.002 | 0.007 | 0.000 | 0.001 | 0.002 | 6.43E-12 | 6.78E-03 | 2.73E-04 |
| rs11857386 | C | G | LIPC_ALDH1A2 | 4 | 0.003 | 0.002 | 0.009 | 0.000 | 0.001 | 0.002 | 4.95E-12 | 1.10E-04 | 5.82E-08 |
| rs123698 | C | G | PTBP1 | 4 | -0.003 | -0.002 | -0.006 | 0.000 | 0.001 | 0.002 | 4.06E-10 | 1.31E-02 | 4.75E-04 |
| rs12601655 | G | A | PGS1_SOCS3 | 4 | 0.003 | 0.003 | 0.012 | 0.001 | 0.001 | 0.002 | 2.12E-09 | 1.40E-03 | 1.76E-08 |
| rs1264518 | T | C | RPP21_HLA-E | 4 | -0.003 | -0.003 | -0.013 | 0.001 | 0.001 | 0.002 | 6.50E-07 | 6.02E-04 | 3.37E-08 |
| rs12669528 | G | C | SP4 | 4 | -0.007 | -0.004 | -0.019 | 0.000 | 0.001 | 0.002 | 9.26E-46 | 1.62E-09 | 3.01E-24 |
| rs12728258 | C | T | MAN1C1 | 4 | 0.003 | 0.003 | 0.010 | 0.001 | 0.001 | 0.002 | 1.11E-09 | 1.45E-03 | 1.48E-06 |
| rs12974306 | T | G | DNM2 | 4 | -0.004 | -0.002 | -0.010 | 0.000 | 0.001 | 0.002 | 4.11E-17 | 1.23E-03 | 2.79E-09 |
| rs13268 | G | A | FBLN1 | 4 | 0.008 | 0.005 | 0.026 | 0.001 | 0.002 | 0.006 | 3.73E-08 | 2.48E-02 | 4.07E-06 |
| rs13408439 | G | A | KLHL29_LOC645949 | 4 | 0.010 | 0.006 | 0.023 | 0.001 | 0.002 | 0.005 | 1.40E-15 | 9.59E-04 | 2.92E-06 |
| rs140584594 | G | A | GSTM1_GSTM2 | 4 | 0.007 | 0.003 | 0.012 | 0.001 | 0.001 | 0.002 | 1.66E-41 | 5.16E-06 | 2.60E-09 |
| rs14234 | G | A | FAM136A | 4 | -0.002 | -0.002 | -0.007 | 0.000 | 0.001 | 0.002 | 3.60E-07 | 1.82E-02 | 9.08E-05 |
| rs144923024 | A | G | RAPH1_CD28 | 4 | 0.007 | 0.006 | 0.027 | 0.001 | 0.002 | 0.005 | 4.77E-08 | 1.77E-03 | 7.84E-08 |
| rs1661052 | A | G | SLC22A18 | 4 | -0.004 | -0.003 | -0.013 | 0.001 | 0.001 | 0.003 | 2.96E-06 | 7.50E-03 | 1.63E-05 |
| rs16895223 | G | C | MOG | 4 | -0.005 | -0.005 | -0.016 | 0.001 | 0.001 | 0.003 | 1.63E-11 | 2.13E-06 | 1.44E-10 |
| rs16982241 | A | G | FUT2 | 4 | 0.005 | 0.004 | 0.015 | 0.001 | 0.001 | 0.003 | 7.83E-14 | 1.54E-04 | 9.86E-09 |
| rs17029597 | C | A | CMTM6_CMTM7 | 4 | 0.008 | 0.006 | 0.032 | 0.001 | 0.001 | 0.003 | 1.37E-20 | 3.17E-06 | 4.46E-20 |
| rs17031488 | T | C | DYNC2LI1_PLEKHH2 | 4 | -0.006 | -0.004 | -0.016 | 0.001 | 0.001 | 0.002 | 4.89E-19 | 1.85E-06 | 1.65E-11 |
| rs17036085 | G | A | PSRC1_MYBPHL | 4 | 0.017 | 0.011 | 0.045 | 0.002 | 0.003 | 0.007 | 1.97E-18 | 1.14E-04 | 1.74E-09 |
| rs17206700 | C | T | PLEKHA4 | 4 | -0.005 | -0.003 | -0.012 | 0.001 | 0.001 | 0.002 | 9.90E-15 | 7.99E-05 | 1.87E-07 |
| rs17357122 | A | G | CKM | 4 | -0.014 | -0.006 | -0.040 | 0.002 | 0.003 | 0.009 | 1.17E-09 | 4.27E-02 | 2.49E-06 |
| rs17519958 | A | C | IRF2BP2_LINC00184 | 4 | 0.004 | 0.002 | 0.010 | 0.000 | 0.001 | 0.002 | 4.67E-13 | 1.18E-02 | 1.43E-07 |
| rs17641881 | C | T | KIAA1324 | 4 | -0.007 | -0.005 | -0.019 | 0.001 | 0.002 | 0.004 | 6.14E-10 | 2.43E-03 | 2.79E-05 |
| rs180728024 | A | G | HYDIN | 4 | 0.005 | 0.004 | 0.019 | 0.001 | 0.001 | 0.004 | 9.83E-07 | 4.68E-03 | 3.14E-06 |
| rs187183066 | G | A | PVRL2 | 4 | -0.014 | -0.007 | -0.037 | 0.002 | 0.003 | 0.008 | 7.18E-11 | 9.79E-03 | 1.89E-06 |
| rs206322 | G | A | N4BP2L1 | 4 | 0.005 | 0.004 | 0.013 | 0.001 | 0.001 | 0.002 | 2.46E-20 | 4.20E-06 | 7.37E-10 |
| rs217180 | T | C | PMFBP1 | 4 | -0.006 | -0.003 | -0.016 | 0.001 | 0.001 | 0.003 | 4.04E-16 | 1.07E-02 | 2.60E-08 |
| rs2289055 | G | A | DDX56 | 4 | -0.006 | -0.005 | -0.019 | 0.001 | 0.001 | 0.003 | 1.25E-13 | 1.30E-05 | 1.61E-10 |
| rs2358956 | C | T | TAF13 | 4 | -0.011 | -0.004 | -0.021 | 0.001 | 0.001 | 0.004 | 1.17E-30 | 7.75E-04 | 3.19E-09 |
| rs240780 | C | G | ASCC3 | 4 | 0.002 | 0.001 | 0.003 | 0.000 | 0.001 | 0.002 | 1.12E-07 | 2.85E-01 | 4.95E-02 |
| rs2548957 | A | G | FGF21_BCAT2 | 4 | 0.004 | 0.003 | 0.012 | 0.000 | 0.001 | 0.002 | 1.48E-14 | 4.99E-05 | 5.56E-12 |
| rs2802950 | G | A | TOMM20_LOC100506810 | 4 | 0.003 | 0.002 | 0.009 | 0.000 | 0.001 | 0.002 | 7.55E-12 | 4.58E-03 | 2.89E-07 |
| rs2836908 | T | C | PSMG1_ETS2 | 4 | -0.003 | -0.002 | -0.006 | 0.000 | 0.001 | 0.002 | 2.84E-10 | 9.42E-03 | 1.16E-03 |
| rs28399443 | A | G | CYP2A6 | 4 | 0.009 | 0.005 | 0.026 | 0.002 | 0.002 | 0.006 | 2.22E-07 | 3.87E-02 | 4.02E-05 |
| rs3094065 | C | T | HCG18_HCG17 | 4 | -0.005 | -0.004 | -0.020 | 0.001 | 0.001 | 0.003 | 3.36E-13 | 2.76E-05 | 3.26E-14 |
| rs34114202 | A | G | ZSCAN18 | 4 | 0.004 | 0.002 | 0.012 | 0.001 | 0.001 | 0.002 | 2.50E-10 | 5.38E-03 | 4.81E-08 |
| rs34978331 | C | T | PVRL2_BCAM | 4 | 0.009 | 0.005 | 0.022 | 0.001 | 0.001 | 0.002 | 4.19E-60 | 3.16E-09 | 3.55E-26 |
| rs3738621 | G | A | TOMM20_LOC100506810 | 4 | 0.004 | 0.003 | 0.013 | 0.001 | 0.001 | 0.002 | 2.58E-15 | 9.06E-06 | 3.29E-10 |
| rs3742520 | C | A | KHNYN | 4 | -0.003 | -0.001 | -0.008 | 0.000 | 0.001 | 0.002 | 2.47E-14 | 3.25E-02 | 5.62E-06 |
| rs3748433 | A | G | CEP250 | 4 | 0.005 | 0.004 | 0.017 | 0.001 | 0.001 | 0.003 | 3.35E-10 | 2.77E-04 | 5.52E-08 |
| rs3816873 | C | T | MTTP | 4 | 0.003 | 0.002 | 0.008 | 0.001 | 0.001 | 0.002 | 4.06E-11 | 1.00E-02 | 8.40E-05 |
| rs3848726 | T | G | SLC12A5 | 4 | 0.003 | 0.002 | 0.008 | 0.000 | 0.001 | 0.002 | 3.19E-10 | 3.79E-03 | 3.27E-06 |
| rs4073455 | G | T | PLEC_EPPK1 | 4 | -0.003 | -0.001 | -0.007 | 0.000 | 0.001 | 0.002 | 6.83E-14 | 8.43E-02 | 2.02E-04 |
| rs409331 | G | T | APOB_LOC645949 | 4 | -0.011 | -0.006 | -0.024 | 0.001 | 0.001 | 0.004 | 1.16E-31 | 2.32E-05 | 1.37E-11 |
| rs41278174 | A | G | ABCC6 | 4 | -0.008 | -0.004 | -0.021 | 0.001 | 0.002 | 0.005 | 3.01E-08 | 3.79E-02 | 9.28E-05 |
| rs41289512 | G | C | PVRL2 | 4 | -0.052 | -0.026 | -0.115 | 0.001 | 0.002 | 0.004 | 0.00E+00 | 1.67E-59 | 4.09E-148 |
| rs4310184 | C | T | SLC7A13_WWP1 | 4 | 0.003 | 0.002 | 0.010 | 0.001 | 0.001 | 0.002 | 5.80E-09 | 6.81E-03 | 3.01E-07 |
| rs4775041 | C | G | ALDH1A2_LIPC | 4 | -0.007 | -0.003 | -0.015 | 0.000 | 0.001 | 0.002 | 1.93E-41 | 3.50E-05 | 2.89E-15 |
| rs4804149 | C | T | KANK2 | 4 | -0.006 | -0.003 | -0.014 | 0.001 | 0.001 | 0.002 | 1.75E-30 | 5.42E-06 | 3.78E-12 |
| rs545587 | C | A | HSD17B14 | 4 | 0.003 | 0.002 | 0.008 | 0.001 | 0.001 | 0.002 | 3.32E-09 | 8.48E-04 | 1.53E-05 |
| rs561035 | T | C | PTEN_RNLS | 4 | 0.008 | 0.005 | 0.020 | 0.001 | 0.002 | 0.005 | 2.13E-10 | 5.33E-03 | 8.11E-05 |
| rs6029228 | G | T | MAFB_LOC339568 | 4 | 0.005 | 0.004 | 0.016 | 0.000 | 0.001 | 0.002 | 6.61E-24 | 1.32E-10 | 2.91E-18 |
| rs603424 | A | G | PKD2L1 | 4 | -0.005 | -0.002 | -0.010 | 0.001 | 0.001 | 0.002 | 9.90E-18 | 3.53E-02 | 4.34E-06 |
| rs6115094 | G | A | PYGB_ENTPD6 | 4 | -0.004 | -0.002 | -0.011 | 0.000 | 0.001 | 0.002 | 2.19E-15 | 1.00E-04 | 5.17E-10 |
| rs6124298 | A | G | LOC100128988_TOP1 | 4 | -0.005 | -0.003 | -0.014 | 0.000 | 0.001 | 0.002 | 3.98E-24 | 1.04E-04 | 8.41E-14 |
| rs61754230 | T | C | RAB21 | 4 | -0.013 | -0.008 | -0.036 | 0.002 | 0.002 | 0.006 | 4.91E-14 | 3.06E-04 | 1.11E-08 |
| rs61756355 | A | C | TRIM6_TRIM34 | 4 | 0.008 | 0.004 | 0.017 | 0.001 | 0.002 | 0.005 | 6.77E-09 | 6.24E-02 | 7.76E-04 |
| rs62074057 | G | A | TBX21_TBKBP1 | 4 | 0.005 | 0.003 | 0.015 | 0.001 | 0.001 | 0.003 | 1.31E-14 | 1.97E-03 | 8.98E-09 |
| rs621167 | C | A | LINC00184_LOC100506810 | 4 | -0.003 | -0.003 | -0.009 | 0.000 | 0.001 | 0.002 | 8.29E-09 | 1.21E-04 | 1.73E-07 |
| rs62130341 | T | C | SEC1P | 4 | 0.005 | 0.003 | 0.014 | 0.001 | 0.001 | 0.003 | 1.42E-12 | 9.40E-04 | 4.73E-07 |
| rs6603883 | G | A | EPHA2_ARHGEF19 | 4 | -0.004 | -0.003 | -0.010 | 0.000 | 0.001 | 0.002 | 2.42E-19 | 1.07E-04 | 7.04E-08 |
| rs662138 | G | C | SLC22A1 | 4 | -0.011 | -0.008 | -0.033 | 0.001 | 0.001 | 0.002 | 1.47E-84 | 2.23E-20 | 9.73E-50 |
| rs6650591 | T | C | FOXK2 | 4 | 0.003 | 0.002 | 0.006 | 0.000 | 0.001 | 0.002 | 6.75E-10 | 1.29E-02 | 1.08E-03 |
| rs66720010 | G | A | MLYCD_OSGIN1 | 4 | 0.005 | 0.003 | 0.013 | 0.000 | 0.001 | 0.002 | 3.54E-27 | 2.54E-07 | 5.65E-13 |
| rs66777321 | A | G | NUFIP2_MIR4523 | 4 | 0.005 | 0.004 | 0.016 | 0.001 | 0.001 | 0.002 | 4.65E-15 | 4.14E-05 | 4.00E-11 |
| rs684959 | T | C | BCAT2_FGF21 | 4 | -0.003 | -0.002 | -0.007 | 0.000 | 0.001 | 0.002 | 6.06E-09 | 2.39E-03 | 1.11E-04 |
| rs6942171 | T | C | FIG4 | 4 | 0.003 | 0.002 | 0.007 | 0.000 | 0.001 | 0.002 | 1.30E-08 | 9.55E-04 | 7.64E-05 |
| rs7249753 | C | T | LDLR_SMARCA4 | 4 | -0.015 | -0.009 | -0.040 | 0.001 | 0.001 | 0.004 | 1.15E-50 | 2.41E-11 | 9.56E-28 |
| rs72626214 | T | C | CEACAM16_CEACAM19 | 4 | 0.006 | 0.004 | 0.017 | 0.001 | 0.001 | 0.003 | 2.51E-17 | 7.59E-05 | 1.44E-09 |
| rs72654445 | A | G | APOE_APOC1 | 4 | -0.019 | -0.010 | -0.057 | 0.002 | 0.003 | 0.008 | 3.54E-17 | 5.81E-04 | 1.83E-12 |
| rs72705211 | A | G | AMPD2_GSTM4 | 4 | -0.007 | -0.003 | -0.013 | 0.001 | 0.002 | 0.004 | 1.82E-10 | 1.05E-01 | 2.30E-03 |
| rs72768400 | T | G | POC5_SV2C | 4 | 0.006 | 0.005 | 0.017 | 0.001 | 0.001 | 0.004 | 1.54E-07 | 1.78E-03 | 2.97E-05 |
| rs72782175 | C | T | APOB_LOC645949 | 4 | 0.030 | 0.020 | 0.078 | 0.002 | 0.003 | 0.008 | 1.49E-47 | 1.41E-11 | 2.45E-23 |
| rs72807535 | G | A | EHBP1_TMEM17 | 4 | 0.005 | 0.004 | 0.016 | 0.001 | 0.001 | 0.003 | 1.88E-14 | 2.61E-04 | 1.22E-09 |
| rs72861370 | A | C | MAP2K6 | 4 | -0.007 | -0.005 | -0.020 | 0.001 | 0.001 | 0.003 | 3.77E-19 | 7.17E-05 | 9.05E-11 |
| rs72905574 | T | C | TMEM61_DHCR24 | 4 | 0.007 | 0.005 | 0.020 | 0.001 | 0.001 | 0.003 | 4.31E-17 | 2.23E-06 | 2.42E-11 |
| rs72976313 | A | G | MAP3K19 | 4 | -0.005 | -0.003 | -0.013 | 0.001 | 0.001 | 0.003 | 1.33E-08 | 1.31E-02 | 7.51E-05 |
| rs73118985 | C | T | MAFB_LOC339568 | 4 | 0.007 | 0.006 | 0.021 | 0.001 | 0.002 | 0.004 | 2.05E-10 | 8.99E-05 | 2.44E-06 |
| rs73611229 | T | G | LOC100128988_TOP1 | 4 | -0.004 | -0.003 | -0.009 | 0.001 | 0.001 | 0.002 | 1.63E-10 | 2.00E-03 | 1.10E-04 |
| rs7415034 | T | C | RUNX3_SYF2 | 4 | 0.006 | 0.003 | 0.014 | 0.001 | 0.001 | 0.003 | 1.83E-13 | 2.20E-03 | 1.61E-06 |
| rs74361782 | T | C | MYO15A_DRG2 | 4 | -0.009 | -0.005 | -0.023 | 0.001 | 0.002 | 0.005 | 1.18E-10 | 2.17E-02 | 2.03E-05 |
| rs75237799 | T | C | CELSR2 | 4 | 0.035 | 0.019 | 0.082 | 0.002 | 0.003 | 0.008 | 1.05E-66 | 4.47E-11 | 2.51E-26 |
| rs7539980 | C | A | GSTM5 | 4 | -0.006 | -0.003 | -0.014 | 0.001 | 0.001 | 0.003 | 2.71E-12 | 3.60E-03 | 3.77E-06 |
| rs75679231 | C | A | GDF7_HS1BP3 | 4 | -0.014 | -0.009 | -0.037 | 0.001 | 0.002 | 0.005 | 1.10E-22 | 3.45E-06 | 1.16E-11 |
| rs7569328 | T | C | APOB_C2orf43 | 4 | 0.016 | 0.009 | 0.039 | 0.002 | 0.002 | 0.007 | 6.36E-21 | 3.43E-04 | 3.50E-09 |
| rs7572443 | C | A | SULT1C2P1_SULT1C4 | 4 | -0.005 | -0.004 | -0.014 | 0.001 | 0.001 | 0.002 | 1.46E-12 | 1.10E-04 | 6.79E-09 |
| rs76419583 | C | T | CEACAM16_BCL3 | 4 | -0.007 | -0.003 | -0.015 | 0.001 | 0.001 | 0.002 | 2.22E-25 | 4.03E-04 | 1.25E-09 |
| rs77011887 | T | C | USP24 | 4 | -0.009 | -0.006 | -0.028 | 0.002 | 0.002 | 0.006 | 2.18E-07 | 1.14E-02 | 1.08E-05 |
| rs77380569 | A | G | CLPTM1 | 4 | -0.022 | -0.012 | -0.046 | 0.003 | 0.004 | 0.010 | 3.93E-16 | 8.62E-04 | 1.75E-06 |
| rs77600363 | T | G | NPAS4 | 4 | 0.003 | 0.003 | 0.011 | 0.001 | 0.001 | 0.002 | 9.73E-10 | 2.28E-05 | 2.14E-07 |
| rs78635447 | A | G | THADA | 4 | -0.007 | -0.007 | -0.024 | 0.001 | 0.002 | 0.004 | 2.42E-09 | 2.47E-05 | 1.60E-08 |
| rs79167099 | A | C | PUM2 | 4 | -0.010 | -0.005 | -0.027 | 0.002 | 0.003 | 0.007 | 5.41E-08 | 6.48E-02 | 8.35E-05 |
| rs7961554 | C | A | HNF1A-AS1_SPPL3 | 4 | 0.003 | 0.002 | 0.009 | 0.000 | 0.001 | 0.002 | 2.24E-09 | 9.70E-03 | 1.32E-06 |
| rs796291 | A | G | PPARG_TSEN2 | 4 | 0.003 | 0.002 | 0.010 | 0.000 | 0.001 | 0.002 | 3.25E-10 | 8.99E-03 | 2.92E-08 |
| rs80257887 | A | G | CEACAM20 | 4 | -0.010 | -0.007 | -0.025 | 0.001 | 0.002 | 0.005 | 1.35E-14 | 1.73E-04 | 2.97E-07 |
| rs80315641 | A | G | COL4A3BP | 4 | 0.009 | 0.006 | 0.030 | 0.001 | 0.002 | 0.005 | 2.99E-14 | 6.36E-04 | 1.64E-10 |
| rs8051191 | G | C | MIR138-2_SLC12A3 | 4 | 0.005 | 0.002 | 0.008 | 0.001 | 0.001 | 0.002 | 1.61E-14 | 6.12E-03 | 1.99E-04 |
| rs896738 | G | A | HS1BP3_RHOB | 4 | 0.013 | 0.006 | 0.027 | 0.001 | 0.002 | 0.005 | 8.62E-22 | 1.96E-03 | 3.40E-08 |
| rs909334 | A | C | STMN3_LOC100505771 | 4 | -0.003 | -0.002 | -0.010 | 0.001 | 0.001 | 0.002 | 4.81E-08 | 3.46E-03 | 3.96E-07 |
| rs934287 | G | A | ICA1L | 4 | -0.005 | -0.004 | -0.016 | 0.001 | 0.001 | 0.002 | 1.10E-19 | 1.07E-05 | 1.59E-12 |
| rs9366627 | T | C | SCGN | 4 | 0.003 | 0.002 | 0.010 | 0.001 | 0.001 | 0.002 | 1.91E-08 | 1.28E-02 | 3.85E-07 |
| rs9376090 | C | T | HBS1L_MYB | 4 | 0.005 | 0.004 | 0.015 | 0.001 | 0.001 | 0.002 | 5.56E-25 | 3.42E-07 | 1.52E-14 |
| rs9468007 | T | C | MIR3143_PRSS16 | 4 | -0.007 | -0.004 | -0.020 | 0.001 | 0.002 | 0.005 | 2.97E-08 | 1.81E-02 | 1.29E-05 |
| rs9470298 | C | T | ADTRP_HIVEP1 | 4 | 0.006 | 0.004 | 0.014 | 0.001 | 0.001 | 0.003 | 1.77E-14 | 1.29E-04 | 1.22E-06 |
| rs9819988 | C | T | PPP2R3A_EPHB1 | 4 | -0.003 | -0.002 | -0.010 | 0.000 | 0.001 | 0.002 | 1.58E-11 | 3.58E-04 | 6.20E-08 |
| rs985586 | A | G | SDC4 | 4 | 0.004 | 0.002 | 0.010 | 0.001 | 0.001 | 0.002 | 1.51E-11 | 9.41E-04 | 9.10E-07 |
| rs10175646 | C | T | APOB_C2orf43 | 5 | 0.017 | 0.011 | 0.042 | 0.001 | 0.002 | 0.005 | 5.51E-42 | 3.02E-10 | 7.32E-19 |
| rs10484563 | G | A | HLA-DQA2_HLA-DQB1 | 5 | -0.006 | -0.005 | -0.018 | 0.001 | 0.001 | 0.004 | 5.91E-10 | 3.65E-05 | 2.60E-07 |
| rs10491298 | G | A | SPRY4_FGF1 | 5 | -0.004 | -0.003 | -0.013 | 0.001 | 0.001 | 0.003 | 9.35E-08 | 1.59E-03 | 1.97E-06 |
| rs10832948 | A | G | SPTY2D1_SPTY2D1-AS1 | 5 | -0.003 | -0.002 | -0.008 | 0.000 | 0.001 | 0.002 | 6.31E-14 | 2.44E-03 | 7.60E-06 |
| rs10846490 | A | G | CDK2AP1 | 5 | -0.005 | -0.004 | -0.014 | 0.001 | 0.001 | 0.004 | 8.74E-07 | 9.27E-03 | 2.18E-04 |
| rs10885998 | A | G | PNLIPRP2_C10orf82 | 5 | -0.003 | -0.003 | -0.010 | 0.000 | 0.001 | 0.002 | 1.88E-08 | 1.06E-05 | 2.19E-07 |
| rs11024685 | C | T | TSG101 | 5 | 0.004 | 0.003 | 0.011 | 0.001 | 0.001 | 0.002 | 1.61E-13 | 1.35E-03 | 4.94E-06 |
| rs111577916 | T | G | RPUSD4_CDON | 5 | -0.020 | -0.013 | -0.055 | 0.002 | 0.002 | 0.006 | 3.30E-36 | 8.00E-09 | 3.13E-20 |
| rs111751551 | A | G | PRPF38B | 5 | 0.016 | 0.011 | 0.038 | 0.002 | 0.003 | 0.008 | 4.13E-13 | 3.12E-04 | 7.57E-06 |
| rs114168203 | T | C | SORT1 | 5 | -0.011 | -0.007 | -0.024 | 0.002 | 0.002 | 0.006 | 8.74E-14 | 7.07E-04 | 2.70E-05 |
| rs115904025 | A | G | CLCC1_AKNAD1 | 5 | 0.011 | 0.010 | 0.032 | 0.002 | 0.002 | 0.006 | 2.48E-12 | 1.03E-05 | 8.11E-08 |
| rs116009799 | T | C | LINC00243_DDR1 | 5 | -0.012 | -0.010 | -0.033 | 0.002 | 0.003 | 0.008 | 4.83E-09 | 1.02E-03 | 2.44E-05 |
| rs116032419 | C | A | ACAD11_NPHP3 | 5 | 0.010 | 0.005 | 0.026 | 0.002 | 0.003 | 0.007 | 3.28E-08 | 8.81E-02 | 3.16E-04 |
| rs11626364 | C | T | ELMSAN1 | 5 | 0.004 | 0.003 | 0.010 | 0.001 | 0.001 | 0.002 | 7.92E-11 | 4.25E-03 | 2.09E-05 |
| rs11629005 | C | T | NYNRIN_NFATC4 | 5 | -0.003 | -0.002 | -0.009 | 0.000 | 0.001 | 0.002 | 1.13E-09 | 6.70E-04 | 4.12E-07 |
| rs11632618 | A | G | LIPC_ALDH1A2 | 5 | -0.006 | -0.004 | -0.016 | 0.001 | 0.001 | 0.003 | 7.95E-13 | 1.01E-03 | 1.72E-06 |
| rs11651753 | T | C | PNPO_PRR15L | 5 | 0.004 | 0.002 | 0.009 | 0.000 | 0.001 | 0.002 | 1.52E-16 | 1.63E-04 | 1.16E-07 |
| rs11655536 | G | A | MYL4_ITGB3 | 5 | 0.004 | 0.002 | 0.009 | 0.001 | 0.001 | 0.002 | 6.21E-13 | 3.44E-03 | 1.39E-06 |
| rs116661368 | T | C | LOC100507562_APOB | 5 | 0.016 | 0.009 | 0.032 | 0.002 | 0.003 | 0.008 | 5.67E-14 | 1.68E-03 | 3.81E-05 |
| rs11673516 | G | T | OPA3 | 5 | -0.003 | -0.002 | -0.010 | 0.000 | 0.001 | 0.002 | 5.62E-08 | 3.49E-04 | 2.44E-08 |
| rs117743564 | A | C | ILF3 | 5 | -0.008 | -0.005 | -0.022 | 0.001 | 0.002 | 0.005 | 6.25E-08 | 7.71E-03 | 3.23E-05 |
| rs117780561 | A | G | RILPL1 | 5 | 0.010 | 0.004 | 0.017 | 0.002 | 0.002 | 0.006 | 3.71E-11 | 6.06E-02 | 3.51E-03 |
| rs118014788 | T | G | ACACB | 5 | -0.009 | -0.009 | -0.025 | 0.001 | 0.002 | 0.005 | 2.43E-10 | 1.21E-05 | 2.42E-06 |
| rs11905831 | G | A | PCMTD2 | 5 | -0.004 | -0.004 | -0.015 | 0.001 | 0.001 | 0.002 | 6.30E-15 | 3.48E-08 | 6.75E-12 |
| rs12162782 | G | T | PPP6R2 | 5 | -0.004 | -0.003 | -0.010 | 0.000 | 0.001 | 0.002 | 9.14E-16 | 7.81E-06 | 3.32E-08 |
| rs1235378 | G | A | CEACAM20 | 5 | 0.004 | 0.002 | 0.008 | 0.001 | 0.001 | 0.002 | 2.49E-12 | 2.40E-04 | 1.44E-05 |
| rs12433338 | C | T | NFATC4_RIPK3 | 5 | 0.004 | 0.003 | 0.014 | 0.001 | 0.001 | 0.002 | 1.60E-14 | 2.09E-05 | 1.37E-10 |
| rs12528797 | G | A | C6orf10_HCG23 | 5 | -0.010 | -0.010 | -0.031 | 0.001 | 0.001 | 0.003 | 1.64E-41 | 1.45E-25 | 1.40E-30 |
| rs12783584 | C | T | PNLIPRP2 | 5 | -0.003 | -0.002 | -0.008 | 0.000 | 0.001 | 0.002 | 3.29E-10 | 8.94E-04 | 2.76E-06 |
| rs12968116 | T | C | ATP8B1_LOC100505549 | 5 | -0.004 | -0.005 | -0.013 | 0.001 | 0.001 | 0.003 | 5.38E-09 | 3.25E-06 | 3.19E-07 |
| rs13406645 | T | G | LOC100652824 | 5 | 0.004 | 0.003 | 0.011 | 0.001 | 0.001 | 0.002 | 1.02E-09 | 1.20E-04 | 1.86E-06 |
| rs1362965 | T | C | METTL7A | 5 | -0.003 | -0.002 | -0.009 | 0.000 | 0.001 | 0.002 | 1.11E-09 | 6.88E-04 | 2.05E-07 |
| rs1379263 | A | G | ZNF366_TNPO1 | 5 | 0.005 | 0.004 | 0.016 | 0.001 | 0.001 | 0.003 | 2.85E-09 | 5.92E-04 | 8.73E-08 |
| rs138914864 | T | C | PVRL2 | 5 | -0.078 | -0.064 | -0.194 | 0.005 | 0.006 | 0.018 | 2.29E-56 | 3.14E-23 | 8.03E-28 |
| rs1395613 | C | A | FAM110B_UBXN2B | 5 | 0.008 | 0.008 | 0.028 | 0.001 | 0.002 | 0.005 | 1.87E-11 | 9.38E-06 | 3.05E-09 |
| rs141828689 | T | C | ABCG5 | 5 | -0.042 | -0.034 | -0.108 | 0.006 | 0.009 | 0.023 | 2.93E-11 | 9.34E-05 | 3.92E-06 |
| rs1464603 | A | G | NR1I2 | 5 | 0.002 | 0.003 | 0.010 | 0.000 | 0.001 | 0.002 | 1.03E-05 | 2.06E-04 | 6.52E-08 |
| rs1473147 | C | T | ABCA6_ABCA9 | 5 | 0.006 | 0.004 | 0.017 | 0.001 | 0.001 | 0.002 | 7.98E-28 | 1.68E-09 | 5.35E-18 |
| rs1491942 | G | C | LRRK2 | 5 | -0.003 | -0.002 | -0.009 | 0.001 | 0.001 | 0.002 | 6.29E-07 | 2.68E-03 | 1.35E-05 |
| rs1539680 | C | G | CACNB2 | 5 | -0.003 | -0.003 | -0.011 | 0.001 | 0.001 | 0.002 | 3.64E-09 | 1.22E-05 | 1.36E-07 |
| rs157583 | T | G | TOMM40 | 5 | -0.047 | -0.032 | -0.099 | 0.004 | 0.006 | 0.016 | 2.37E-25 | 1.01E-07 | 1.61E-09 |
| rs16884048 | C | A | GCLC_KLHL31 | 5 | -0.005 | -0.003 | -0.010 | 0.001 | 0.001 | 0.002 | 1.23E-14 | 1.05E-04 | 1.93E-05 |
| rs16923452 | G | A | UBXN2B | 5 | 0.004 | 0.004 | 0.013 | 0.000 | 0.001 | 0.002 | 3.75E-16 | 6.31E-07 | 3.47E-12 |
| rs17047429 | C | T | DPP10_DDX18 | 5 | -0.009 | -0.007 | -0.027 | 0.001 | 0.001 | 0.004 | 1.72E-17 | 1.04E-06 | 1.06E-11 |
| rs17050272 | A | G | GLI2_LOC84931 | 5 | 0.007 | 0.005 | 0.018 | 0.000 | 0.001 | 0.002 | 1.28E-48 | 1.90E-13 | 5.24E-25 |
| rs17103672 | G | A | NYNRIN | 5 | 0.006 | 0.004 | 0.017 | 0.001 | 0.001 | 0.004 | 4.68E-11 | 4.24E-03 | 6.40E-06 |
| rs1714013 | A | G | IGFBP7 | 5 | 0.003 | 0.002 | 0.007 | 0.001 | 0.001 | 0.002 | 2.16E-09 | 5.69E-03 | 5.30E-04 |
| rs17184798 | C | T | DNAJC13_ACPP | 5 | 0.007 | 0.005 | 0.018 | 0.001 | 0.001 | 0.003 | 7.45E-21 | 3.45E-05 | 1.79E-09 |
| rs17201964 | G | A | SP4_RPL23P8 | 5 | 0.004 | 0.004 | 0.011 | 0.001 | 0.001 | 0.002 | 7.74E-12 | 1.74E-05 | 3.71E-07 |
| rs17399768 | C | A | APOB_LOC645949 | 5 | 0.046 | 0.025 | 0.102 | 0.006 | 0.008 | 0.022 | 5.99E-16 | 1.74E-03 | 3.21E-06 |
| rs1892496 | G | A | TRIM63 | 5 | -0.003 | -0.002 | -0.006 | 0.000 | 0.001 | 0.002 | 8.60E-11 | 5.85E-04 | 1.46E-03 |
| rs1902023 | C | A | UGT2B15 | 5 | -0.003 | -0.003 | -0.009 | 0.000 | 0.001 | 0.002 | 7.92E-09 | 1.18E-06 | 2.92E-07 |
| rs1997833 | C | T | TOP1 | 5 | -0.008 | -0.005 | -0.019 | 0.001 | 0.001 | 0.002 | 2.00E-53 | 6.35E-13 | 5.98E-22 |
| rs201478913 | T | C | PPP1R13L | 5 | -0.045 | -0.040 | -0.135 | 0.007 | 0.009 | 0.026 | 3.47E-10 | 2.23E-05 | 1.88E-07 |
| rs222859 | A | C | YBX2 | 5 | 0.003 | 0.003 | 0.007 | 0.001 | 0.001 | 0.002 | 1.84E-08 | 6.38E-04 | 9.11E-04 |
| rs2234694 | C | A | SOD1 | 5 | -0.008 | -0.006 | -0.016 | 0.001 | 0.002 | 0.004 | 7.82E-13 | 4.63E-04 | 1.87E-04 |
| rs2285888 | C | T | CYP4F12 | 5 | -0.003 | -0.002 | -0.011 | 0.000 | 0.001 | 0.002 | 2.20E-09 | 2.05E-04 | 1.19E-09 |
| rs235999 | T | C | FTSJD1_HYDIN | 5 | -0.004 | -0.002 | -0.008 | 0.001 | 0.001 | 0.002 | 7.27E-12 | 1.76E-02 | 2.37E-04 |
| rs2537855 | G | A | THOP1 | 5 | -0.003 | -0.003 | -0.011 | 0.001 | 0.001 | 0.002 | 6.49E-10 | 1.27E-04 | 3.22E-08 |
| rs2777799 | A | G | ABCA1 | 5 | -0.003 | -0.003 | -0.009 | 0.001 | 0.001 | 0.003 | 1.20E-04 | 5.51E-03 | 7.74E-04 |
| rs2823146 | G | A | NRIP1_USP25 | 5 | 0.010 | 0.009 | 0.032 | 0.002 | 0.002 | 0.006 | 1.84E-09 | 4.32E-05 | 1.69E-07 |
| rs28480204 | T | G | TOMM40 | 5 | -0.051 | -0.039 | -0.119 | 0.008 | 0.011 | 0.029 | 2.71E-10 | 2.55E-04 | 5.53E-05 |
| rs28638160 | A | C | TCEA2 | 5 | -0.004 | -0.002 | -0.006 | 0.001 | 0.001 | 0.002 | 1.67E-16 | 4.54E-03 | 5.60E-03 |
| rs2865162 | G | A | MAFB_LOC339568 | 5 | 0.006 | 0.005 | 0.016 | 0.001 | 0.001 | 0.003 | 5.04E-12 | 3.12E-05 | 4.52E-07 |
| rs2937749 | G | T | SV2C | 5 | -0.005 | -0.004 | -0.012 | 0.001 | 0.001 | 0.003 | 1.08E-07 | 2.32E-03 | 2.63E-04 |
| rs2940184 | C | T | NF1 | 5 | -0.004 | -0.004 | -0.014 | 0.000 | 0.001 | 0.002 | 2.68E-19 | 2.94E-09 | 1.58E-14 |
| rs3130062 | T | C | NFKBIL1 | 5 | -0.006 | -0.004 | -0.014 | 0.001 | 0.001 | 0.003 | 3.34E-12 | 2.55E-04 | 9.04E-06 |
| rs3170660 | C | T | GPN2 | 5 | -0.004 | -0.002 | -0.006 | 0.000 | 0.001 | 0.002 | 1.71E-14 | 2.77E-03 | 2.08E-03 |
| rs35624358 | A | G | MAFB_LOC339568 | 5 | 0.003 | 0.003 | 0.010 | 0.001 | 0.001 | 0.002 | 5.45E-12 | 3.76E-05 | 4.89E-07 |
| rs35645619 | A | G | PLCG1 | 5 | -0.010 | -0.006 | -0.024 | 0.001 | 0.002 | 0.005 | 5.22E-15 | 7.62E-04 | 1.48E-06 |
| rs35959395 | C | G | PVR | 5 | -0.035 | -0.027 | -0.083 | 0.005 | 0.007 | 0.019 | 5.21E-11 | 8.65E-05 | 1.35E-05 |
| rs3741782 | G | A | CORO1C | 5 | -0.004 | -0.002 | -0.007 | 0.000 | 0.001 | 0.002 | 1.21E-13 | 9.88E-04 | 1.37E-04 |
| rs3820071 | A | G | CELA2B | 5 | -0.004 | -0.003 | -0.006 | 0.001 | 0.001 | 0.002 | 7.28E-12 | 2.94E-04 | 1.76E-03 |
| rs3826408 | T | C | DLG4 | 5 | 0.005 | 0.004 | 0.012 | 0.000 | 0.001 | 0.002 | 1.07E-26 | 2.29E-08 | 5.64E-13 |
| rs41434449 | T | A | PPIB_SNX22 | 5 | -0.004 | -0.004 | -0.013 | 0.001 | 0.001 | 0.003 | 1.39E-10 | 3.08E-05 | 1.06E-06 |
| rs4148008 | G | C | ABCA8 | 5 | -0.004 | -0.003 | -0.010 | 0.000 | 0.001 | 0.002 | 1.38E-14 | 4.23E-06 | 6.02E-08 |
| rs4346380 | A | G | SEPT2 | 5 | 0.004 | 0.003 | 0.009 | 0.001 | 0.001 | 0.002 | 7.05E-10 | 6.56E-04 | 5.80E-05 |
| rs4374942 | C | T | HTR5A_INSIG1 | 5 | -0.004 | -0.004 | -0.015 | 0.001 | 0.001 | 0.003 | 1.47E-07 | 3.21E-04 | 3.24E-06 |
| rs454182 | C | G | GPX5_SCAND3 | 5 | -0.003 | -0.003 | -0.010 | 0.001 | 0.001 | 0.002 | 3.31E-08 | 5.83E-05 | 3.35E-07 |
| rs45575338 | G | A | FAM208B | 5 | 0.003 | 0.002 | 0.005 | 0.001 | 0.001 | 0.002 | 1.58E-08 | 2.48E-03 | 2.46E-02 |
| rs4648892 | T | C | TCEA3 | 5 | -0.003 | -0.003 | -0.010 | 0.001 | 0.001 | 0.002 | 2.21E-09 | 4.99E-06 | 5.31E-08 |
| rs4801172 | G | A | ZFP28 | 5 | 0.003 | 0.003 | 0.009 | 0.001 | 0.001 | 0.002 | 1.04E-09 | 5.12E-04 | 2.36E-06 |
| rs5404 | T | C | SLC2A2 | 5 | -0.005 | -0.005 | -0.014 | 0.001 | 0.001 | 0.003 | 6.86E-12 | 2.02E-05 | 5.93E-07 |
| rs55669835 | G | A | FAM117B | 5 | -0.005 | -0.004 | -0.015 | 0.001 | 0.001 | 0.002 | 4.06E-23 | 2.03E-08 | 2.04E-13 |
| rs55676788 | C | T | CHST4_ZNF19 | 5 | -0.005 | -0.005 | -0.016 | 0.001 | 0.001 | 0.003 | 8.82E-14 | 2.20E-07 | 1.06E-09 |
| rs55824586 | A | G | DPP10_DDX18 | 5 | 0.009 | 0.008 | 0.026 | 0.001 | 0.001 | 0.003 | 3.47E-27 | 2.25E-12 | 6.47E-18 |
| rs55995239 | T | C | C6orf58_SOGA3 | 5 | -0.005 | -0.006 | -0.016 | 0.001 | 0.001 | 0.004 | 1.28E-05 | 1.57E-04 | 3.40E-05 |
| rs56237434 | T | C | MYBPHL | 5 | -0.009 | -0.003 | -0.011 | 0.001 | 0.002 | 0.005 | 4.63E-11 | 7.70E-02 | 2.40E-02 |
| rs59895800 | G | C | BEST4 | 5 | -0.004 | -0.003 | -0.011 | 0.001 | 0.001 | 0.002 | 2.13E-10 | 5.66E-04 | 5.08E-07 |
| rs6022849 | C | G | BCAS1_SUMO1P1 | 5 | -0.003 | -0.002 | -0.009 | 0.000 | 0.001 | 0.002 | 1.64E-08 | 2.37E-04 | 7.31E-07 |
| rs60239918 | T | C | GEMIN7_ZNF296 | 5 | -0.009 | -0.007 | -0.023 | 0.001 | 0.002 | 0.004 | 1.89E-13 | 2.91E-05 | 1.24E-07 |
| rs60701149 | A | G | PMFBP1_ZFHX3 | 5 | 0.006 | 0.007 | 0.025 | 0.001 | 0.001 | 0.003 | 3.48E-13 | 1.44E-08 | 5.14E-14 |
| rs6080762 | C | T | RRBP1 | 5 | 0.004 | 0.002 | 0.010 | 0.001 | 0.001 | 0.002 | 4.07E-10 | 1.13E-02 | 2.64E-05 |
| rs61097887 | C | T | ABCG8 | 5 | 0.003 | 0.002 | 0.007 | 0.000 | 0.001 | 0.002 | 4.21E-11 | 2.50E-03 | 3.51E-05 |
| rs61636751 | A | G | APOB_C2orf43 | 5 | -0.010 | -0.004 | -0.016 | 0.001 | 0.002 | 0.004 | 1.58E-15 | 8.41E-03 | 4.07E-04 |
| rs61642202 | C | A | PVRL2_BCAM | 5 | -0.055 | -0.040 | -0.133 | 0.002 | 0.003 | 0.009 | 3.05E-108 | 3.13E-34 | 1.86E-50 |
| rs61755050 | C | T | NR1H4 | 5 | 0.015 | 0.018 | 0.045 | 0.003 | 0.004 | 0.011 | 5.72E-07 | 2.33E-05 | 5.25E-05 |
| rs61871243 | A | G | NAP1L4 | 5 | -0.004 | -0.004 | -0.011 | 0.001 | 0.001 | 0.003 | 5.75E-09 | 3.87E-05 | 3.59E-05 |
| rs62070652 | T | C | ATAD5 | 5 | -0.004 | -0.003 | -0.010 | 0.001 | 0.001 | 0.002 | 7.69E-12 | 1.99E-04 | 4.43E-07 |
| rs62123888 | C | T | C2orf43_APOB | 5 | -0.006 | -0.005 | -0.021 | 0.001 | 0.001 | 0.004 | 2.19E-11 | 1.54E-04 | 2.34E-09 |
| rs62399429 | A | G | MUC22 | 5 | -0.006 | -0.006 | -0.018 | 0.001 | 0.001 | 0.002 | 4.45E-19 | 2.42E-12 | 2.31E-13 |
| rs62639219 | C | T | IST1 | 5 | -0.012 | -0.010 | -0.030 | 0.002 | 0.002 | 0.006 | 4.00E-14 | 1.07E-05 | 4.13E-07 |
| rs6547409 | T | C | APOB_C2orf43 | 5 | 0.044 | 0.028 | 0.088 | 0.001 | 0.002 | 0.004 | 0.00E+00 | 2.41E-73 | 1.11E-102 |
| rs67143157 | G | A | ASGR1_ASGR2 | 5 | 0.008 | 0.005 | 0.018 | 0.001 | 0.001 | 0.002 | 4.04E-47 | 5.69E-11 | 1.25E-19 |
| rs6733550 | G | T | TMEM150A | 5 | 0.002 | 0.002 | 0.008 | 0.000 | 0.001 | 0.002 | 1.37E-06 | 1.80E-03 | 2.29E-05 |
| rs6750836 | A | C | CCDC93 | 5 | 0.003 | 0.003 | 0.010 | 0.000 | 0.001 | 0.002 | 1.73E-13 | 2.90E-06 | 1.06E-08 |
| rs6761276 | C | T | IL1F10 | 5 | 0.003 | 0.002 | 0.007 | 0.000 | 0.001 | 0.002 | 6.34E-11 | 6.46E-04 | 6.20E-05 |
| rs679224 | G | A | KIAA1324 | 5 | -0.007 | -0.004 | -0.016 | 0.001 | 0.001 | 0.002 | 5.29E-29 | 7.00E-07 | 5.41E-11 |
| rs6908739 | C | T | REPS1 | 5 | -0.004 | -0.003 | -0.010 | 0.001 | 0.001 | 0.002 | 3.72E-12 | 1.34E-04 | 2.48E-05 |
| rs6912701 | T | C | HLA-DRA_BTNL2 | 5 | -0.012 | -0.010 | -0.033 | 0.002 | 0.002 | 0.006 | 4.48E-12 | 1.67E-05 | 1.48E-07 |
| rs7240405 | G | A | LIPG_ACAA2 | 5 | 0.001 | -0.004 | -0.013 | 0.001 | 0.001 | 0.002 | 9.50E-02 | 7.97E-06 | 1.17E-08 |
| rs73011369 | A | G | SMARCA4 | 5 | 0.005 | 0.003 | 0.015 | 0.001 | 0.001 | 0.003 | 9.66E-10 | 5.74E-03 | 2.11E-06 |
| rs73034885 | T | C | EXOC3L2_MARK4 | 5 | -0.008 | -0.004 | -0.016 | 0.001 | 0.002 | 0.005 | 4.77E-10 | 1.02E-02 | 5.05E-04 |
| rs73116882 | T | C | SPINT4_WFDC3 | 5 | 0.005 | 0.004 | 0.013 | 0.001 | 0.001 | 0.003 | 1.44E-08 | 1.92E-03 | 1.83E-04 |
| rs73215978 | C | T | ACPP_DNAJC13 | 5 | 0.006 | 0.006 | 0.023 | 0.001 | 0.001 | 0.003 | 6.09E-12 | 6.72E-06 | 7.43E-11 |
| rs73959582 | C | T | LIPG_ACAA2 | 5 | 0.000 | -0.004 | -0.014 | 0.001 | 0.001 | 0.003 | 7.06E-01 | 9.60E-05 | 4.72E-06 |
| rs7483 | T | C | GSTM3 | 5 | 0.004 | 0.002 | 0.008 | 0.001 | 0.001 | 0.002 | 3.41E-13 | 3.45E-03 | 5.28E-05 |
| rs751102 | T | C | BANF2_SNX5 | 5 | -0.004 | -0.003 | -0.011 | 0.001 | 0.001 | 0.002 | 2.00E-12 | 1.50E-03 | 4.07E-06 |
| rs75211012 | A | C | LOC100507562_APOB | 5 | -0.011 | -0.006 | -0.018 | 0.001 | 0.001 | 0.003 | 4.59E-31 | 5.62E-06 | 1.36E-07 |
| rs76222503 | A | C | LOC157273_TNKS | 5 | -0.005 | -0.006 | -0.022 | 0.001 | 0.002 | 0.004 | 2.49E-06 | 4.09E-04 | 3.01E-07 |
| rs76856627 | G | A | CLASRP | 5 | -0.019 | -0.013 | -0.045 | 0.001 | 0.002 | 0.005 | 5.13E-44 | 4.05E-13 | 2.66E-19 |
| rs769452 | C | T | APOE | 5 | -0.082 | -0.064 | -0.213 | 0.006 | 0.008 | 0.022 | 1.52E-40 | 5.96E-15 | 1.62E-21 |
| rs77319662 | C | T | LOC100652824 | 5 | -0.006 | -0.005 | -0.013 | 0.001 | 0.001 | 0.003 | 1.24E-16 | 5.90E-07 | 9.92E-07 |
| rs7752556 | A | G | KLHL31_GCLC | 5 | -0.004 | -0.003 | -0.009 | 0.001 | 0.001 | 0.002 | 3.96E-15 | 1.50E-05 | 1.14E-05 |
| rs77556405 | A | G | SKAP1 | 5 | -0.005 | -0.003 | -0.011 | 0.001 | 0.001 | 0.002 | 6.97E-14 | 1.97E-03 | 1.83E-06 |
| rs77623030 | T | C | BCAT2_FGF21 | 5 | 0.005 | 0.004 | 0.014 | 0.001 | 0.001 | 0.003 | 2.98E-09 | 3.33E-04 | 2.07E-05 |
| rs78173576 | G | T | GPS2 | 5 | 0.008 | 0.006 | 0.019 | 0.001 | 0.002 | 0.005 | 6.88E-12 | 9.62E-04 | 4.31E-05 |
| rs78175438 | C | T | BRWD1 | 5 | 0.006 | 0.005 | 0.013 | 0.001 | 0.001 | 0.003 | 9.04E-19 | 1.35E-06 | 3.21E-07 |
| rs78376718 | C | T | ANKRD31_HMGCR | 5 | -0.005 | -0.005 | -0.019 | 0.001 | 0.001 | 0.004 | 2.74E-08 | 8.66E-05 | 5.37E-07 |
| rs78432537 | T | C | WWP2 | 5 | -0.007 | -0.005 | -0.017 | 0.001 | 0.001 | 0.004 | 2.75E-14 | 1.42E-04 | 2.06E-06 |
| rs79783247 | G | A | FAM169A_GCNT4 | 5 | -0.014 | -0.014 | -0.044 | 0.002 | 0.002 | 0.007 | 4.69E-15 | 1.21E-08 | 1.79E-11 |
| rs8069974 | C | G | ZMYND15_TM4SF5 | 5 | -0.003 | -0.002 | -0.008 | 0.000 | 0.001 | 0.002 | 6.43E-10 | 3.42E-04 | 2.59E-05 |
| rs8103121 | G | A | SEC1P | 5 | 0.004 | 0.003 | 0.009 | 0.001 | 0.001 | 0.002 | 2.31E-12 | 1.66E-05 | 2.04E-06 |
| rs9320552 | T | C | FRK | 5 | -0.003 | -0.003 | -0.013 | 0.001 | 0.001 | 0.002 | 4.72E-11 | 3.84E-06 | 7.79E-12 |
| rs941796 | A | G | CHD6_PTPRT | 5 | -0.003 | -0.002 | -0.008 | 0.000 | 0.001 | 0.002 | 1.52E-12 | 9.03E-04 | 1.60E-06 |
| rs9438866 | C | A | SYF2_RUNX3 | 5 | 0.006 | 0.004 | 0.014 | 0.001 | 0.001 | 0.002 | 1.76E-20 | 1.31E-05 | 6.64E-09 |
| rs10087499 | T | G | CYP7A1_UBXN2B | 6 | 0.005 | 0.006 | 0.016 | 0.001 | 0.001 | 0.002 | 7.45E-25 | 5.43E-15 | 1.05E-16 |
| rs1042311 | T | C | PPARA | 6 | -0.026 | -0.016 | -0.051 | 0.003 | 0.004 | 0.012 | 1.59E-17 | 2.38E-04 | 1.40E-05 |
| rs10495703 | T | C | SDC1_LAPTM4A | 6 | 0.003 | 0.004 | 0.012 | 0.000 | 0.001 | 0.002 | 1.55E-09 | 8.32E-11 | 1.77E-11 |
| rs10495713 | G | A | APOB_C2orf43 | 6 | 0.006 | 0.005 | 0.012 | 0.000 | 0.001 | 0.002 | 5.36E-43 | 5.19E-16 | 1.18E-11 |
| rs1057868 | T | C | POR | 6 | -0.003 | -0.004 | -0.010 | 0.000 | 0.001 | 0.002 | 2.63E-09 | 3.59E-10 | 1.01E-07 |
| rs10957055 | T | C | CYP7A1_UBXN2B | 6 | -0.006 | -0.007 | -0.020 | 0.001 | 0.001 | 0.003 | 2.50E-11 | 5.65E-09 | 4.58E-10 |
| rs11057602 | C | T | NCOR2 | 6 | 0.003 | 0.003 | 0.008 | 0.000 | 0.001 | 0.002 | 5.51E-13 | 2.77E-07 | 1.87E-06 |
| rs11057830 | A | G | SCARB1 | 6 | -0.008 | -0.006 | -0.013 | 0.001 | 0.001 | 0.003 | 1.69E-29 | 3.76E-10 | 4.75E-07 |
| rs11128152 | T | C | MITF | 6 | 0.003 | 0.003 | 0.009 | 0.001 | 0.001 | 0.002 | 1.54E-07 | 3.15E-05 | 3.64E-05 |
| rs1123021 | G | A | KDM1A | 6 | -0.004 | -0.004 | -0.010 | 0.001 | 0.001 | 0.002 | 9.51E-11 | 4.68E-06 | 1.18E-05 |
| rs113172657 | ACTG | A | DNAH10 | 6 | 0.004 | 0.003 | 0.008 | 0.001 | 0.001 | 0.002 | 3.54E-12 | 7.60E-06 | 9.60E-05 |
| rs114488427 | T | C | VPS4A_COG8 | 6 | -0.004 | -0.003 | -0.007 | 0.001 | 0.001 | 0.003 | 7.02E-10 | 5.62E-04 | 8.53E-03 |
| rs114878174 | G | A | HCP5_MICA | 6 | -0.010 | -0.013 | -0.036 | 0.002 | 0.002 | 0.007 | 8.63E-09 | 3.39E-08 | 4.17E-08 |
| rs114969413 | A | G | NELFE | 6 | -0.008 | -0.008 | -0.025 | 0.001 | 0.002 | 0.005 | 1.66E-10 | 5.38E-06 | 6.23E-08 |
| rs11610045 | A | G | FBRSL1_MUC8 | 6 | -0.003 | -0.003 | -0.008 | 0.000 | 0.001 | 0.002 | 9.66E-13 | 1.24E-06 | 5.97E-06 |
| rs117198034 | T | C | CLPTM1 | 6 | 0.013 | 0.015 | 0.035 | 0.002 | 0.002 | 0.006 | 2.43E-14 | 2.50E-11 | 1.15E-08 |
| rs117414940 | C | A | ZNF14_LINC00663 | 6 | 0.017 | 0.021 | 0.051 | 0.002 | 0.002 | 0.006 | 2.65E-24 | 1.14E-20 | 7.69E-17 |
| rs117558284 | C | T | ZFHX3 | 6 | -0.012 | -0.010 | -0.032 | 0.001 | 0.002 | 0.005 | 6.27E-18 | 2.87E-07 | 1.84E-09 |
| rs117789739 | T | C | PPP1R37_MARK4 | 6 | 0.008 | 0.007 | 0.017 | 0.001 | 0.002 | 0.005 | 1.43E-07 | 3.10E-04 | 1.73E-03 |
| rs117874826 | C | A | PLCB3 | 6 | -0.013 | -0.013 | -0.024 | 0.002 | 0.003 | 0.007 | 2.98E-11 | 5.51E-06 | 1.20E-03 |
| rs11803457 | C | T | RPAP2_GFI1 | 6 | -0.003 | -0.004 | -0.012 | 0.000 | 0.001 | 0.002 | 1.78E-11 | 4.60E-10 | 1.59E-12 |
| rs118092024 | C | A | NLRC5 | 6 | 0.008 | 0.007 | 0.021 | 0.001 | 0.002 | 0.005 | 1.05E-10 | 5.98E-05 | 1.56E-05 |
| rs11872886 | T | C | LIPG_ACAA2 | 6 | 0.000 | -0.003 | -0.006 | 0.000 | 0.001 | 0.002 | 3.96E-01 | 6.38E-05 | 8.71E-04 |
| rs12187482 | G | A | TIMD4_PPP1R2P3 | 6 | -0.003 | -0.003 | -0.009 | 0.001 | 0.001 | 0.002 | 4.22E-08 | 3.71E-04 | 1.37E-05 |
| rs12410656 | T | C | FAM46B_SLC9A1 | 6 | -0.010 | -0.007 | -0.015 | 0.001 | 0.001 | 0.003 | 5.50E-26 | 3.27E-07 | 1.67E-05 |
| rs12444980 | G | A | ZFPM1 | 6 | -0.005 | -0.006 | -0.019 | 0.001 | 0.002 | 0.004 | 5.17E-06 | 3.02E-05 | 2.18E-06 |
| rs12450700 | G | T | PSMD12_PITPNC1 | 6 | 0.004 | 0.003 | 0.006 | 0.000 | 0.001 | 0.002 | 7.32E-14 | 1.51E-05 | 3.16E-04 |
| rs12460388 | T | C | BCL3_CEACAM16 | 6 | 0.006 | 0.006 | 0.019 | 0.001 | 0.001 | 0.004 | 1.36E-09 | 1.85E-05 | 5.42E-07 |
| rs12869880 | A | G | GAS6_GAS6-AS1 | 6 | -0.006 | -0.008 | -0.022 | 0.001 | 0.001 | 0.003 | 1.11E-13 | 4.91E-11 | 2.07E-11 |
| rs12927044 | A | C | IST1 | 6 | 0.006 | 0.006 | 0.021 | 0.001 | 0.001 | 0.003 | 3.26E-14 | 4.78E-09 | 2.77E-13 |
| rs12946388 | C | T | MAP2K6 | 6 | 0.003 | 0.002 | 0.006 | 0.000 | 0.001 | 0.002 | 1.25E-10 | 3.62E-03 | 7.81E-04 |
| rs13203469 | A | C | L3MBTL3_TMEM244 | 6 | -0.003 | -0.003 | -0.008 | 0.000 | 0.001 | 0.002 | 6.57E-10 | 7.74E-06 | 8.91E-06 |
| rs133084 | C | T | MCHR1_SLC25A17 | 6 | -0.003 | -0.002 | -0.008 | 0.000 | 0.001 | 0.002 | 2.74E-10 | 2.11E-04 | 1.77E-06 |
| rs138607350 | G | T | PVRL2 | 6 | -0.044 | -0.037 | -0.098 | 0.003 | 0.004 | 0.010 | 8.35E-58 | 7.92E-25 | 2.45E-23 |
| rs139267469 | A | C | PVR | 6 | -0.036 | -0.029 | -0.084 | 0.005 | 0.007 | 0.019 | 3.30E-12 | 1.54E-05 | 5.88E-06 |
| rs146125856 | C | T | USP8 | 6 | -0.002 | -0.002 | -0.004 | 0.001 | 0.001 | 0.003 | 8.44E-03 | 8.27E-02 | 1.83E-01 |
| rs154977 | G | C | HLA-DMB_LOC100294145 | 6 | -0.003 | -0.004 | -0.007 | 0.000 | 0.001 | 0.002 | 9.62E-09 | 3.65E-08 | 8.98E-05 |
| rs163428 | T | C | ITK | 6 | 0.003 | 0.003 | 0.009 | 0.000 | 0.001 | 0.002 | 2.45E-14 | 3.15E-06 | 4.60E-08 |
| rs17208000 | T | G | FKBPL_PRRT1 | 6 | -0.005 | -0.006 | -0.014 | 0.001 | 0.001 | 0.003 | 1.38E-12 | 6.23E-10 | 4.81E-07 |
| rs17392679 | T | C | SECISBP2L | 6 | 0.005 | 0.004 | 0.013 | 0.001 | 0.001 | 0.003 | 7.32E-10 | 3.62E-05 | 1.51E-05 |
| rs17699030 | G | A | DOCK6 | 6 | 0.006 | 0.014 | 0.036 | 0.001 | 0.002 | 0.005 | 2.15E-06 | 9.00E-15 | 5.76E-14 |
| rs17712208 | A | T | PROX1_LINC00538 | 6 | -0.007 | -0.007 | -0.011 | 0.001 | 0.002 | 0.005 | 7.26E-08 | 2.02E-04 | 1.66E-02 |
| rs17789218 | C | T | SIM1_LOC728012 | 6 | 0.006 | 0.006 | 0.016 | 0.001 | 0.001 | 0.002 | 5.70E-26 | 8.28E-16 | 5.68E-15 |
| rs1800027 | T | C | FUT2 | 6 | 0.009 | 0.008 | 0.022 | 0.001 | 0.001 | 0.004 | 6.26E-19 | 5.04E-09 | 1.04E-09 |
| rs1809165 | C | T | TRIB1_LINC00861 | 6 | -0.010 | -0.011 | -0.027 | 0.001 | 0.002 | 0.004 | 4.76E-18 | 2.77E-11 | 5.36E-10 |
| rs203273 | T | C | RBM47_CHRNA9 | 6 | -0.004 | -0.002 | -0.008 | 0.000 | 0.001 | 0.002 | 2.92E-14 | 2.10E-03 | 4.77E-06 |
| rs2047774 | A | G | ASGR2_ASGR1 | 6 | -0.003 | -0.002 | -0.006 | 0.000 | 0.001 | 0.002 | 2.30E-09 | 3.31E-03 | 7.79E-04 |
| rs2073717 | C | G | CCHCR1 | 6 | -0.003 | -0.003 | -0.006 | 0.000 | 0.001 | 0.002 | 3.21E-09 | 1.03E-06 | 1.95E-04 |
| rs2153271 | T | C | BNC2 | 6 | 0.004 | 0.004 | 0.009 | 0.000 | 0.001 | 0.002 | 3.46E-20 | 1.72E-11 | 9.39E-08 |
| rs2228603 | T | C | NCAN | 6 | 0.024 | 0.030 | 0.078 | 0.001 | 0.001 | 0.003 | 1.55E-152 | 2.93E-129 | 5.01E-117 |
| rs2240068 | T | G | TRIM31 | 6 | -0.003 | -0.003 | -0.009 | 0.000 | 0.001 | 0.002 | 7.10E-11 | 2.83E-06 | 2.37E-07 |
| rs228781 | C | T | TMEM101_LSM12 | 6 | 0.006 | 0.005 | 0.013 | 0.001 | 0.001 | 0.003 | 1.29E-11 | 9.70E-05 | 1.28E-04 |
| rs2498323 | A | G | HGFAC | 6 | -0.007 | -0.007 | -0.017 | 0.001 | 0.001 | 0.003 | 9.70E-21 | 7.08E-10 | 1.50E-09 |
| rs2596501 | T | C | HLA-B_HLA-C | 6 | -0.003 | -0.004 | -0.009 | 0.000 | 0.001 | 0.002 | 2.10E-13 | 1.19E-09 | 7.05E-07 |
| rs278981 | C | T | RBM47 | 6 | -0.004 | -0.004 | -0.012 | 0.001 | 0.001 | 0.002 | 1.42E-14 | 2.48E-07 | 3.07E-09 |
| rs28378222 | A | G | MITF | 6 | -0.003 | -0.004 | -0.009 | 0.000 | 0.001 | 0.002 | 4.67E-11 | 4.21E-08 | 1.04E-07 |
| rs2844477 | C | T | NCR3_AIF1 | 6 | -0.003 | -0.003 | -0.007 | 0.000 | 0.001 | 0.002 | 4.05E-12 | 3.22E-05 | 5.13E-05 |
| rs2875973 | T | C | RAB2A | 6 | -0.002 | -0.003 | -0.009 | 0.000 | 0.001 | 0.002 | 7.48E-06 | 3.51E-05 | 1.71E-06 |
| rs289715 | T | A | CETP | 6 | -0.008 | -0.006 | -0.014 | 0.001 | 0.001 | 0.003 | 1.57E-28 | 4.98E-09 | 5.02E-08 |
| rs291044 | A | G | CETP | 6 | -0.006 | -0.004 | -0.011 | 0.000 | 0.001 | 0.002 | 2.05E-33 | 3.83E-08 | 8.44E-09 |
| rs3117286 | C | T | MOG | 6 | -0.007 | -0.008 | -0.022 | 0.001 | 0.002 | 0.005 | 5.34E-07 | 2.41E-05 | 1.33E-05 |
| rs312966 | G | A | APOB_LOC645949 | 6 | 0.005 | 0.005 | 0.014 | 0.000 | 0.001 | 0.002 | 3.18E-26 | 1.43E-11 | 8.26E-15 |
| rs332923 | G | T | DPP10_DDX18 | 6 | 0.004 | 0.004 | 0.009 | 0.001 | 0.001 | 0.002 | 2.91E-08 | 4.96E-05 | 9.59E-05 |
| rs34030799 | A | G | AMPD2 | 6 | -0.009 | -0.005 | -0.017 | 0.001 | 0.002 | 0.006 | 2.78E-09 | 1.48E-02 | 2.73E-03 |
| rs34041362 | G | T | TMPRSS11E_YTHDC1 | 6 | -0.004 | -0.004 | -0.010 | 0.001 | 0.001 | 0.002 | 1.75E-13 | 1.04E-08 | 8.80E-08 |
| rs34208166 | G | A | H3F3A_GALK1 | 6 | 0.005 | 0.004 | 0.010 | 0.001 | 0.001 | 0.003 | 2.00E-11 | 5.09E-05 | 1.52E-04 |
| rs35337492 | A | G | LIPG_ACAA2 | 6 | 0.000 | -0.003 | -0.007 | 0.000 | 0.001 | 0.002 | 7.57E-01 | 1.08E-04 | 8.94E-05 |
| rs35641646 | C | T | LIPG | 6 | -0.002 | -0.005 | -0.014 | 0.001 | 0.001 | 0.003 | 5.93E-02 | 4.42E-05 | 4.17E-05 |
| rs362285 | A | G | MSANTD1 | 6 | -0.012 | -0.009 | -0.021 | 0.002 | 0.002 | 0.007 | 1.86E-12 | 1.75E-04 | 1.74E-03 |
| rs3744010 | A | G | UNC13D | 6 | 0.004 | 0.004 | 0.007 | 0.001 | 0.001 | 0.002 | 1.49E-14 | 3.27E-07 | 6.31E-04 |
| rs3744262 | A | G | EFNB3 | 6 | 0.004 | 0.004 | 0.011 | 0.000 | 0.001 | 0.002 | 1.86E-14 | 1.07E-08 | 1.23E-08 |
| rs3761081 | A | G | MEF2BNB-MEF2B_MEF2BNB | 6 | -0.005 | -0.004 | -0.013 | 0.001 | 0.001 | 0.002 | 1.20E-14 | 9.21E-08 | 4.34E-09 |
| rs3764567 | T | C | MAU2 | 6 | -0.007 | -0.008 | -0.020 | 0.001 | 0.001 | 0.002 | 2.31E-40 | 5.89E-35 | 2.58E-27 |
| rs3771241 | A | G | SDC1 | 6 | 0.003 | 0.004 | 0.009 | 0.000 | 0.001 | 0.002 | 7.90E-11 | 1.28E-07 | 1.71E-06 |
| rs3815692 | A | C | SULT2B1 | 6 | 0.007 | 0.006 | 0.018 | 0.001 | 0.001 | 0.004 | 1.47E-12 | 1.10E-05 | 6.64E-07 |
| rs3828191 | T | C | GRB14 | 6 | -0.003 | -0.004 | -0.008 | 0.000 | 0.001 | 0.002 | 1.87E-09 | 4.38E-08 | 9.23E-06 |
| rs409558 | C | T | MSH5_SAPCD1 | 6 | -0.006 | -0.007 | -0.017 | 0.001 | 0.001 | 0.002 | 1.21E-18 | 2.46E-14 | 1.85E-12 |
| rs41302776 | A | G | PIK3R3 | 6 | 0.007 | 0.008 | 0.021 | 0.001 | 0.002 | 0.004 | 2.34E-10 | 2.73E-06 | 1.69E-06 |
| rs463728 | A | G | SGCD | 6 | -0.005 | -0.004 | -0.011 | 0.001 | 0.001 | 0.002 | 1.03E-12 | 8.21E-07 | 5.28E-06 |
| rs4683702 | G | T | PCOLCE2_LOC100507389 | 6 | 0.003 | 0.003 | 0.010 | 0.000 | 0.001 | 0.002 | 1.67E-11 | 3.77E-07 | 1.35E-07 |
| rs4690096 | T | G | RGS12 | 6 | 0.003 | 0.003 | 0.007 | 0.000 | 0.001 | 0.002 | 6.50E-12 | 1.77E-07 | 4.92E-05 |
| rs471364 | T | C | TTC39B | 6 | -0.005 | -0.003 | -0.009 | 0.001 | 0.001 | 0.003 | 1.71E-10 | 2.40E-03 | 5.37E-04 |
| rs4847327 | G | A | EVI5 | 6 | -0.005 | -0.005 | -0.017 | 0.001 | 0.001 | 0.003 | 2.00E-11 | 1.39E-06 | 1.87E-09 |
| rs484959 | C | T | EPS8L3_CSF1 | 6 | -0.003 | -0.002 | -0.006 | 0.000 | 0.001 | 0.002 | 1.13E-11 | 2.13E-04 | 4.39E-04 |
| rs4895815 | G | A | CENPW_RSPO3 | 6 | 0.004 | 0.003 | 0.010 | 0.001 | 0.001 | 0.002 | 6.20E-11 | 3.97E-06 | 1.27E-06 |
| rs547976 | A | G | SRSF4 | 6 | 0.003 | 0.003 | 0.006 | 0.001 | 0.001 | 0.002 | 6.57E-10 | 9.68E-06 | 3.64E-03 |
| rs55784804 | T | G | SHBG | 6 | -0.006 | -0.006 | -0.016 | 0.001 | 0.001 | 0.003 | 1.27E-16 | 3.62E-08 | 1.99E-08 |
| rs56007657 | C | T | NCAN_NR2C2AP | 6 | -0.004 | -0.005 | -0.011 | 0.001 | 0.001 | 0.003 | 1.20E-07 | 8.45E-08 | 1.83E-05 |
| rs60049679 | C | G | APOC1P1_APOC1 | 6 | -0.039 | -0.031 | -0.090 | 0.001 | 0.001 | 0.003 | 0.00E+00 | 2.00E-135 | 5.37E-156 |
| rs6060369 | C | T | UQCC | 6 | 0.003 | 0.003 | 0.009 | 0.000 | 0.001 | 0.002 | 1.05E-11 | 5.59E-06 | 1.49E-07 |
| rs6074032 | T | G | CD40_CDH22 | 6 | -0.003 | -0.003 | -0.005 | 0.000 | 0.001 | 0.002 | 1.10E-10 | 5.91E-05 | 2.64E-03 |
| rs6085661 | T | C | BMP2_FERMT1 | 6 | -0.003 | -0.002 | -0.005 | 0.000 | 0.001 | 0.002 | 1.99E-09 | 8.39E-03 | 4.66E-03 |
| rs61361928 | C | T | UGT2B7 | 6 | 0.022 | 0.023 | 0.059 | 0.003 | 0.004 | 0.012 | 1.40E-12 | 1.23E-07 | 5.88E-07 |
| rs62111293 | C | T | NOVA2_NANOS2 | 6 | -0.005 | -0.004 | -0.013 | 0.001 | 0.001 | 0.002 | 1.02E-15 | 7.69E-06 | 1.59E-07 |
| rs62210638 | G | C | MAFB_LOC339568 | 6 | 0.006 | 0.005 | 0.016 | 0.001 | 0.001 | 0.003 | 1.64E-19 | 8.04E-07 | 8.19E-09 |
| rs62382402 | T | C | TIMD4_HAVCR1 | 6 | 0.006 | 0.008 | 0.019 | 0.001 | 0.001 | 0.002 | 9.26E-24 | 1.74E-17 | 4.59E-15 |
| rs6457401 | T | G | HLA-B_MICA | 6 | -0.004 | -0.006 | -0.014 | 0.001 | 0.001 | 0.002 | 6.12E-14 | 2.57E-12 | 7.32E-11 |
| rs6722626 | G | A | LOC645949 | 6 | 0.005 | 0.005 | 0.016 | 0.001 | 0.001 | 0.003 | 2.08E-09 | 2.96E-05 | 2.57E-06 |
| rs6753777 | C | T | C2orf43_GDF7 | 6 | -0.007 | -0.006 | -0.016 | 0.001 | 0.001 | 0.003 | 3.55E-21 | 3.11E-09 | 8.49E-08 |
| rs6756943 | A | G | SERTAD2_LOC400958 | 6 | -0.003 | -0.004 | -0.011 | 0.000 | 0.001 | 0.002 | 4.57E-12 | 9.58E-09 | 3.63E-09 |
| rs6934645 | A | C | HLA-DMB | 6 | -0.009 | -0.012 | -0.029 | 0.002 | 0.002 | 0.007 | 3.06E-07 | 3.29E-07 | 7.84E-06 |
| rs6992093 | A | G | NAT2_NAT1 | 6 | 0.003 | 0.004 | 0.008 | 0.000 | 0.001 | 0.002 | 8.08E-08 | 8.88E-08 | 2.30E-05 |
| rs7157785 | T | G | SGPP1_SYNE2 | 6 | -0.004 | -0.006 | -0.014 | 0.001 | 0.001 | 0.002 | 4.75E-13 | 2.10E-11 | 9.50E-10 |
| rs7166771 | C | T | USP3_CA12 | 6 | 0.003 | 0.003 | 0.008 | 0.000 | 0.001 | 0.002 | 1.27E-08 | 7.30E-06 | 4.60E-06 |
| rs7257072 | T | C | MEF2BNB-MEF2B_MEF2B | 6 | 0.003 | 0.004 | 0.011 | 0.000 | 0.001 | 0.002 | 9.29E-11 | 4.23E-09 | 1.54E-09 |
| rs7259930 | A | G | FGF21_BCAT2 | 6 | 0.004 | 0.003 | 0.010 | 0.001 | 0.001 | 0.002 | 6.47E-13 | 1.92E-04 | 1.06E-05 |
| rs72722651 | G | A | TRIB1_LINC00861 | 6 | -0.014 | -0.013 | -0.039 | 0.001 | 0.002 | 0.006 | 5.51E-21 | 1.66E-10 | 1.71E-12 |
| rs72823015 | T | G | ADRB1_NHLRC2 | 6 | 0.005 | 0.006 | 0.017 | 0.001 | 0.001 | 0.003 | 1.23E-14 | 9.69E-09 | 5.49E-11 |
| rs72851052 | T | C | HLA-DRB5_HLA-DRB6 | 6 | 0.002 | 0.004 | 0.008 | 0.000 | 0.001 | 0.002 | 5.40E-07 | 3.04E-08 | 1.34E-05 |
| rs73004826 | A | G | HOMER3_SUGP2 | 6 | 0.016 | 0.018 | 0.045 | 0.001 | 0.002 | 0.005 | 2.82E-30 | 3.07E-21 | 4.97E-18 |
| rs73153160 | A | G | ZNF512B_SOX18 | 6 | -0.004 | -0.005 | -0.012 | 0.001 | 0.001 | 0.003 | 1.09E-08 | 4.68E-07 | 4.47E-06 |
| rs73239113 | C | T | GDF7_HS1BP3 | 6 | 0.004 | 0.003 | 0.009 | 0.001 | 0.001 | 0.003 | 4.86E-10 | 3.60E-03 | 4.65E-04 |
| rs73302510 | T | C | MAFB_LOC339568 | 6 | 0.005 | 0.005 | 0.014 | 0.001 | 0.001 | 0.003 | 1.86E-11 | 1.09E-06 | 6.13E-07 |
| rs73893999 | G | A | LINC00654_LOC643406 | 6 | -0.011 | -0.009 | -0.023 | 0.001 | 0.002 | 0.006 | 1.28E-14 | 2.36E-05 | 3.44E-05 |
| rs74995823 | G | T | LOC100507562_APOB | 6 | 0.011 | 0.009 | 0.025 | 0.001 | 0.002 | 0.006 | 7.17E-13 | 9.78E-06 | 9.11E-06 |
| rs750516 | C | A | MAF_WWOX | 6 | 0.004 | 0.003 | 0.010 | 0.001 | 0.001 | 0.002 | 1.51E-08 | 2.47E-04 | 3.23E-05 |
| rs7534162 | A | G | EFNA3_EFNA1 | 6 | -0.003 | -0.002 | -0.006 | 0.000 | 0.001 | 0.002 | 5.60E-09 | 2.91E-04 | 4.75E-04 |
| rs75916629 | G | A | ZNF90 | 6 | 0.014 | 0.019 | 0.045 | 0.002 | 0.002 | 0.006 | 2.76E-18 | 1.66E-17 | 4.49E-14 |
| rs76530229 | A | G | ZNF14_ZNF101 | 6 | 0.010 | 0.009 | 0.029 | 0.001 | 0.001 | 0.004 | 1.79E-22 | 4.03E-13 | 4.04E-16 |
| rs76786359 | G | A | SGMS1 | 6 | -0.011 | -0.011 | -0.032 | 0.002 | 0.002 | 0.006 | 1.13E-13 | 8.03E-07 | 3.99E-08 |
| rs76995491 | G | A | LOC100507562_APOB | 6 | -0.010 | -0.006 | -0.020 | 0.001 | 0.001 | 0.003 | 2.20E-39 | 6.54E-09 | 3.15E-13 |
| rs77134701 | C | T | CLCC1_AKNAD1 | 6 | 0.009 | 0.007 | 0.021 | 0.001 | 0.002 | 0.004 | 2.53E-16 | 4.60E-06 | 1.19E-06 |
| rs77480718 | G | A | FER1L4_SPAG4 | 6 | 0.006 | 0.006 | 0.017 | 0.001 | 0.001 | 0.004 | 2.60E-09 | 4.08E-06 | 2.06E-06 |
| rs7748291 | T | C | PIM1_FGD2 | 6 | 0.003 | 0.003 | 0.009 | 0.000 | 0.001 | 0.002 | 1.43E-10 | 5.67E-05 | 6.00E-08 |
| rs77987064 | T | C | FAM110B_UBXN2B | 6 | -0.005 | -0.007 | -0.017 | 0.001 | 0.002 | 0.004 | 2.39E-06 | 3.79E-06 | 5.57E-05 |
| rs7860634 | A | G | LHX3 | 6 | 0.003 | 0.002 | 0.008 | 0.000 | 0.001 | 0.002 | 5.84E-12 | 1.48E-04 | 4.25E-06 |
| rs7920112 | C | T | VIM-AS1_VIM | 6 | -0.002 | -0.004 | -0.009 | 0.000 | 0.001 | 0.002 | 1.83E-07 | 1.91E-10 | 9.71E-08 |
| rs79532776 | G | C | FGR | 6 | -0.006 | -0.005 | -0.013 | 0.001 | 0.001 | 0.004 | 1.09E-11 | 5.74E-05 | 2.16E-04 |
| rs8062292 | C | T | NUP93 | 6 | 0.004 | 0.003 | 0.009 | 0.000 | 0.001 | 0.002 | 3.09E-18 | 4.73E-06 | 6.34E-07 |
| rs8063291 | C | T | SLC12A3 | 6 | -0.003 | -0.004 | -0.008 | 0.001 | 0.001 | 0.002 | 2.04E-08 | 2.94E-05 | 3.04E-04 |
| rs8082188 | T | C | STAT5B | 6 | -0.003 | -0.004 | -0.010 | 0.000 | 0.001 | 0.002 | 3.33E-12 | 5.39E-09 | 2.22E-07 |
| rs873870 | A | G | LPAR2 | 6 | 0.004 | 0.005 | 0.014 | 0.000 | 0.001 | 0.002 | 4.50E-18 | 5.87E-16 | 4.00E-16 |
| rs903453 | C | T | NPTX2_BAIAP2L1 | 6 | -0.006 | -0.004 | -0.010 | 0.001 | 0.001 | 0.003 | 3.23E-11 | 1.65E-03 | 3.16E-03 |
| rs926664 | G | A | MAFB_TOP1 | 6 | -0.011 | -0.009 | -0.028 | 0.001 | 0.002 | 0.006 | 1.03E-13 | 5.20E-06 | 4.58E-07 |
| rs9372114 | C | T | PRDM1_PREP | 6 | -0.003 | -0.003 | -0.006 | 0.000 | 0.001 | 0.002 | 1.00E-11 | 4.67E-06 | 2.38E-04 |
| rs9465733 | C | A | E2F3 | 6 | -0.003 | -0.003 | -0.008 | 0.000 | 0.001 | 0.002 | 7.52E-10 | 1.96E-06 | 2.57E-05 |
| rs1015109 | A | G | TRIB1_LINC00861 | 7 | -0.005 | -0.007 | -0.011 | 0.001 | 0.001 | 0.002 | 3.11E-13 | 9.37E-14 | 3.83E-06 |
| rs10164749 | A | G | PUM2 | 7 | 0.003 | 0.006 | 0.011 | 0.001 | 0.001 | 0.002 | 6.49E-08 | 9.95E-10 | 1.42E-05 |
| rs10171839 | A | G | CXCR1_ARPC2 | 7 | 0.002 | 0.002 | 0.004 | 0.000 | 0.001 | 0.002 | 6.42E-05 | 3.53E-04 | 2.82E-02 |
| rs10498200 | A | G | MIR5702_LOC646736 | 7 | 0.002 | 0.003 | 0.002 | 0.001 | 0.001 | 0.002 | 3.46E-05 | 3.43E-04 | 3.90E-01 |
| rs1063412 | A | G | PIGC_C1orf105 | 7 | 0.002 | 0.003 | 0.003 | 0.000 | 0.001 | 0.002 | 1.36E-07 | 4.54E-05 | 6.44E-02 |
| rs10851475 | G | A | FAM227B_GALK2 | 7 | 0.003 | 0.003 | 0.005 | 0.000 | 0.001 | 0.002 | 1.47E-09 | 2.09E-05 | 3.29E-03 |
| rs11078596 | T | C | MIR22HG | 7 | -0.004 | -0.005 | -0.009 | 0.001 | 0.001 | 0.002 | 1.12E-11 | 8.99E-10 | 2.38E-05 |
| rs11134551 | G | A | HAVCR2_HAVCR1 | 7 | -0.004 | -0.006 | -0.012 | 0.001 | 0.001 | 0.002 | 1.19E-13 | 4.41E-11 | 8.00E-08 |
| rs111508381 | A | C | HS1BP3 | 7 | 0.009 | 0.008 | 0.017 | 0.001 | 0.002 | 0.005 | 1.42E-10 | 4.79E-05 | 1.46E-03 |
| rs111620326 | A | G | MEF2BNB-MEF2B_TMEM161A | 7 | 0.005 | 0.007 | 0.018 | 0.001 | 0.001 | 0.004 | 4.42E-07 | 2.60E-07 | 2.56E-07 |
| rs11187157 | C | T | EXOC6_HHEX | 7 | 0.002 | 0.003 | 0.004 | 0.000 | 0.001 | 0.002 | 6.64E-08 | 1.40E-05 | 1.27E-02 |
| rs11187225 | G | C | EXOC6 | 7 | 0.005 | 0.005 | 0.012 | 0.001 | 0.001 | 0.003 | 8.38E-13 | 4.51E-07 | 1.49E-05 |
| rs11187279 | T | C | CYP26A1_MYOF | 7 | -0.003 | -0.003 | -0.008 | 0.000 | 0.001 | 0.002 | 1.26E-09 | 3.76E-07 | 1.86E-05 |
| rs112639655 | A | G | R3HDM2_INHBC | 7 | 0.007 | 0.008 | 0.012 | 0.001 | 0.001 | 0.003 | 3.97E-17 | 6.97E-12 | 1.04E-04 |
| rs115482652 | G | A | VEGFA_LOC100132354 | 7 | -0.011 | -0.012 | -0.022 | 0.002 | 0.003 | 0.007 | 4.57E-10 | 2.61E-06 | 1.13E-03 |
| rs11584073 | G | A | KANK4_USP1 | 7 | -0.004 | -0.006 | -0.011 | 0.001 | 0.001 | 0.004 | 5.60E-05 | 1.34E-06 | 2.52E-03 |
| rs116226697 | C | T | MICB | 7 | -0.009 | -0.011 | -0.023 | 0.002 | 0.002 | 0.006 | 2.30E-09 | 6.51E-07 | 5.32E-05 |
| rs117189332 | T | C | CYP26A1_MYOF | 7 | 0.009 | 0.011 | 0.020 | 0.002 | 0.002 | 0.007 | 6.49E-07 | 6.80E-06 | 2.09E-03 |
| rs117326714 | G | A | CBLC | 7 | 0.008 | 0.012 | 0.027 | 0.002 | 0.002 | 0.006 | 3.08E-07 | 6.60E-09 | 9.01E-07 |
| rs117573122 | C | G | POLR2A | 7 | -0.015 | -0.021 | -0.036 | 0.003 | 0.004 | 0.010 | 2.27E-08 | 3.29E-08 | 4.73E-04 |
| rs117623631 | T | C | LIPG | 7 | 0.005 | -0.035 | -0.070 | 0.005 | 0.007 | 0.018 | 2.54E-01 | 1.08E-07 | 6.13E-05 |
| rs117673551 | A | G | TRIB1_LINC00861 | 7 | 0.016 | 0.020 | 0.030 | 0.001 | 0.002 | 0.005 | 7.14E-36 | 1.98E-27 | 6.65E-10 |
| rs11781960 | C | G | TRIB1_LINC00861 | 7 | 0.011 | 0.016 | 0.026 | 0.001 | 0.002 | 0.006 | 5.92E-13 | 4.13E-14 | 2.51E-06 |
| rs118140434 | G | A | TRIB1_LINC00861 | 7 | -0.011 | -0.013 | -0.021 | 0.002 | 0.003 | 0.007 | 3.88E-09 | 6.36E-07 | 3.00E-03 |
| rs12202204 | G | A | CENPW_RSPO3 | 7 | -0.004 | -0.005 | -0.012 | 0.000 | 0.001 | 0.002 | 2.17E-15 | 8.83E-12 | 2.74E-10 |
| rs12225230 | C | G | SIK3 | 7 | -0.005 | -0.007 | -0.011 | 0.001 | 0.001 | 0.002 | 1.29E-15 | 2.49E-15 | 1.87E-06 |
| rs12406542 | G | A | ATG4C | 7 | -0.003 | -0.007 | -0.011 | 0.001 | 0.001 | 0.004 | 5.35E-03 | 7.10E-06 | 5.39E-03 |
| rs12448528 | G | A | HERPUD1_CETP | 7 | 0.010 | 0.008 | 0.018 | 0.001 | 0.001 | 0.002 | 1.15E-68 | 2.57E-27 | 1.49E-17 |
| rs12514918 | A | G | TIMD4_PPP1R2P3 | 7 | -0.009 | -0.012 | -0.027 | 0.002 | 0.002 | 0.006 | 2.31E-08 | 1.45E-07 | 4.16E-06 |
| rs12721054 | G | A | APOC1 | 7 | 0.031 | 0.047 | 0.109 | 0.005 | 0.007 | 0.020 | 5.09E-09 | 3.29E-11 | 2.86E-08 |
| rs13067911 | C | T | PPARG_TSEN2 | 7 | 0.003 | 0.004 | 0.005 | 0.001 | 0.001 | 0.002 | 2.18E-10 | 4.32E-09 | 4.50E-03 |
| rs13265978 | C | T | TRPS1 | 7 | -0.006 | -0.006 | -0.010 | 0.001 | 0.001 | 0.003 | 1.22E-12 | 8.72E-07 | 1.07E-03 |
| rs13266634 | T | C | SLC30A8 | 7 | 0.003 | 0.003 | 0.004 | 0.000 | 0.001 | 0.002 | 1.20E-07 | 1.45E-05 | 3.56E-02 |
| rs13734 | A | G | RRBP1 | 7 | -0.004 | -0.002 | -0.007 | 0.001 | 0.001 | 0.002 | 2.66E-11 | 9.24E-03 | 1.52E-03 |
| rs139271800 | G | A | PLIN1 | 7 | 0.034 | 0.044 | 0.037 | 0.006 | 0.009 | 0.025 | 9.86E-08 | 2.58E-06 | 1.40E-01 |
| rs141226346 | G | C | ZNF285 | 7 | 0.049 | 0.066 | 0.107 | 0.008 | 0.011 | 0.029 | 3.05E-10 | 4.36E-10 | 2.16E-04 |
| rs145502455 | A | G | PLCB3 | 7 | -0.026 | -0.023 | -0.028 | 0.004 | 0.005 | 0.014 | 2.82E-12 | 6.93E-06 | 4.24E-02 |
| rs149793040 | G | A | ACACB | 7 | 0.041 | 0.057 | 0.104 | 0.005 | 0.007 | 0.020 | 1.63E-15 | 1.77E-14 | 1.91E-07 |
| rs150358728 | T | G | PVRL2 | 7 | 0.014 | 0.014 | 0.030 | 0.002 | 0.003 | 0.008 | 5.47E-10 | 8.03E-07 | 2.20E-04 |
| rs1556124 | A | G | COL4A2 | 7 | -0.003 | -0.004 | -0.009 | 0.001 | 0.001 | 0.002 | 1.76E-09 | 9.61E-08 | 6.08E-06 |
| rs157934 | C | T | LOC646329 | 7 | 0.003 | 0.005 | 0.010 | 0.000 | 0.001 | 0.002 | 2.20E-12 | 4.65E-12 | 1.61E-07 |
| rs16844249 | T | C | RGS12 | 7 | -0.005 | -0.006 | -0.009 | 0.001 | 0.001 | 0.003 | 1.10E-11 | 6.30E-09 | 3.13E-04 |
| rs16844401 | A | G | HGFAC | 7 | -0.009 | -0.009 | -0.018 | 0.001 | 0.001 | 0.003 | 2.39E-24 | 1.01E-12 | 1.13E-07 |
| rs16871026 | A | G | TAP1_PSMB9 | 7 | -0.008 | -0.012 | -0.030 | 0.002 | 0.002 | 0.006 | 3.21E-07 | 2.62E-08 | 6.54E-07 |
| rs17007241 | C | A | LOC285419 | 7 | -0.005 | -0.006 | -0.012 | 0.001 | 0.001 | 0.003 | 5.70E-11 | 3.05E-08 | 5.70E-06 |
| rs17123728 | A | G | DOCK7_ANGPTL3 | 7 | -0.005 | -0.009 | -0.016 | 0.001 | 0.002 | 0.004 | 1.39E-05 | 2.26E-08 | 2.23E-04 |
| rs17369400 | G | A | TRIM2 | 7 | 0.003 | 0.005 | 0.013 | 0.001 | 0.001 | 0.003 | 1.04E-05 | 2.83E-06 | 4.99E-06 |
| rs17377148 | G | T | NFIA | 7 | 0.007 | 0.006 | 0.014 | 0.001 | 0.001 | 0.003 | 3.57E-14 | 2.17E-07 | 3.41E-05 |
| rs17405319 | T | C | TRIB1 | 7 | -0.010 | -0.012 | -0.024 | 0.001 | 0.001 | 0.002 | 4.04E-58 | 7.85E-46 | 3.57E-26 |
| rs213487 | A | G | SSBP3 | 7 | 0.002 | 0.004 | 0.006 | 0.000 | 0.001 | 0.002 | 4.51E-07 | 1.06E-09 | 4.39E-04 |
| rs2244579 | C | G | HCP5_HCG26 | 7 | -0.004 | -0.006 | -0.010 | 0.001 | 0.001 | 0.002 | 6.55E-17 | 1.20E-17 | 3.63E-07 |
| rs2280401 | A | G | RPS11 | 7 | 0.006 | 0.005 | 0.012 | 0.001 | 0.001 | 0.002 | 2.46E-19 | 1.12E-09 | 7.50E-07 |
| rs2290505 | G | A | LLGL1 | 7 | 0.003 | 0.004 | 0.006 | 0.001 | 0.001 | 0.002 | 2.29E-09 | 3.50E-06 | 3.17E-03 |
| rs2292572 | T | G | GAB2 | 7 | 0.002 | 0.004 | 0.009 | 0.001 | 0.001 | 0.002 | 5.29E-03 | 5.80E-06 | 1.74E-04 |
| rs23544 | T | C | HLA-DMB | 7 | -0.002 | -0.003 | -0.005 | 0.000 | 0.001 | 0.002 | 8.28E-07 | 1.13E-07 | 4.58E-03 |
| rs2358087 | G | A | ZAK_MLK7-AS1 | 7 | 0.003 | 0.004 | 0.007 | 0.001 | 0.001 | 0.002 | 2.00E-08 | 2.60E-06 | 6.60E-04 |
| rs241429 | G | A | TAP2 | 7 | -0.003 | -0.005 | -0.010 | 0.000 | 0.001 | 0.002 | 1.03E-10 | 2.57E-16 | 2.17E-09 |
| rs2424904 | C | T | DNMT3B | 7 | -0.002 | -0.004 | -0.006 | 0.000 | 0.001 | 0.002 | 7.00E-05 | 3.09E-08 | 1.04E-03 |
| rs2488062 | C | T | EXOC6_HHEX | 7 | -0.003 | -0.005 | -0.009 | 0.001 | 0.001 | 0.002 | 3.09E-09 | 7.39E-10 | 6.54E-06 |
| rs2490741 | A | G | EXOC6 | 7 | -0.004 | -0.005 | -0.009 | 0.000 | 0.001 | 0.002 | 6.61E-18 | 2.51E-12 | 1.53E-06 |
| rs2586886 | T | C | KCNK3 | 7 | -0.003 | -0.004 | -0.010 | 0.000 | 0.001 | 0.002 | 1.01E-09 | 5.44E-10 | 9.90E-09 |
| rs2699436 | T | C | DOK7 | 7 | -0.005 | -0.006 | -0.013 | 0.001 | 0.001 | 0.003 | 5.78E-12 | 8.24E-09 | 6.33E-07 |
| rs2797581 | T | C | MYOF | 7 | -0.003 | -0.005 | -0.010 | 0.001 | 0.001 | 0.002 | 4.98E-07 | 6.35E-08 | 3.47E-05 |
| rs284279 | C | T | CASZ1 | 7 | -0.003 | -0.005 | -0.012 | 0.000 | 0.001 | 0.002 | 2.81E-12 | 1.13E-13 | 4.53E-12 |
| rs2844545 | A | G | HLA-B_MICA | 7 | 0.003 | 0.007 | 0.012 | 0.000 | 0.001 | 0.002 | 1.13E-09 | 6.38E-25 | 2.00E-11 |
| rs28579719 | G | A | TRIB1_LINC00861 | 7 | 0.005 | 0.006 | 0.009 | 0.000 | 0.001 | 0.002 | 1.69E-27 | 8.57E-20 | 5.65E-08 |
| rs2862954 | C | T | ERLIN1 | 7 | -0.001 | -0.004 | -0.008 | 0.000 | 0.001 | 0.002 | 3.94E-03 | 1.73E-09 | 4.52E-06 |
| rs2954006 | G | C | NSMCE2_TRIB1 | 7 | 0.003 | 0.003 | 0.005 | 0.000 | 0.001 | 0.002 | 2.08E-10 | 6.69E-07 | 1.87E-03 |
| rs310762 | C | T | SYN2 | 7 | -0.003 | -0.003 | -0.007 | 0.000 | 0.001 | 0.002 | 3.99E-10 | 1.20E-07 | 5.45E-05 |
| rs3129948 | A | C | C6orf10_HCG23 | 7 | -0.004 | -0.004 | -0.007 | 0.001 | 0.001 | 0.002 | 9.55E-13 | 4.57E-09 | 6.29E-04 |
| rs3135146 | G | C | RGS12_MSANTD1 | 7 | -0.003 | -0.003 | -0.004 | 0.000 | 0.001 | 0.002 | 6.25E-11 | 1.53E-05 | 3.09E-02 |
| rs314246 | C | A | ASGR2_ASGR1 | 7 | -0.004 | -0.004 | -0.010 | 0.001 | 0.001 | 0.002 | 4.72E-13 | 3.37E-06 | 2.21E-05 |
| rs34175245 | G | A | RPS6KA1_ARID1A | 7 | -0.010 | -0.007 | -0.014 | 0.001 | 0.001 | 0.003 | 4.48E-32 | 2.65E-09 | 8.73E-06 |
| rs34308086 | GA | G | PTK2 | 7 | -0.003 | -0.003 | -0.009 | 0.000 | 0.001 | 0.002 | 5.83E-09 | 6.00E-08 | 1.84E-07 |
| rs34668587 | G | A | MYT1_NPBWR2 | 7 | 0.003 | 0.003 | 0.005 | 0.001 | 0.001 | 0.002 | 7.54E-11 | 5.09E-05 | 7.56E-03 |
| rs3751120 | T | C | FLRT1_MACROD1 | 7 | -0.007 | -0.008 | -0.009 | 0.001 | 0.001 | 0.003 | 8.41E-18 | 1.15E-11 | 5.13E-03 |
| rs375244 | A | G | NOTCH4 | 7 | -0.003 | -0.005 | -0.010 | 0.001 | 0.001 | 0.002 | 1.15E-07 | 2.68E-10 | 3.72E-07 |
| rs3764400 | C | T | NFE2L1_COPZ2 | 7 | 0.004 | 0.005 | 0.006 | 0.001 | 0.001 | 0.003 | 2.00E-07 | 1.16E-06 | 1.21E-02 |
| rs3810068 | C | T | EMILIN2_SMCHD1 | 7 | 0.003 | 0.003 | 0.006 | 0.000 | 0.001 | 0.002 | 1.12E-09 | 2.11E-06 | 1.16E-03 |
| rs3823325 | C | G | HLA-A | 7 | -0.006 | -0.007 | -0.012 | 0.001 | 0.001 | 0.004 | 5.69E-09 | 5.91E-07 | 1.55E-03 |
| rs388234 | C | T | GABBR1_UBD | 7 | -0.002 | -0.004 | -0.007 | 0.000 | 0.001 | 0.002 | 2.93E-06 | 5.91E-08 | 5.84E-05 |
| rs4006563 | T | C | TRIB1_LINC00861 | 7 | 0.011 | 0.011 | 0.021 | 0.001 | 0.001 | 0.003 | 3.62E-37 | 1.62E-21 | 9.19E-12 |
| rs411326 | T | C | NOTCH4_C6orf10 | 7 | -0.004 | -0.005 | -0.012 | 0.001 | 0.001 | 0.002 | 4.02E-13 | 6.83E-14 | 3.58E-10 |
| rs4135268 | G | C | PPARG | 7 | -0.005 | -0.005 | -0.010 | 0.001 | 0.001 | 0.003 | 2.45E-09 | 1.64E-06 | 1.38E-03 |
| rs41780 | G | A | CAPZA2_MET | 7 | 0.003 | 0.002 | 0.004 | 0.000 | 0.001 | 0.002 | 9.43E-09 | 2.67E-04 | 1.33E-02 |
| rs4328458 | A | G | LINC00917_FENDRR | 7 | -0.003 | -0.004 | -0.009 | 0.000 | 0.001 | 0.002 | 1.86E-09 | 9.94E-12 | 6.56E-08 |
| rs440090 | T | C | C20orf196 | 7 | -0.004 | -0.004 | -0.010 | 0.001 | 0.001 | 0.002 | 1.40E-12 | 2.25E-07 | 4.57E-07 |
| rs4410790 | C | T | AHR_AGR3 | 7 | -0.002 | -0.004 | -0.006 | 0.000 | 0.001 | 0.002 | 3.27E-07 | 6.93E-08 | 1.67E-03 |
| rs45512696 | T | C | HPN_HPN-AS1 | 7 | -0.004 | -0.005 | -0.012 | 0.001 | 0.001 | 0.002 | 2.62E-10 | 8.91E-10 | 3.92E-07 |
| rs4632263 | T | C | SLC25A42 | 7 | 0.002 | 0.004 | 0.005 | 0.001 | 0.001 | 0.002 | 1.54E-05 | 7.31E-07 | 1.02E-02 |
| rs4663335 | A | G | MROH2A_UGT1A6 | 7 | -0.003 | -0.004 | -0.008 | 0.001 | 0.001 | 0.002 | 1.03E-07 | 4.05E-07 | 9.45E-04 |
| rs4918664 | G | A | CYP26A1_MYOF | 7 | 0.004 | 0.004 | 0.009 | 0.001 | 0.001 | 0.003 | 8.93E-08 | 4.18E-06 | 8.90E-04 |
| rs55637010 | A | C | TRIB1 | 7 | 0.008 | 0.010 | 0.015 | 0.001 | 0.001 | 0.004 | 9.10E-18 | 1.06E-12 | 2.96E-05 |
| rs55709232 | G | C | TRIB1 | 7 | -0.007 | -0.008 | -0.015 | 0.001 | 0.001 | 0.002 | 3.00E-40 | 5.81E-32 | 1.54E-14 |
| rs56120442 | C | G | DOCK7 | 7 | 0.008 | 0.016 | 0.033 | 0.001 | 0.002 | 0.005 | 1.23E-08 | 7.22E-15 | 2.22E-09 |
| rs56208677 | T | C | CETP | 7 | 0.009 | 0.009 | 0.023 | 0.001 | 0.001 | 0.003 | 3.05E-24 | 2.69E-13 | 4.95E-11 |
| rs57587560 | A | G | DOK7 | 7 | -0.004 | -0.005 | -0.008 | 0.001 | 0.001 | 0.002 | 2.07E-13 | 5.96E-11 | 4.37E-05 |
| rs58727484 | T | G | LYPLAL1_RNU5F-1 | 7 | 0.003 | 0.004 | 0.004 | 0.001 | 0.001 | 0.002 | 2.01E-05 | 4.05E-05 | 1.19E-01 |
| rs6065904 | A | G | PLTP | 7 | -0.011 | -0.008 | -0.014 | 0.001 | 0.001 | 0.002 | 1.35E-82 | 1.01E-22 | 1.08E-11 |
| rs6070139 | G | A | CTCFL_PCK1 | 7 | -0.002 | -0.004 | -0.007 | 0.000 | 0.001 | 0.002 | 1.73E-06 | 1.24E-09 | 1.29E-04 |
| rs6085202 | G | A | GPCPD1 | 7 | 0.003 | 0.003 | 0.007 | 0.000 | 0.001 | 0.002 | 1.07E-12 | 4.67E-07 | 4.68E-05 |
| rs6139748 | T | C | LINC00654 | 7 | 0.004 | 0.003 | 0.008 | 0.001 | 0.001 | 0.002 | 4.93E-12 | 8.91E-06 | 1.14E-04 |
| rs62372052 | G | A | GHR_CCDC152 | 7 | 0.004 | 0.005 | 0.009 | 0.001 | 0.001 | 0.003 | 8.55E-09 | 1.92E-05 | 8.96E-04 |
| rs62521592 | T | C | TRIB1_LINC00861 | 7 | 0.011 | 0.014 | 0.029 | 0.001 | 0.001 | 0.004 | 1.13E-27 | 5.97E-22 | 1.30E-13 |
| rs62523860 | A | G | TRIB1_LINC00861 | 7 | 0.014 | 0.017 | 0.031 | 0.002 | 0.002 | 0.006 | 2.89E-21 | 3.44E-15 | 5.02E-08 |
| rs625524 | A | G | APOA5_APOA4 | 7 | 0.007 | 0.011 | 0.013 | 0.001 | 0.002 | 0.005 | 5.94E-07 | 4.43E-09 | 8.88E-03 |
| rs645040 | T | G | PCCB_MSL2 | 7 | -0.005 | -0.006 | -0.008 | 0.001 | 0.001 | 0.002 | 1.72E-24 | 2.16E-13 | 9.34E-05 |
| rs6499862 | A | G | CETP_HERPUD1 | 7 | -0.006 | -0.006 | -0.013 | 0.001 | 0.001 | 0.002 | 6.33E-21 | 5.26E-11 | 6.55E-08 |
| rs6587971 | T | C | KANK4_USP1 | 7 | 0.004 | 0.007 | 0.011 | 0.000 | 0.001 | 0.002 | 1.54E-14 | 1.49E-24 | 7.56E-10 |
| rs663576 | T | C | FLJ40194 | 7 | 0.003 | 0.003 | 0.003 | 0.000 | 0.001 | 0.002 | 7.42E-10 | 1.65E-05 | 5.50E-02 |
| rs665268 | G | A | MLX | 7 | -0.003 | -0.003 | -0.005 | 0.001 | 0.001 | 0.002 | 6.49E-08 | 1.32E-05 | 5.46E-03 |
| rs6867983 | T | C | MAP3K1_ANKRD55 | 7 | -0.007 | -0.008 | -0.011 | 0.001 | 0.001 | 0.002 | 7.71E-24 | 6.65E-17 | 1.88E-05 |
| rs6983849 | C | T | NAT1_NAT2 | 7 | -0.003 | -0.005 | -0.006 | 0.001 | 0.001 | 0.002 | 1.90E-07 | 1.15E-08 | 1.10E-02 |
| rs7094463 | G | A | TCF7L2 | 7 | -0.003 | -0.004 | -0.009 | 0.000 | 0.001 | 0.002 | 6.31E-10 | 5.08E-09 | 5.09E-07 |
| rs72655677 | A | G | TRIB1_LINC00861 | 7 | 0.013 | 0.017 | 0.030 | 0.001 | 0.001 | 0.004 | 3.30E-34 | 1.67E-30 | 4.78E-14 |
| rs72801137 | C | T | SGCD | 7 | -0.009 | -0.014 | -0.026 | 0.001 | 0.002 | 0.005 | 8.03E-10 | 4.45E-12 | 6.69E-07 |
| rs73238159 | T | C | XRN1 | 7 | 0.004 | 0.005 | 0.012 | 0.001 | 0.001 | 0.003 | 3.25E-10 | 2.25E-08 | 5.33E-06 |
| rs738409 | G | C | PNPLA3 | 7 | 0.000 | 0.005 | 0.007 | 0.001 | 0.001 | 0.002 | 7.43E-01 | 4.34E-09 | 4.82E-04 |
| rs75978204 | G | A | KANK4_USP1 | 7 | 0.006 | 0.011 | 0.019 | 0.001 | 0.001 | 0.004 | 1.59E-08 | 4.69E-15 | 6.67E-07 |
| rs76191003 | T | C | SLC38A4 | 7 | -0.006 | -0.006 | -0.014 | 0.001 | 0.001 | 0.003 | 2.92E-10 | 3.74E-07 | 5.28E-05 |
| rs78044163 | G | A | ZSWIM3 | 7 | -0.008 | -0.006 | -0.013 | 0.001 | 0.002 | 0.004 | 5.15E-12 | 1.78E-04 | 2.49E-03 |
| rs78141093 | C | G | DNAJC5G | 7 | 0.008 | 0.013 | 0.021 | 0.002 | 0.002 | 0.006 | 2.25E-06 | 4.75E-08 | 9.54E-04 |
| rs78854340 | T | C | SGCD | 7 | 0.008 | 0.009 | 0.018 | 0.001 | 0.002 | 0.006 | 1.59E-07 | 8.06E-06 | 2.05E-03 |
| rs78960801 | A | C | APOB_C2orf43 | 7 | 0.004 | 0.005 | 0.005 | 0.001 | 0.001 | 0.004 | 1.51E-05 | 8.58E-04 | 2.10E-01 |
| rs79336077 | A | G | TRIB1_LINC00861 | 7 | 0.013 | 0.018 | 0.030 | 0.002 | 0.003 | 0.008 | 4.50E-11 | 4.92E-10 | 9.17E-05 |
| rs79429216 | A | G | APOC2_APOC4 | 7 | -0.028 | -0.034 | -0.072 | 0.002 | 0.003 | 0.008 | 5.67E-38 | 4.13E-33 | 1.76E-20 |
| rs79617217 | G | C | C2orf43 | 7 | -0.008 | -0.006 | -0.012 | 0.001 | 0.001 | 0.004 | 8.26E-14 | 7.34E-05 | 2.14E-03 |
| rs80265946 | A | C | HCAR1_KNTC1 | 7 | -0.006 | -0.008 | -0.013 | 0.001 | 0.002 | 0.004 | 1.49E-08 | 1.08E-06 | 2.38E-03 |
| rs9258740 | T | C | HLA-G_HLA-H | 7 | -0.003 | -0.004 | -0.009 | 0.001 | 0.001 | 0.002 | 1.08E-07 | 7.37E-08 | 2.76E-05 |
| rs9267547 | A | G | LY6G6F_LY6G6E | 7 | -0.003 | -0.006 | -0.009 | 0.001 | 0.001 | 0.003 | 5.49E-05 | 4.88E-07 | 3.26E-03 |
| rs9332817 | C | G | KMT2A | 7 | 0.006 | 0.010 | 0.014 | 0.001 | 0.002 | 0.005 | 8.91E-05 | 2.45E-06 | 1.13E-02 |
| rs9568410 | A | G | DLEU1_DLEU7 | 7 | -0.005 | -0.006 | -0.011 | 0.001 | 0.001 | 0.003 | 2.78E-11 | 7.13E-11 | 4.61E-05 |
| rs9577896 | G | A | TMEM255B | 7 | -0.003 | -0.004 | -0.009 | 0.000 | 0.001 | 0.002 | 6.11E-09 | 5.06E-10 | 2.36E-07 |
| rs9604527 | T | C | TMEM255B | 7 | -0.003 | -0.005 | -0.009 | 0.001 | 0.001 | 0.002 | 7.85E-08 | 7.52E-08 | 8.81E-05 |
| rs9653547 | A | C | AHSA2 | 7 | 0.004 | 0.005 | 0.009 | 0.001 | 0.001 | 0.002 | 3.55E-09 | 1.95E-09 | 2.36E-04 |
| rs9940128 | A | G | FTO | 7 | 0.003 | 0.005 | 0.012 | 0.000 | 0.001 | 0.002 | 2.33E-08 | 3.98E-16 | 1.52E-12 |
| rs10176110 | C | T | FLJ31356_BRE | 8 | -0.002 | -0.004 | -0.002 | 0.001 | 0.001 | 0.003 | 7.32E-04 | 2.94E-06 | 3.83E-01 |
| rs1038694 | G | A | BRE | 8 | -0.004 | -0.006 | -0.008 | 0.001 | 0.001 | 0.002 | 5.99E-14 | 5.29E-17 | 3.77E-05 |
| rs10426036 | A | G | CERS4_FBN3 | 8 | -0.002 | -0.002 | -0.003 | 0.001 | 0.001 | 0.002 | 1.73E-04 | 5.85E-04 | 1.16E-01 |
| rs10497251 | C | T | SLC38A11_COBLL1 | 8 | -0.003 | -0.004 | -0.005 | 0.001 | 0.001 | 0.002 | 7.81E-08 | 3.25E-08 | 1.66E-02 |
| rs10516788 | G | A | AFF1 | 8 | -0.003 | -0.007 | -0.007 | 0.001 | 0.001 | 0.002 | 1.29E-06 | 3.23E-16 | 1.04E-03 |
| rs10773048 | C | T | CCDC92 | 8 | -0.003 | -0.004 | -0.004 | 0.000 | 0.001 | 0.002 | 3.90E-11 | 2.37E-08 | 1.06E-02 |
| rs11054397 | A | G | ETV6_LOC338817 | 8 | -0.002 | -0.003 | -0.004 | 0.000 | 0.001 | 0.002 | 8.51E-05 | 1.76E-06 | 1.96E-02 |
| rs11057353 | C | T | DNAH10 | 8 | 0.004 | 0.006 | 0.009 | 0.000 | 0.001 | 0.002 | 3.88E-19 | 6.51E-20 | 1.47E-07 |
| rs111712352 | T | C | USP40 | 8 | -0.007 | -0.010 | -0.017 | 0.001 | 0.002 | 0.004 | 2.37E-09 | 1.51E-10 | 1.56E-04 |
| rs111981233 | G | T | FCGRT | 8 | -0.005 | -0.006 | -0.010 | 0.001 | 0.001 | 0.003 | 7.37E-09 | 1.10E-07 | 1.28E-03 |
| rs112222843 | T | C | VEGFA_LOC100132354 | 8 | 0.006 | 0.008 | 0.007 | 0.001 | 0.002 | 0.005 | 6.71E-06 | 4.60E-06 | 1.39E-01 |
| rs115268969 | T | C | DPYSL5_MAPRE3 | 8 | -0.008 | -0.012 | -0.013 | 0.002 | 0.002 | 0.006 | 5.58E-06 | 1.64E-07 | 3.48E-02 |
| rs115984680 | T | C | MPV17 | 8 | -0.007 | -0.012 | -0.010 | 0.002 | 0.003 | 0.007 | 5.24E-04 | 6.01E-06 | 1.74E-01 |
| rs117291242 | T | C | B4GALNT4 | 8 | -0.004 | -0.010 | -0.013 | 0.001 | 0.002 | 0.005 | 6.13E-04 | 4.12E-08 | 5.33E-03 |
| rs117528284 | A | G | BAZ1B | 8 | -0.005 | -0.008 | -0.006 | 0.001 | 0.002 | 0.005 | 6.64E-06 | 2.57E-06 | 1.77E-01 |
| rs11902458 | T | C | BRE | 8 | 0.003 | 0.003 | 0.005 | 0.000 | 0.001 | 0.002 | 1.80E-08 | 8.23E-07 | 8.41E-03 |
| rs11949724 | A | G | MAP3K1_ANKRD55 | 8 | -0.002 | -0.003 | -0.005 | 0.000 | 0.001 | 0.002 | 2.48E-06 | 1.19E-06 | 1.02E-02 |
| rs12145455 | C | T | KANK4_USP1 | 8 | 0.006 | 0.013 | 0.021 | 0.001 | 0.001 | 0.003 | 7.49E-16 | 3.10E-33 | 2.77E-12 |
| rs12804297 | A | G | SIK3 | 8 | 0.003 | 0.005 | 0.005 | 0.001 | 0.001 | 0.003 | 1.17E-04 | 1.93E-04 | 1.55E-01 |
| rs13250456 | T | C | EYA1_MSC | 8 | -0.005 | -0.008 | -0.009 | 0.001 | 0.001 | 0.003 | 5.68E-10 | 2.10E-09 | 4.76E-03 |
| rs13273592 | C | T | TRIB1_LINC00861 | 8 | -0.004 | -0.006 | -0.008 | 0.001 | 0.001 | 0.002 | 1.87E-13 | 2.01E-19 | 5.24E-05 |
| rs1493042 | C | A | NAT1_NAT2 | 8 | 0.003 | 0.005 | 0.007 | 0.001 | 0.001 | 0.002 | 3.61E-06 | 9.93E-10 | 8.25E-04 |
| rs1609574 | C | T | MYOF | 8 | -0.006 | -0.008 | -0.017 | 0.001 | 0.002 | 0.005 | 4.86E-07 | 9.01E-07 | 2.05E-04 |
| rs1647280 | G | C | PPM1G | 8 | 0.008 | 0.010 | 0.021 | 0.001 | 0.002 | 0.005 | 6.92E-09 | 1.62E-07 | 3.27E-05 |
| rs1716403 | C | T | ZNF664-FAM101A | 8 | -0.005 | -0.006 | -0.008 | 0.000 | 0.001 | 0.002 | 3.54E-25 | 1.96E-18 | 1.71E-05 |
| rs17184382 | C | A | USP3_CA12 | 8 | 0.003 | 0.004 | 0.004 | 0.000 | 0.001 | 0.002 | 6.85E-09 | 2.81E-08 | 1.78E-02 |
| rs17449582 | T | C | MAPK10 | 8 | -0.002 | -0.005 | -0.005 | 0.000 | 0.001 | 0.002 | 7.39E-07 | 4.47E-14 | 4.85E-03 |
| rs17734320 | T | C | IQCK | 8 | -0.003 | -0.005 | -0.005 | 0.001 | 0.001 | 0.003 | 1.81E-04 | 1.96E-05 | 1.00E-01 |
| rs1931081 | T | C | ATG4C_LINC00466 | 8 | -0.002 | -0.004 | -0.004 | 0.000 | 0.001 | 0.002 | 1.64E-04 | 3.29E-09 | 1.33E-02 |
| rs2075260 | A | G | ACACB | 8 | -0.003 | -0.004 | -0.008 | 0.001 | 0.001 | 0.002 | 1.84E-07 | 4.92E-07 | 2.92E-04 |
| rs2306374 | C | T | MRAS | 8 | -0.003 | -0.003 | 0.001 | 0.001 | 0.001 | 0.002 | 1.79E-05 | 5.81E-04 | 7.15E-01 |
| rs2306719 | C | T | CA12 | 8 | -0.004 | -0.006 | -0.006 | 0.001 | 0.001 | 0.004 | 1.60E-04 | 5.39E-06 | 9.49E-02 |
| rs234051 | G | A | NCEH1_TNFSF10 | 8 | -0.002 | -0.004 | -0.006 | 0.000 | 0.001 | 0.002 | 1.66E-06 | 3.58E-07 | 3.10E-03 |
| rs2442728 | T | G | HLA-B_HLA-C | 8 | 0.002 | 0.005 | 0.006 | 0.000 | 0.001 | 0.002 | 4.90E-05 | 4.05E-13 | 2.43E-04 |
| rs2534686 | C | T | HCP5_MICB | 8 | -0.003 | -0.006 | -0.007 | 0.001 | 0.001 | 0.002 | 3.53E-08 | 5.17E-11 | 1.13E-03 |
| rs2657888 | G | T | RBMS2 | 8 | -0.002 | -0.003 | -0.004 | 0.000 | 0.001 | 0.002 | 4.57E-06 | 1.10E-05 | 1.62E-02 |
| rs2803240 | C | A | ATG4C_LINC00466 | 8 | -0.005 | -0.010 | -0.013 | 0.001 | 0.001 | 0.003 | 2.97E-08 | 1.18E-18 | 5.52E-05 |
| rs283174 | T | G | MAP3K1_ANKRD55 | 8 | -0.004 | -0.008 | -0.006 | 0.002 | 0.003 | 0.007 | 1.71E-02 | 8.56E-04 | 3.85E-01 |
| rs28489942 | C | T | TCF23_C2orf53 | 8 | 0.003 | 0.006 | 0.006 | 0.000 | 0.001 | 0.002 | 2.30E-10 | 3.20E-16 | 3.47E-04 |
| rs2869441 | G | A | MAPK10_PTPN13 | 8 | 0.003 | 0.005 | 0.004 | 0.001 | 0.001 | 0.003 | 1.10E-04 | 4.99E-07 | 1.12E-01 |
| rs2971677 | A | C | GCK | 8 | -0.004 | -0.006 | -0.009 | 0.001 | 0.001 | 0.002 | 6.63E-12 | 1.49E-10 | 1.96E-04 |
| rs3025053 | A | G | VEGFA | 8 | 0.004 | 0.005 | 0.005 | 0.001 | 0.001 | 0.003 | 1.90E-08 | 1.15E-07 | 6.53E-02 |
| rs3093664 | G | A | TNF | 8 | -0.002 | -0.007 | -0.011 | 0.001 | 0.001 | 0.003 | 3.35E-02 | 6.60E-09 | 4.24E-04 |
| rs3130980 | T | C | C6orf15_PSORS1C1 | 8 | -0.002 | -0.005 | -0.005 | 0.000 | 0.001 | 0.002 | 2.70E-05 | 2.56E-14 | 8.14E-03 |
| rs3132584 | T | G | TUBB | 8 | 0.003 | 0.006 | 0.009 | 0.001 | 0.001 | 0.002 | 3.74E-07 | 2.86E-14 | 1.00E-05 |
| rs3134608 | G | T | PRRT1 | 8 | 0.003 | 0.006 | 0.009 | 0.001 | 0.001 | 0.002 | 3.59E-10 | 3.44E-13 | 1.76E-05 |
| rs337638 | C | T | FLJ13197_TBC1D1 | 8 | -0.002 | -0.003 | -0.006 | 0.000 | 0.001 | 0.002 | 2.55E-04 | 5.15E-06 | 1.82E-03 |
| rs34340041 | T | G | APOB_LOC645949 | 8 | -0.006 | -0.006 | -0.007 | 0.001 | 0.001 | 0.003 | 1.29E-13 | 1.31E-08 | 1.32E-02 |
| rs34370554 | G | T | NAT2_PSD3 | 8 | -0.004 | -0.007 | -0.010 | 0.001 | 0.001 | 0.002 | 9.78E-16 | 2.30E-24 | 7.80E-07 |
| rs34822542 | C | T | PREX1 | 8 | -0.002 | -0.004 | -0.003 | 0.001 | 0.001 | 0.002 | 4.79E-04 | 7.15E-05 | 1.60E-01 |
| rs352159 | C | T | POC1A_ALAS1 | 8 | 0.006 | 0.008 | 0.015 | 0.001 | 0.002 | 0.004 | 3.45E-08 | 4.83E-08 | 1.38E-04 |
| rs35665085 | A | G | CECR5 | 8 | -0.005 | -0.007 | -0.006 | 0.001 | 0.001 | 0.004 | 1.70E-06 | 1.39E-06 | 1.39E-01 |
| rs35668759 | C | A | CYP26A1_MYOF | 8 | -0.004 | -0.006 | -0.007 | 0.001 | 0.001 | 0.002 | 5.13E-09 | 3.62E-12 | 5.23E-03 |
| rs35761929 | C | G | JAG1 | 8 | 0.003 | 0.006 | 0.008 | 0.001 | 0.001 | 0.003 | 2.25E-04 | 2.83E-06 | 2.76E-02 |
| rs3746503 | G | A | ZNF335 | 8 | 0.004 | 0.004 | 0.004 | 0.001 | 0.001 | 0.003 | 2.34E-09 | 1.96E-04 | 1.67E-01 |
| rs37530 | C | A | PLK2_ACTBL2 | 8 | 0.002 | 0.003 | 0.004 | 0.000 | 0.001 | 0.002 | 9.19E-05 | 3.44E-05 | 1.28E-02 |
| rs3809114 | A | G | INHBE_INHBC | 8 | 0.004 | 0.005 | 0.006 | 0.000 | 0.001 | 0.002 | 7.01E-17 | 3.27E-13 | 1.84E-03 |
| rs3889571 | T | G | DSCAML1 | 8 | 0.001 | 0.004 | 0.005 | 0.000 | 0.001 | 0.002 | 1.57E-02 | 3.94E-08 | 5.10E-03 |
| rs3923113 | C | A | GRB14_COBLL1 | 8 | 0.005 | 0.008 | 0.008 | 0.000 | 0.001 | 0.002 | 6.08E-26 | 4.53E-34 | 1.22E-05 |
| rs3926666 | T | C | ZNF664-FAM101A | 8 | 0.003 | 0.004 | 0.006 | 0.000 | 0.001 | 0.002 | 1.30E-08 | 1.02E-07 | 1.59E-03 |
| rs41288817 | A | G | CAD | 8 | 0.005 | 0.011 | 0.010 | 0.002 | 0.003 | 0.007 | 5.49E-03 | 3.41E-05 | 1.39E-01 |
| rs41290100 | T | C | PVRL2 | 8 | 0.009 | 0.015 | 0.020 | 0.002 | 0.002 | 0.006 | 1.68E-08 | 2.32E-14 | 3.25E-04 |
| rs4151121 | G | A | FGF11_TMEM102 | 8 | 0.002 | 0.004 | 0.007 | 0.000 | 0.001 | 0.002 | 4.60E-07 | 1.84E-09 | 1.79E-04 |
| rs42124 | A | G | FZD9_FKBP6 | 8 | -0.013 | -0.019 | -0.021 | 0.001 | 0.002 | 0.005 | 2.29E-21 | 3.59E-22 | 4.25E-05 |
| rs4244455 | T | G | CSGALNACT1 | 8 | 0.003 | 0.006 | 0.004 | 0.001 | 0.001 | 0.003 | 8.56E-05 | 5.65E-09 | 1.25E-01 |
| rs4450871 | G | A | CYTL1_MSX1 | 8 | 0.002 | 0.003 | 0.004 | 0.000 | 0.001 | 0.002 | 7.61E-08 | 2.44E-07 | 2.69E-02 |
| rs45449995 | G | A | UGT1A6_UGT1A10 | 8 | 0.009 | 0.016 | 0.025 | 0.001 | 0.002 | 0.005 | 1.35E-10 | 1.42E-16 | 8.37E-07 |
| rs46522 | T | C | UBE2Z | 8 | -0.002 | -0.003 | -0.003 | 0.000 | 0.001 | 0.002 | 8.94E-07 | 6.52E-05 | 1.28E-01 |
| rs4665347 | A | C | CENPA | 8 | 0.003 | 0.005 | 0.008 | 0.001 | 0.001 | 0.002 | 9.91E-07 | 1.96E-13 | 3.46E-05 |
| rs4731702 | T | C | KLF14_MIR29A | 8 | 0.005 | 0.007 | 0.006 | 0.000 | 0.001 | 0.002 | 5.52E-28 | 1.58E-31 | 4.22E-04 |
| rs4802307 | T | G | PPP5C_HIF3A | 8 | 0.002 | 0.004 | 0.004 | 0.001 | 0.001 | 0.002 | 8.62E-04 | 2.15E-08 | 4.16E-02 |
| rs4806073 | C | T | HPN_HPN-AS1 | 8 | -0.005 | -0.006 | -0.010 | 0.001 | 0.001 | 0.003 | 1.50E-07 | 1.32E-07 | 1.96E-03 |
| rs4871601 | G | A | TRIB1_LINC00861 | 8 | -0.009 | -0.015 | -0.018 | 0.001 | 0.001 | 0.004 | 1.14E-19 | 7.23E-28 | 3.49E-07 |
| rs4915833 | G | A | KANK4_USP1 | 8 | 0.003 | 0.007 | 0.009 | 0.001 | 0.001 | 0.003 | 2.65E-05 | 1.92E-13 | 4.76E-04 |
| rs495033 | T | C | BUD13_LINC00900 | 8 | 0.004 | 0.009 | 0.011 | 0.001 | 0.001 | 0.003 | 1.37E-06 | 3.45E-12 | 6.31E-04 |
| rs55981065 | A | G | MYO1A | 8 | 0.010 | 0.015 | 0.023 | 0.002 | 0.003 | 0.007 | 3.47E-08 | 2.36E-08 | 1.20E-03 |
| rs56091605 | T | C | RBKS_MRPL33 | 8 | 0.007 | 0.011 | 0.014 | 0.001 | 0.002 | 0.006 | 3.04E-06 | 8.91E-08 | 1.16E-02 |
| rs5767634 | A | G | PPARA | 8 | -0.002 | -0.003 | -0.003 | 0.001 | 0.001 | 0.002 | 3.67E-04 | 5.12E-05 | 1.23E-01 |
| rs58053084 | G | T | GCK_YKT6 | 8 | 0.004 | 0.007 | 0.006 | 0.001 | 0.001 | 0.003 | 3.06E-06 | 1.01E-07 | 5.25E-02 |
| rs58199976 | C | T | LOC100129995_SLC4A1AP | 8 | 0.007 | 0.012 | 0.017 | 0.001 | 0.001 | 0.002 | 2.02E-38 | 3.02E-60 | 8.40E-18 |
| rs598503 | C | T | SIK3 | 8 | -0.009 | -0.013 | -0.021 | 0.002 | 0.002 | 0.006 | 1.42E-07 | 1.47E-08 | 5.66E-04 |
| rs6063048 | A | G | EYA2 | 8 | 0.003 | 0.004 | 0.004 | 0.001 | 0.001 | 0.002 | 1.35E-07 | 1.82E-09 | 5.16E-02 |
| rs61736639 | C | G | PDE3B | 8 | 0.015 | 0.020 | 0.026 | 0.003 | 0.004 | 0.011 | 6.17E-08 | 8.87E-07 | 1.36E-02 |
| rs61748245 | T | A | GRB14 | 8 | -0.003 | -0.004 | -0.003 | 0.000 | 0.001 | 0.002 | 9.37E-08 | 1.70E-08 | 1.34E-01 |
| rs61749613 | G | A | VCAN | 8 | 0.005 | 0.008 | 0.006 | 0.001 | 0.002 | 0.004 | 1.69E-05 | 3.11E-06 | 1.54E-01 |
| rs61884049 | T | C | COPB1_RRAS2 | 8 | 0.006 | 0.006 | 0.007 | 0.001 | 0.001 | 0.004 | 3.93E-08 | 2.58E-05 | 5.87E-02 |
| rs61904855 | A | C | BUD13_LINC00900 | 8 | -0.008 | -0.014 | -0.016 | 0.001 | 0.002 | 0.005 | 2.52E-09 | 2.33E-13 | 1.97E-03 |
| rs62075812 | T | C | ATP6V0A1 | 8 | 0.003 | 0.004 | 0.005 | 0.001 | 0.001 | 0.002 | 1.94E-05 | 2.49E-07 | 2.28E-02 |
| rs62129550 | A | G | KCNK3 | 8 | -0.006 | -0.012 | -0.022 | 0.001 | 0.002 | 0.004 | 1.94E-09 | 9.97E-15 | 1.17E-07 |
| rs62140389 | T | C | BRE_MRPL33 | 8 | -0.002 | -0.004 | -0.005 | 0.001 | 0.001 | 0.002 | 4.29E-04 | 3.38E-07 | 8.00E-03 |
| rs62521034 | T | C | TRIB1 | 8 | 0.004 | 0.006 | 0.006 | 0.001 | 0.001 | 0.002 | 3.16E-11 | 4.29E-13 | 4.79E-03 |
| rs6470368 | T | C | TRIB1_LINC00861 | 8 | -0.002 | -0.003 | -0.002 | 0.000 | 0.001 | 0.002 | 6.17E-07 | 2.86E-04 | 3.33E-01 |
| rs6488898 | A | G | ATP6V0A2 | 8 | 0.005 | 0.006 | 0.009 | 0.001 | 0.001 | 0.003 | 1.01E-08 | 5.73E-07 | 6.97E-03 |
| rs651837 | G | A | LOC645434_LOC100132735 | 8 | 0.004 | 0.006 | 0.007 | 0.000 | 0.001 | 0.002 | 1.39E-20 | 6.49E-23 | 1.11E-04 |
| rs6586720 | T | G | NAT1_NAT2 | 8 | 0.002 | 0.003 | 0.004 | 0.000 | 0.001 | 0.002 | 1.25E-04 | 2.31E-05 | 3.45E-02 |
| rs66485845 | C | CCA | CENPA | 8 | -0.006 | -0.010 | -0.012 | 0.001 | 0.001 | 0.003 | 1.99E-11 | 1.21E-16 | 2.49E-04 |
| rs67611724 | T | C | PXMP4_ZNF341 | 8 | -0.003 | -0.004 | -0.006 | 0.001 | 0.001 | 0.002 | 6.21E-08 | 1.68E-05 | 7.93E-03 |
| rs6893184 | A | G | TNFAIP8_HSD17B4 | 8 | 0.002 | 0.003 | 0.004 | 0.000 | 0.001 | 0.002 | 7.24E-07 | 9.93E-07 | 2.78E-02 |
| rs6919603 | G | A | HCG27_HLA-C | 8 | -0.003 | -0.007 | -0.006 | 0.001 | 0.001 | 0.002 | 2.01E-09 | 1.84E-18 | 1.51E-03 |
| rs693975 | A | G | CREB3L3_MAP2K2 | 8 | -0.002 | -0.004 | -0.003 | 0.001 | 0.001 | 0.002 | 8.88E-04 | 1.32E-07 | 9.00E-02 |
| rs6974424 | G | A | VPS37D_MLXIPL | 8 | -0.002 | -0.003 | -0.004 | 0.000 | 0.001 | 0.002 | 4.89E-06 | 9.30E-08 | 1.50E-02 |
| rs7081 | T | C | SLC5A6 | 8 | -0.009 | -0.014 | -0.019 | 0.001 | 0.001 | 0.004 | 2.94E-20 | 3.13E-26 | 7.38E-08 |
| rs71624136 | A | C | MAP3K1_ANKRD55 | 8 | -0.011 | -0.015 | -0.021 | 0.002 | 0.003 | 0.008 | 8.01E-08 | 3.48E-08 | 4.53E-03 |
| rs71643639 | C | T | DOCK7 | 8 | -0.004 | -0.007 | -0.011 | 0.001 | 0.001 | 0.004 | 2.65E-04 | 3.03E-07 | 3.36E-03 |
| rs72669514 | T | C | ATG4C_LOC400756 | 8 | -0.005 | -0.011 | -0.018 | 0.001 | 0.002 | 0.004 | 1.70E-05 | 6.47E-12 | 1.30E-05 |
| rs72800469 | A | G | KCNK3 | 8 | -0.007 | -0.008 | -0.014 | 0.001 | 0.002 | 0.005 | 1.04E-09 | 4.53E-07 | 2.48E-03 |
| rs72976106 | T | C | CENPW_RSPO3 | 8 | -0.008 | -0.011 | -0.018 | 0.001 | 0.002 | 0.005 | 8.99E-11 | 8.27E-11 | 6.33E-05 |
| rs73009178 | A | C | BUD13_LINC00900 | 8 | 0.002 | 0.007 | 0.009 | 0.001 | 0.001 | 0.003 | 2.90E-03 | 3.26E-11 | 6.06E-04 |
| rs73207887 | G | A | NAT2_PSD3 | 8 | -0.005 | -0.011 | -0.010 | 0.001 | 0.002 | 0.005 | 3.73E-05 | 9.20E-11 | 2.57E-02 |
| rs73243877 | G | A | SMIM20_RBPJ | 8 | -0.004 | -0.006 | -0.007 | 0.001 | 0.001 | 0.002 | 2.30E-11 | 7.61E-11 | 1.33E-03 |
| rs73579379 | A | G | LINC00452_LOC100506394 | 8 | 0.005 | 0.007 | 0.012 | 0.001 | 0.001 | 0.002 | 9.52E-13 | 5.86E-15 | 3.24E-06 |
| rs7419069 | C | G | DOCK7 | 8 | 0.004 | 0.009 | 0.013 | 0.001 | 0.001 | 0.004 | 2.87E-05 | 2.82E-11 | 1.83E-04 |
| rs74496689 | A | G | BUD13 | 8 | 0.015 | 0.024 | 0.032 | 0.002 | 0.002 | 0.007 | 1.02E-17 | 1.73E-21 | 1.20E-06 |
| rs75252677 | A | G | CAD_SLC30A3 | 8 | 0.010 | 0.018 | 0.026 | 0.002 | 0.002 | 0.006 | 1.18E-10 | 1.11E-16 | 8.98E-06 |
| rs75493897 | T | C | BCL7A | 8 | -0.003 | -0.004 | -0.006 | 0.001 | 0.001 | 0.003 | 1.78E-04 | 9.58E-04 | 8.46E-02 |
| rs76164683 | C | G | MPV17_GTF3C2 | 8 | -0.009 | -0.013 | -0.019 | 0.002 | 0.002 | 0.006 | 3.78E-07 | 2.02E-07 | 3.62E-03 |
| rs76599771 | A | G | TRIB1_LINC00861 | 8 | -0.009 | -0.012 | -0.015 | 0.002 | 0.002 | 0.007 | 1.33E-06 | 2.69E-06 | 2.55E-02 |
| rs76993561 | G | C | TRIB1_LINC00861 | 8 | -0.013 | -0.018 | -0.029 | 0.001 | 0.002 | 0.005 | 6.26E-25 | 1.32E-24 | 1.32E-09 |
| rs77925742 | T | C | SOAT2 | 8 | 0.005 | 0.002 | -0.001 | 0.001 | 0.001 | 0.003 | 1.25E-10 | 1.18E-01 | 6.90E-01 |
| rs78025076 | T | C | CCDC109B | 8 | -0.006 | -0.012 | -0.016 | 0.002 | 0.002 | 0.006 | 2.49E-04 | 3.37E-07 | 8.29E-03 |
| rs78287364 | G | A | DOCK7 | 8 | 0.007 | 0.014 | 0.016 | 0.001 | 0.002 | 0.004 | 7.78E-10 | 3.52E-18 | 4.18E-04 |
| rs78309295 | G | A | TRIB1_LINC00861 | 8 | -0.012 | -0.018 | -0.026 | 0.002 | 0.002 | 0.006 | 6.78E-14 | 1.93E-15 | 1.15E-05 |
| rs7832357 | G | A | TRIB1_LINC00861 | 8 | -0.007 | -0.010 | -0.015 | 0.000 | 0.001 | 0.002 | 5.19E-51 | 2.75E-53 | 1.20E-15 |
| rs7903146 | T | C | TCF7L2 | 8 | -0.003 | -0.001 | 0.008 | 0.001 | 0.001 | 0.002 | 3.34E-07 | 7.32E-02 | 6.39E-05 |
| rs7935656 | T | C | BUD13_LINC00900 | 8 | 0.001 | 0.003 | 0.004 | 0.000 | 0.001 | 0.002 | 4.41E-03 | 8.38E-06 | 2.90E-02 |
| rs7978567 | T | C | LRP1 | 8 | 0.003 | 0.003 | 0.005 | 0.000 | 0.001 | 0.002 | 3.96E-09 | 1.04E-06 | 1.01E-02 |
| rs8046999 | G | A | LMF1 | 8 | 0.002 | 0.003 | 0.005 | 0.000 | 0.001 | 0.002 | 2.39E-06 | 1.50E-06 | 1.24E-02 |
| rs881858 | A | G | VEGFA_LOC100132354 | 8 | -0.002 | -0.004 | -0.003 | 0.000 | 0.001 | 0.002 | 2.07E-05 | 6.01E-09 | 1.36E-01 |
| rs891088 | G | A | INSR | 8 | 0.002 | 0.002 | 0.002 | 0.001 | 0.001 | 0.002 | 1.95E-03 | 7.62E-04 | 2.27E-01 |
| rs9276909 | T | C | PPP1R2P1_PSMB9 | 8 | 0.002 | 0.005 | 0.006 | 0.001 | 0.001 | 0.002 | 2.12E-05 | 1.05E-12 | 2.50E-03 |
| rs9368716 | A | G | C6orf10 | 8 | -0.003 | -0.004 | -0.004 | 0.000 | 0.001 | 0.002 | 8.74E-10 | 4.99E-11 | 2.39E-02 |
| rs9394047 | C | A | HLA-C_HCG27 | 8 | -0.002 | -0.004 | -0.007 | 0.001 | 0.001 | 0.003 | 4.51E-03 | 4.74E-04 | 2.44E-02 |
| rs9425291 | A | G | DNM3 | 8 | -0.003 | -0.003 | -0.004 | 0.000 | 0.001 | 0.002 | 3.32E-09 | 1.30E-07 | 3.65E-02 |
| rs9697415 | G | A | RILPL1 | 8 | -0.002 | -0.003 | -0.003 | 0.001 | 0.001 | 0.002 | 5.52E-05 | 8.23E-04 | 1.41E-01 |
| rs972275 | G | C | CENPW_RSPO3 | 8 | 0.003 | 0.004 | 0.006 | 0.000 | 0.001 | 0.002 | 1.01E-09 | 4.80E-10 | 2.01E-04 |
| rs1008953 | C | T | SYS1_SDC4 | 9 | 0.002 | 0.002 | 0.003 | 0.001 | 0.001 | 0.002 | 2.44E-04 | 2.75E-03 | 1.01E-01 |
| rs10194882 | A | G | MIR5702_LOC646736 | 9 | -0.003 | -0.005 | 0.001 | 0.000 | 0.001 | 0.002 | 8.90E-11 | 9.71E-14 | 4.93E-01 |
| rs10445598 | G | C | CEBPG_CEBPA-AS1 | 9 | 0.002 | 0.003 | 0.004 | 0.000 | 0.001 | 0.002 | 1.06E-04 | 2.39E-07 | 2.92E-02 |
| rs10466531 | T | C | BUD13_LINC00900 | 9 | -0.001 | -0.004 | -0.005 | 0.000 | 0.001 | 0.002 | 7.46E-03 | 3.50E-08 | 6.72E-03 |
| rs1049817 | G | A | GTF3C2 | 9 | 0.007 | 0.012 | 0.013 | 0.000 | 0.001 | 0.002 | 8.24E-48 | 1.47E-72 | 2.10E-14 |
| rs10790164 | G | A | APOA4_APOC3 | 9 | 0.022 | 0.052 | 0.042 | 0.001 | 0.002 | 0.005 | 5.27E-70 | 1.92E-190 | 9.49E-19 |
| rs10802508 | G | A | OR2B11_OR2W5 | 9 | 0.001 | 0.002 | 0.001 | 0.001 | 0.001 | 0.002 | 8.61E-02 | 1.89E-02 | 5.85E-01 |
| rs10991417 | C | A | ABCA1 | 9 | -0.002 | -0.003 | -0.003 | 0.000 | 0.001 | 0.002 | 8.99E-04 | 3.78E-05 | 6.89E-02 |
| rs11127131 | T | G | BRE | 9 | -0.006 | -0.011 | 0.007 | 0.003 | 0.004 | 0.012 | 7.14E-02 | 1.18E-02 | 5.52E-01 |
| rs111732554 | C | G | ZNF259_BUD13 | 9 | 0.006 | 0.011 | 0.010 | 0.002 | 0.002 | 0.006 | 3.83E-04 | 1.38E-06 | 9.56E-02 |
| rs11187065 | C | T | IDE | 9 | -0.002 | -0.003 | -0.002 | 0.001 | 0.001 | 0.002 | 6.24E-03 | 6.22E-05 | 3.13E-01 |
| rs11216060 | A | G | BUD13_LINC00900 | 9 | 0.003 | 0.008 | 0.003 | 0.001 | 0.001 | 0.002 | 2.68E-05 | 2.54E-16 | 1.83E-01 |
| rs11250031 | T | G | RP1L1_PRSS55 | 9 | 0.001 | 0.002 | 0.001 | 0.000 | 0.001 | 0.002 | 1.47E-01 | 3.74E-04 | 5.37E-01 |
| rs1126513 | T | G | HLA-DPB1_HLA-DPA1 | 9 | -0.002 | -0.005 | -0.004 | 0.001 | 0.001 | 0.002 | 1.39E-03 | 4.30E-12 | 3.78E-02 |
| rs1139653 | T | A | DNAJA3 | 9 | 0.002 | 0.004 | 0.004 | 0.001 | 0.001 | 0.002 | 6.91E-05 | 5.71E-08 | 6.14E-02 |
| rs114269061 | C | T | BRE | 9 | -0.009 | -0.017 | -0.025 | 0.003 | 0.004 | 0.011 | 3.57E-03 | 4.46E-05 | 2.75E-02 |
| rs114541589 | T | C | RBKS_MRPL33 | 9 | -0.006 | -0.010 | -0.008 | 0.002 | 0.003 | 0.007 | 1.07E-03 | 3.74E-05 | 2.45E-01 |
| rs1154410 | G | A | ADH5 | 9 | -0.001 | -0.003 | -0.001 | 0.001 | 0.001 | 0.002 | 4.78E-02 | 3.02E-04 | 5.68E-01 |
| rs116021689 | A | G | DPYSL5 | 9 | -0.008 | -0.016 | -0.025 | 0.002 | 0.003 | 0.008 | 7.80E-05 | 1.68E-08 | 1.23E-03 |
| rs11630209 | T | C | TYRO3_MGA | 9 | 0.002 | 0.003 | 0.002 | 0.001 | 0.001 | 0.002 | 2.70E-05 | 2.48E-05 | 2.80E-01 |
| rs11681399 | G | A | CASP8 | 9 | -0.002 | -0.004 | -0.004 | 0.000 | 0.001 | 0.002 | 4.35E-04 | 9.34E-08 | 3.76E-02 |
| rs116998829 | G | A | CSGALNACT1 | 9 | 0.003 | 0.007 | 0.005 | 0.001 | 0.002 | 0.004 | 1.11E-02 | 1.72E-05 | 2.28E-01 |
| rs117794084 | T | G | BUD13_LINC00900 | 9 | -0.011 | -0.030 | -0.017 | 0.002 | 0.003 | 0.007 | 1.09E-08 | 1.76E-30 | 1.37E-02 |
| rs11782626 | G | T | LPL_SLC18A1 | 9 | -0.009 | -0.018 | -0.020 | 0.002 | 0.003 | 0.007 | 1.21E-06 | 1.05E-11 | 5.28E-03 |
| rs118045108 | T | C | LPL_SLC18A1 | 9 | -0.006 | -0.010 | -0.006 | 0.002 | 0.002 | 0.006 | 7.76E-05 | 6.94E-06 | 3.06E-01 |
| rs12139515 | G | C | ELAVL4_DMRTA2 | 9 | 0.001 | 0.003 | 0.001 | 0.001 | 0.001 | 0.002 | 1.49E-02 | 1.61E-04 | 6.74E-01 |
| rs12280753 | T | C | BUD13_LINC00900 | 9 | -0.022 | -0.049 | -0.041 | 0.001 | 0.001 | 0.003 | 2.12E-142 | 0.00E+00 | 3.12E-37 |
| rs12298717 | G | A | RFX4_LOC100287944 | 9 | -0.002 | -0.004 | -0.004 | 0.000 | 0.001 | 0.002 | 2.14E-07 | 2.09E-11 | 2.09E-02 |
| rs12362930 | C | T | BUD13_LINC00900 | 9 | 0.003 | 0.007 | 0.006 | 0.001 | 0.001 | 0.004 | 7.48E-03 | 9.60E-08 | 1.33E-01 |
| rs1240660 | A | G | BUD13_LINC00900 | 9 | -0.002 | -0.006 | -0.003 | 0.000 | 0.001 | 0.002 | 2.66E-06 | 7.48E-18 | 8.03E-02 |
| rs1242498 | T | G | MED9_SMCR9 | 9 | 0.002 | 0.004 | 0.002 | 0.001 | 0.001 | 0.002 | 5.31E-06 | 4.43E-07 | 3.77E-01 |
| rs12454712 | C | T | BCL2 | 9 | 0.002 | 0.003 | -0.001 | 0.000 | 0.001 | 0.002 | 7.72E-06 | 1.29E-05 | 7.24E-01 |
| rs1263173 | A | G | APOA5_APOA4 | 9 | 0.002 | 0.007 | 0.005 | 0.001 | 0.001 | 0.002 | 1.67E-06 | 1.08E-22 | 8.11E-03 |
| rs12805779 | T | G | BUD13_LINC00900 | 9 | 0.004 | 0.008 | 0.010 | 0.001 | 0.001 | 0.002 | 1.09E-09 | 2.70E-20 | 1.70E-05 |
| rs13009143 | A | G | MIR5702_LOC646736 | 9 | -0.002 | -0.005 | -0.002 | 0.001 | 0.001 | 0.002 | 2.19E-04 | 1.10E-07 | 4.18E-01 |
| rs13027424 | A | G | MRPL33_LOC100129995 | 9 | -0.008 | -0.013 | -0.017 | 0.002 | 0.003 | 0.007 | 1.60E-05 | 6.26E-07 | 1.43E-02 |
| rs13070993 | T | C | SYN2 | 9 | 0.003 | 0.006 | 0.003 | 0.001 | 0.001 | 0.004 | 3.85E-04 | 2.84E-05 | 4.45E-01 |
| rs13133548 | A | G | FAM13A | 9 | -0.003 | -0.004 | -0.003 | 0.000 | 0.001 | 0.002 | 6.42E-10 | 1.88E-10 | 4.76E-02 |
| rs13147739 | A | G | SLC10A6 | 9 | 0.004 | 0.005 | 0.005 | 0.001 | 0.001 | 0.003 | 5.75E-06 | 8.57E-06 | 1.10E-01 |
| rs13274062 | C | G | XKR9_EYA1 | 9 | 0.002 | 0.003 | 0.001 | 0.000 | 0.001 | 0.002 | 1.74E-04 | 1.77E-06 | 5.18E-01 |
| rs149117895 | A | C | FNDC4_IFT172 | 9 | -0.013 | -0.024 | -0.020 | 0.002 | 0.003 | 0.009 | 4.37E-07 | 5.81E-12 | 3.11E-02 |
| rs1492641 | G | A | INTS10_CSGALNACT1 | 9 | 0.001 | 0.005 | 0.003 | 0.001 | 0.001 | 0.002 | 2.64E-02 | 7.97E-10 | 1.83E-01 |
| rs1508099 | C | A | BUD13_LINC00900 | 9 | -0.001 | -0.005 | -0.006 | 0.000 | 0.001 | 0.002 | 2.70E-03 | 5.55E-11 | 6.39E-04 |
| rs1518303 | A | G | LOC646736_MIR548AR | 9 | -0.002 | -0.005 | -0.004 | 0.001 | 0.001 | 0.002 | 3.18E-03 | 6.64E-08 | 1.26E-01 |
| rs1522837 | G | T | SGK223_CLDN23 | 9 | -0.002 | -0.007 | -0.005 | 0.001 | 0.001 | 0.004 | 9.46E-02 | 2.78E-06 | 1.92E-01 |
| rs1597944 | T | C | USP40_UGT1A8 | 9 | 0.003 | 0.006 | 0.006 | 0.000 | 0.001 | 0.002 | 2.14E-08 | 2.43E-21 | 2.62E-04 |
| rs16895971 | C | T | LCORL | 9 | -0.003 | -0.004 | -0.004 | 0.001 | 0.001 | 0.002 | 9.18E-05 | 1.04E-05 | 1.35E-01 |
| rs17091325 | A | G | INTS10_CSGALNACT1 | 9 | 0.003 | 0.006 | 0.008 | 0.001 | 0.001 | 0.003 | 4.05E-04 | 3.16E-08 | 9.10E-03 |
| rs17419675 | A | G | PTPN13 | 9 | 0.002 | 0.005 | 0.005 | 0.001 | 0.001 | 0.003 | 4.81E-03 | 9.97E-08 | 6.08E-02 |
| rs17489871 | C | T | SLC18A1_LPL | 9 | 0.002 | 0.003 | 0.001 | 0.001 | 0.001 | 0.002 | 1.31E-03 | 1.81E-05 | 6.68E-01 |
| rs17630640 | G | A | C6orf118_QKI | 9 | 0.002 | 0.005 | 0.003 | 0.001 | 0.001 | 0.003 | 4.20E-04 | 1.07E-07 | 2.38E-01 |
| rs1799816 | T | C | INSR | 9 | 0.009 | 0.023 | 0.030 | 0.003 | 0.004 | 0.011 | 3.13E-03 | 4.14E-09 | 4.18E-03 |
| rs1804080 | C | G | HERC3 | 9 | 0.002 | 0.003 | 0.000 | 0.001 | 0.001 | 0.002 | 2.55E-03 | 2.68E-04 | 9.56E-01 |
| rs1982784 | G | A | MGRN1_UBALD1 | 9 | 0.003 | 0.004 | 0.005 | 0.001 | 0.001 | 0.002 | 1.32E-05 | 8.81E-07 | 2.85E-02 |
| rs1998064 | G | T | GALNT2 | 9 | 0.004 | 0.007 | 0.007 | 0.001 | 0.001 | 0.002 | 6.56E-16 | 6.08E-21 | 6.18E-04 |
| rs201862465 | T | C | KANK3 | 9 | 0.003 | 0.005 | 0.003 | 0.001 | 0.001 | 0.002 | 9.97E-07 | 1.00E-10 | 1.63E-01 |
| rs2156122 | A | C | BUD13_LINC00900 | 9 | -0.002 | -0.007 | -0.002 | 0.001 | 0.001 | 0.002 | 2.34E-05 | 3.21E-19 | 3.13E-01 |
| rs2168333 | G | T | LYPLAL1_RNU5F-1 | 9 | 0.001 | 0.003 | 0.001 | 0.000 | 0.001 | 0.002 | 3.48E-03 | 1.50E-06 | 4.02E-01 |
| rs2292137 | G | C | HIP1R | 9 | -0.002 | -0.003 | -0.003 | 0.000 | 0.001 | 0.002 | 2.92E-04 | 4.25E-06 | 5.29E-02 |
| rs2293277 | T | A | CEP55 | 9 | 0.002 | 0.003 | 0.002 | 0.001 | 0.001 | 0.002 | 3.01E-03 | 6.25E-06 | 4.18E-01 |
| rs2431032 | C | T | TCF12 | 9 | 0.001 | 0.004 | 0.004 | 0.001 | 0.001 | 0.002 | 5.78E-03 | 4.12E-07 | 7.37E-02 |
| rs2509209 | G | A | PCSK7 | 9 | -0.002 | -0.004 | -0.005 | 0.000 | 0.001 | 0.002 | 3.13E-05 | 2.12E-10 | 5.34E-03 |
| rs2518049 | G | A | AKR1C3 | 9 | -0.001 | -0.005 | -0.008 | 0.001 | 0.001 | 0.002 | 1.92E-02 | 3.45E-10 | 5.53E-04 |
| rs2782981 | C | T | ADRB1_NHLRC2 | 9 | 0.002 | 0.003 | 0.004 | 0.001 | 0.001 | 0.002 | 5.97E-04 | 2.01E-05 | 1.98E-02 |
| rs2966079 | T | C | CMIP | 9 | 0.003 | 0.003 | 0.002 | 0.000 | 0.001 | 0.002 | 1.50E-09 | 3.31E-06 | 3.75E-01 |
| rs34645420 | C | T | IGF1R_FAM169B | 9 | -0.001 | -0.002 | -0.001 | 0.000 | 0.001 | 0.002 | 2.27E-02 | 3.51E-04 | 4.15E-01 |
| rs34715654 | A | G | FIZ1 | 9 | -0.002 | -0.005 | -0.005 | 0.001 | 0.001 | 0.002 | 6.75E-04 | 8.17E-10 | 3.74E-02 |
| rs35266076 | A | G | BUD13_LINC00900 | 9 | -0.006 | -0.016 | -0.013 | 0.001 | 0.002 | 0.005 | 3.10E-05 | 2.09E-16 | 9.18E-03 |
| rs38173 | G | A | ISPD_MEOX2 | 9 | 0.001 | 0.002 | 0.000 | 0.001 | 0.001 | 0.002 | 1.79E-02 | 3.55E-03 | 9.58E-01 |
| rs3851294 | G | A | DSTYK | 9 | -0.003 | -0.004 | -0.005 | 0.001 | 0.001 | 0.003 | 8.55E-05 | 1.40E-04 | 1.03E-01 |
| rs3922 | G | A | CXCR5_BCL9L | 9 | -0.002 | -0.003 | -0.002 | 0.000 | 0.001 | 0.002 | 5.48E-04 | 2.24E-05 | 1.56E-01 |
| rs41288829 | T | C | EIF2B4 | 9 | -0.008 | -0.014 | -0.014 | 0.001 | 0.002 | 0.005 | 9.61E-08 | 4.09E-12 | 1.21E-02 |
| rs41292412 | T | C | MIR122_MIR3591 | 9 | -0.011 | -0.019 | -0.018 | 0.002 | 0.003 | 0.008 | 1.32E-06 | 1.22E-09 | 3.77E-02 |
| rs41355649 | A | G | SLC7A10_CEBPA | 9 | -0.003 | -0.007 | -0.005 | 0.001 | 0.001 | 0.004 | 1.56E-03 | 2.10E-08 | 1.62E-01 |
| rs4332747 | T | G | ABCA17P | 9 | 0.001 | 0.003 | 0.004 | 0.000 | 0.001 | 0.002 | 3.71E-03 | 7.47E-06 | 5.33E-02 |
| rs4610302 | G | A | SPARCL1 | 9 | -0.001 | -0.005 | -0.005 | 0.000 | 0.001 | 0.002 | 5.51E-03 | 3.61E-12 | 5.98E-03 |
| rs4711749 | G | A | VEGFA_MRPS18A | 9 | -0.002 | -0.003 | -0.001 | 0.001 | 0.001 | 0.002 | 3.35E-03 | 7.11E-05 | 6.33E-01 |
| rs4711750 | A | T | VEGFA_LOC100132354 | 9 | -0.007 | -0.010 | -0.008 | 0.000 | 0.001 | 0.002 | 5.22E-49 | 2.39E-55 | 2.60E-06 |
| rs4889326 | T | C | CMIP | 9 | 0.003 | 0.004 | 0.000 | 0.001 | 0.001 | 0.002 | 2.73E-06 | 1.54E-06 | 8.36E-01 |
| rs4915846 | T | C | DOCK7 | 9 | -0.002 | -0.007 | -0.010 | 0.001 | 0.001 | 0.004 | 1.35E-02 | 8.97E-08 | 5.86E-03 |
| rs506222 | C | A | BUD13_LINC00900 | 9 | -0.012 | -0.029 | -0.024 | 0.001 | 0.001 | 0.003 | 3.46E-55 | 4.12E-164 | 2.67E-17 |
| rs522645 | A | C | PCSK7 | 9 | -0.004 | -0.008 | -0.008 | 0.001 | 0.001 | 0.002 | 4.20E-13 | 4.35E-27 | 1.70E-04 |
| rs558056 | G | T | BUD13_LINC00900 | 9 | -0.004 | -0.008 | -0.006 | 0.000 | 0.001 | 0.002 | 1.45E-14 | 2.15E-31 | 2.72E-04 |
| rs56005336 | G | C | GRM4_HMGA1 | 9 | -0.006 | -0.009 | -0.010 | 0.001 | 0.002 | 0.004 | 9.02E-08 | 9.64E-09 | 2.65E-02 |
| rs56301507 | A | G | FKBP6 | 9 | 0.004 | 0.008 | 0.003 | 0.001 | 0.001 | 0.004 | 1.33E-05 | 2.05E-08 | 4.78E-01 |
| rs57400569 | A | G | FAM13A | 9 | 0.003 | 0.003 | 0.002 | 0.001 | 0.001 | 0.002 | 2.51E-08 | 1.39E-05 | 4.30E-01 |
| rs59496970 | G | A | BRE | 9 | 0.002 | 0.004 | 0.006 | 0.001 | 0.001 | 0.002 | 9.01E-04 | 4.45E-06 | 9.97E-03 |
| rs6090566 | C | T | SLC2A10_EYA2 | 9 | -0.002 | -0.004 | -0.004 | 0.000 | 0.001 | 0.002 | 3.64E-05 | 7.04E-09 | 1.95E-02 |
| rs61747073 | T | C | IFT172 | 9 | 0.007 | 0.013 | 0.017 | 0.002 | 0.002 | 0.006 | 3.20E-05 | 2.59E-08 | 4.72E-03 |
| rs61903415 | G | A | SIK3 | 9 | 0.004 | 0.009 | 0.007 | 0.001 | 0.001 | 0.003 | 1.30E-07 | 4.54E-17 | 7.43E-03 |
| rs62070804 | T | C | ABHD15 | 9 | -0.007 | -0.013 | -0.013 | 0.002 | 0.002 | 0.007 | 4.56E-05 | 1.99E-07 | 5.19E-02 |
| rs62375113 | T | C | TNFAIP8 | 9 | 0.002 | 0.003 | 0.003 | 0.001 | 0.001 | 0.002 | 4.05E-05 | 3.12E-05 | 1.70E-01 |
| rs62386480 | T | G | CLINT1_EBF1 | 9 | 0.001 | 0.003 | 0.004 | 0.000 | 0.001 | 0.002 | 2.81E-03 | 1.70E-05 | 2.41E-02 |
| rs645411 | C | T | ALG9 | 9 | 0.002 | 0.003 | 0.005 | 0.000 | 0.001 | 0.002 | 7.97E-07 | 2.91E-07 | 2.70E-03 |
| rs6586879 | T | C | LPL_INTS10 | 9 | -0.014 | -0.027 | -0.012 | 0.002 | 0.002 | 0.006 | 1.30E-20 | 2.57E-36 | 4.33E-02 |
| rs66489250 | C | T | BRE | 9 | -0.005 | -0.008 | -0.011 | 0.001 | 0.001 | 0.003 | 8.99E-11 | 1.51E-13 | 2.28E-04 |
| rs6670040 | T | C | ATP2B4_OPTC | 9 | 0.001 | 0.003 | 0.001 | 0.000 | 0.001 | 0.002 | 2.54E-03 | 6.67E-05 | 5.55E-01 |
| rs6710941 | T | C | CENPA | 9 | -0.003 | -0.007 | -0.008 | 0.001 | 0.001 | 0.003 | 1.68E-03 | 1.46E-07 | 1.78E-02 |
| rs71539599 | G | A | CHN2 | 9 | -0.002 | -0.005 | -0.007 | 0.001 | 0.001 | 0.002 | 5.62E-05 | 6.64E-10 | 1.48E-03 |
| rs729005 | C | T | TRIB1_LINC00861 | 9 | -0.003 | -0.005 | -0.004 | 0.001 | 0.001 | 0.003 | 1.08E-04 | 2.90E-05 | 1.90E-01 |
| rs72927213 | C | G | TUT1_MIR3654 | 9 | -0.002 | -0.004 | -0.002 | 0.000 | 0.001 | 0.002 | 5.01E-05 | 1.25E-08 | 3.12E-01 |
| rs73010409 | C | T | RNF214 | 9 | -0.005 | -0.010 | -0.009 | 0.001 | 0.001 | 0.003 | 2.01E-10 | 1.59E-22 | 6.20E-04 |
| rs73011197 | T | C | BUD13_LINC00900 | 9 | -0.004 | -0.010 | -0.006 | 0.001 | 0.002 | 0.005 | 1.89E-03 | 1.61E-08 | 1.77E-01 |
| rs73207880 | A | G | NAT2_PSD3 | 9 | 0.002 | 0.005 | 0.005 | 0.001 | 0.001 | 0.002 | 3.46E-03 | 3.47E-08 | 1.58E-02 |
| rs74332127 | A | G | BUD13_LINC00900 | 9 | -0.005 | -0.016 | -0.017 | 0.002 | 0.003 | 0.007 | 6.96E-03 | 1.10E-08 | 1.97E-02 |
| rs74709851 | G | A | LPL_SLC18A1 | 9 | -0.006 | -0.011 | -0.013 | 0.002 | 0.002 | 0.007 | 5.04E-04 | 4.23E-06 | 5.58E-02 |
| rs74773964 | C | T | ZNF259 | 9 | 0.002 | 0.009 | 0.005 | 0.002 | 0.002 | 0.006 | 1.70E-01 | 2.65E-04 | 4.03E-01 |
| rs74949497 | C | A | CENPW_RSPO3 | 9 | -0.006 | -0.011 | -0.008 | 0.002 | 0.002 | 0.006 | 2.86E-04 | 1.47E-06 | 2.00E-01 |
| rs756145 | A | G | DMRT2_DMRT3 | 9 | -0.001 | -0.003 | -0.002 | 0.000 | 0.001 | 0.002 | 1.31E-02 | 2.46E-04 | 2.43E-01 |
| rs7572857 | A | G | CEP68 | 9 | 0.002 | 0.003 | 0.002 | 0.001 | 0.001 | 0.002 | 8.67E-04 | 1.55E-04 | 4.87E-01 |
| rs7588910 | G | A | SLC5A6_TCF23 | 9 | -0.003 | -0.007 | -0.005 | 0.000 | 0.001 | 0.002 | 3.16E-13 | 1.77E-24 | 6.17E-03 |
| rs76471230 | C | T | DPYSL5_MAPRE3 | 9 | 0.005 | 0.009 | 0.009 | 0.001 | 0.002 | 0.004 | 1.83E-05 | 1.99E-09 | 3.44E-02 |
| rs76490719 | G | T | IRS1 | 9 | -0.003 | -0.006 | -0.006 | 0.001 | 0.002 | 0.004 | 7.43E-03 | 1.94E-04 | 1.35E-01 |
| rs768187 | G | C | INTS10_CSGALNACT1 | 9 | 0.003 | 0.004 | 0.004 | 0.001 | 0.001 | 0.002 | 1.75E-06 | 4.26E-07 | 1.13E-01 |
| rs77244720 | T | G | KCNK3_SLC35F6 | 9 | 0.006 | 0.011 | 0.011 | 0.002 | 0.002 | 0.006 | 5.32E-05 | 1.86E-07 | 6.02E-02 |
| rs77488547 | C | T | HSPA1A_HSPA1B | 9 | -0.002 | -0.009 | -0.003 | 0.002 | 0.002 | 0.006 | 1.68E-01 | 7.12E-05 | 5.79E-01 |
| rs77585907 | T | C | SLC5A6_TCF23 | 9 | -0.007 | -0.014 | -0.021 | 0.002 | 0.003 | 0.008 | 2.04E-03 | 2.95E-06 | 1.38E-02 |
| rs77741956 | T | G | MSRA | 9 | 0.002 | 0.005 | 0.005 | 0.001 | 0.001 | 0.003 | 2.09E-02 | 1.79E-07 | 6.44E-02 |
| rs77853072 | T | G | GALNT2_URB2 | 9 | -0.002 | -0.005 | -0.004 | 0.001 | 0.001 | 0.003 | 4.68E-03 | 1.33E-06 | 8.98E-02 |
| rs78035324 | A | G | DNAH10 | 9 | 0.004 | 0.005 | 0.003 | 0.001 | 0.001 | 0.003 | 2.45E-06 | 9.96E-06 | 2.81E-01 |
| rs7812740 | C | T | CSGALNACT1_INTS10 | 9 | 0.002 | 0.004 | 0.001 | 0.001 | 0.001 | 0.002 | 1.41E-03 | 5.76E-07 | 5.91E-01 |
| rs7818070 | A | G | INTS10_LPL | 9 | -0.002 | -0.005 | -0.003 | 0.000 | 0.001 | 0.002 | 6.01E-08 | 1.04E-12 | 7.48E-02 |
| rs78248243 | A | G | CSGALNACT1_INTS10 | 9 | 0.004 | 0.009 | 0.005 | 0.001 | 0.002 | 0.005 | 8.26E-03 | 3.96E-06 | 3.31E-01 |
| rs78476354 | A | G | GUCY2GP | 9 | 0.001 | 0.005 | 0.003 | 0.001 | 0.001 | 0.003 | 3.90E-01 | 4.30E-06 | 3.53E-01 |
| rs79293855 | A | G | ANGPTL4_KANK3 | 9 | 0.006 | 0.014 | 0.010 | 0.001 | 0.002 | 0.005 | 1.95E-05 | 3.65E-14 | 4.22E-02 |
| rs80312606 | C | T | CLIP2_RFC2 | 9 | 0.002 | 0.004 | 0.005 | 0.001 | 0.001 | 0.003 | 2.14E-02 | 1.80E-03 | 9.83E-02 |
| rs879858 | A | G | BUD13_LINC00900 | 9 | -0.018 | -0.042 | -0.031 | 0.002 | 0.003 | 0.007 | 5.67E-23 | 1.40E-60 | 5.97E-06 |
| rs897456 | G | A | PEMT | 9 | -0.003 | -0.008 | -0.008 | 0.001 | 0.001 | 0.004 | 1.34E-03 | 1.89E-09 | 3.23E-02 |
| rs921968 | T | G | VIL1_CTDSP1 | 9 | -0.002 | -0.005 | -0.005 | 0.000 | 0.001 | 0.002 | 5.49E-06 | 3.90E-12 | 9.24E-03 |
| rs9257793 | C | T | OR5V1_OR12D3 | 9 | 0.002 | 0.007 | 0.006 | 0.001 | 0.001 | 0.003 | 9.18E-04 | 6.36E-13 | 1.07E-02 |
| rs9267820 | A | G | NOTCH4 | 9 | -0.001 | -0.003 | -0.001 | 0.001 | 0.001 | 0.002 | 3.34E-02 | 9.97E-06 | 6.51E-01 |
| rs9376420 | C | A | LOC645434_LOC100132735 | 9 | -0.001 | -0.003 | -0.004 | 0.000 | 0.001 | 0.002 | 2.25E-03 | 6.11E-06 | 4.17E-02 |
| rs9461755 | A | G | HLA-DRB5_HLA-DRA | 9 | -0.006 | -0.016 | -0.016 | 0.001 | 0.002 | 0.004 | 1.66E-07 | 1.77E-23 | 1.53E-04 |
| rs9619724 | G | T | PLA2G6 | 9 | 0.002 | 0.004 | 0.003 | 0.000 | 0.001 | 0.002 | 1.93E-06 | 5.14E-11 | 9.59E-02 |
| rs9926861 | G | T | FAM92B_LOC400548 | 9 | -0.001 | -0.004 | 0.001 | 0.001 | 0.001 | 0.003 | 7.38E-02 | 1.56E-04 | 6.26E-01 |
| rs10103634 | A | G | LPL_SLC18A1 | 10 | 0.003 | 0.012 | 0.002 | 0.000 | 0.001 | 0.002 | 3.55E-12 | 1.25E-69 | 3.39E-01 |
| rs1018079 | C | T | SLC18A1 | 10 | 0.002 | 0.005 | 0.002 | 0.000 | 0.001 | 0.002 | 5.98E-04 | 2.19E-12 | 2.25E-01 |
| rs10431079 | G | A | BUD13_LINC00900 | 10 | 0.000 | -0.003 | 0.001 | 0.001 | 0.001 | 0.002 | 9.00E-01 | 7.18E-04 | 5.57E-01 |
| rs1047964 | C | G | BACE1_RNF214 | 10 | 0.001 | 0.010 | 0.003 | 0.001 | 0.001 | 0.004 | 2.34E-01 | 3.28E-11 | 4.56E-01 |
| rs10484439 | A | G | HIST1H4I_BTN3A2 | 10 | 0.001 | 0.006 | 0.003 | 0.001 | 0.001 | 0.003 | 6.26E-02 | 1.82E-09 | 2.77E-01 |
| rs10503407 | G | C | MSRA | 10 | 0.001 | 0.004 | 0.001 | 0.001 | 0.001 | 0.003 | 2.94E-01 | 2.34E-05 | 8.45E-01 |
| rs1057373 | A | C | TAP1_PSMB9 | 10 | -0.002 | -0.006 | -0.005 | 0.001 | 0.001 | 0.003 | 1.53E-02 | 4.92E-08 | 1.36E-01 |
| rs10838612 | T | G | ARHGAP1 | 10 | 0.002 | 0.006 | 0.001 | 0.001 | 0.001 | 0.003 | 6.07E-03 | 1.50E-07 | 8.06E-01 |
| rs11043307 | T | C | BCL7A | 10 | -0.002 | -0.004 | -0.001 | 0.001 | 0.001 | 0.002 | 3.42E-03 | 1.63E-05 | 6.93E-01 |
| rs11071896 | G | A | ZWILCH | 10 | -0.001 | -0.003 | 0.001 | 0.001 | 0.001 | 0.002 | 3.01E-02 | 1.20E-04 | 5.58E-01 |
| rs111796894 | G | A | VPS37D_MLXIPL | 10 | -0.002 | -0.005 | -0.002 | 0.001 | 0.001 | 0.003 | 4.98E-02 | 1.61E-05 | 4.46E-01 |
| rs11186669 | C | T | PPP1R3C_TNKS2 | 10 | -0.002 | -0.005 | -0.002 | 0.001 | 0.001 | 0.004 | 2.90E-02 | 4.95E-05 | 5.75E-01 |
| rs11204087 | T | C | LPL_SLC18A1 | 10 | 0.005 | 0.013 | 0.005 | 0.000 | 0.001 | 0.002 | 3.64E-27 | 9.64E-83 | 8.39E-03 |
| rs11216028 | T | G | BUD13_LINC00900 | 10 | 0.001 | 0.005 | 0.002 | 0.001 | 0.001 | 0.003 | 1.53E-01 | 2.63E-07 | 5.31E-01 |
| rs112357006 | A | G | RNF214 | 10 | -0.001 | -0.009 | 0.000 | 0.001 | 0.002 | 0.004 | 2.46E-01 | 3.98E-09 | 9.57E-01 |
| rs1128349 | T | C | DNAJC30_WBSCR22 | 10 | 0.003 | 0.006 | 0.001 | 0.000 | 0.001 | 0.002 | 1.68E-08 | 1.74E-18 | 3.93E-01 |
| rs113067173 | A | G | CSGALNACT1_INTS10 | 10 | 0.003 | 0.007 | 0.000 | 0.001 | 0.002 | 0.004 | 9.87E-03 | 1.62E-05 | 9.10E-01 |
| rs113296769 | G | A | BAZ1B | 10 | 0.004 | 0.019 | -0.008 | 0.002 | 0.002 | 0.006 | 1.10E-02 | 3.48E-16 | 1.92E-01 |
| rs114594921 | C | T | BUD13_LINC00900 | 10 | -0.007 | -0.020 | -0.005 | 0.002 | 0.002 | 0.006 | 1.64E-05 | 5.78E-19 | 4.33E-01 |
| rs11600156 | G | A | BUD13_LINC00900 | 10 | 0.001 | 0.005 | 0.002 | 0.001 | 0.001 | 0.002 | 5.09E-02 | 1.68E-09 | 2.86E-01 |
| rs116843064 | A | G | ANGPTL4 | 10 | 0.012 | 0.045 | 0.001 | 0.002 | 0.002 | 0.007 | 1.67E-11 | 1.37E-79 | 8.28E-01 |
| rs116963079 | G | T | TPM1_LACTB | 10 | -0.001 | -0.005 | 0.001 | 0.001 | 0.001 | 0.004 | 1.51E-01 | 6.02E-05 | 8.81E-01 |
| rs11708337 | G | A | SFMBT1 | 10 | 0.002 | 0.006 | 0.005 | 0.001 | 0.001 | 0.003 | 3.09E-04 | 1.11E-10 | 7.68E-02 |
| rs117191103 | T | C | SNX21 | 10 | -0.007 | -0.012 | -0.008 | 0.002 | 0.002 | 0.006 | 2.06E-05 | 4.98E-07 | 1.93E-01 |
| rs117604010 | A | G | LPL_SLC18A1 | 10 | 0.009 | 0.027 | 0.010 | 0.002 | 0.002 | 0.007 | 5.84E-08 | 3.60E-29 | 1.30E-01 |
| rs11760752 | A | C | MLXIPL | 10 | -0.001 | -0.004 | 0.001 | 0.001 | 0.001 | 0.002 | 3.68E-02 | 1.42E-07 | 5.31E-01 |
| rs117742492 | A | G | BUD13_LINC00900 | 10 | 0.002 | 0.009 | 0.000 | 0.001 | 0.001 | 0.004 | 1.03E-01 | 2.35E-12 | 9.41E-01 |
| rs11777330 | G | A | LINC00208_GATA4 | 10 | 0.000 | 0.004 | -0.003 | 0.001 | 0.001 | 0.003 | 6.78E-01 | 1.68E-04 | 3.93E-01 |
| rs118091818 | C | A | EIF3J-AS1_CTDSPL2 | 10 | -0.003 | -0.007 | -0.003 | 0.001 | 0.001 | 0.003 | 2.60E-03 | 5.62E-09 | 3.35E-01 |
| rs11882393 | T | G | MYO1F | 10 | 0.001 | 0.004 | -0.003 | 0.001 | 0.001 | 0.003 | 3.36E-01 | 1.22E-04 | 2.08E-01 |
| rs12174151 | T | C | PPP1R10 | 10 | -0.001 | -0.005 | 0.002 | 0.001 | 0.001 | 0.002 | 3.90E-01 | 2.39E-07 | 3.50E-01 |
| rs1233492 | G | A | MAS1L_LOC100507362 | 10 | 0.000 | -0.003 | -0.002 | 0.001 | 0.001 | 0.002 | 4.20E-01 | 5.91E-06 | 3.74E-01 |
| rs1239947 | T | C | DLEU1_DLEU7 | 10 | 0.000 | -0.002 | 0.001 | 0.000 | 0.001 | 0.002 | 4.65E-01 | 1.99E-03 | 4.56E-01 |
| rs12679591 | C | T | PRSS55 | 10 | 0.000 | 0.003 | -0.003 | 0.001 | 0.001 | 0.003 | 5.97E-01 | 9.05E-04 | 2.12E-01 |
| rs12721043 | A | C | APOA4 | 10 | 0.005 | 0.032 | 0.009 | 0.002 | 0.003 | 0.008 | 1.82E-02 | 5.31E-25 | 2.58E-01 |
| rs12910886 | A | C | FRMD5 | 10 | 0.003 | 0.007 | -0.002 | 0.001 | 0.001 | 0.003 | 4.92E-04 | 2.31E-08 | 5.85E-01 |
| rs12967290 | T | C | SETBP1 | 10 | -0.001 | -0.003 | -0.002 | 0.000 | 0.001 | 0.002 | 2.98E-03 | 5.02E-07 | 4.06E-01 |
| rs13066793 | G | A | VGLL3 | 10 | 0.001 | 0.004 | -0.003 | 0.001 | 0.001 | 0.003 | 2.65E-01 | 1.33E-04 | 2.91E-01 |
| rs13236513 | A | G | TYW1B | 10 | -0.002 | -0.005 | 0.005 | 0.001 | 0.002 | 0.005 | 2.26E-01 | 1.19E-02 | 3.57E-01 |
| rs13242693 | T | C | DNAJC30_VPS37D | 10 | 0.006 | 0.019 | 0.007 | 0.001 | 0.002 | 0.005 | 1.98E-06 | 4.52E-26 | 1.60E-01 |
| rs13267032 | A | G | INTS10_LPL | 10 | -0.001 | -0.005 | -0.002 | 0.001 | 0.001 | 0.002 | 3.33E-03 | 1.22E-10 | 4.32E-01 |
| rs1326775 | T | C | CENPP | 10 | -0.001 | -0.003 | 0.001 | 0.001 | 0.001 | 0.002 | 2.88E-02 | 1.67E-04 | 6.81E-01 |
| rs13434675 | A | G | CLCN3 | 10 | -0.002 | -0.005 | -0.004 | 0.001 | 0.001 | 0.003 | 6.56E-03 | 5.80E-05 | 2.32E-01 |
| rs143192984 | A | G | FCGR1A_HIST2H2BF | 10 | 0.002 | 0.004 | 0.001 | 0.001 | 0.001 | 0.003 | 2.00E-02 | 4.17E-04 | 7.96E-01 |
| rs1441778 | T | C | INTS10_LPL | 10 | -0.004 | -0.012 | -0.002 | 0.001 | 0.001 | 0.002 | 4.68E-10 | 6.93E-40 | 3.56E-01 |
| rs1508102 | A | G | BUD13_LINC00900 | 10 | -0.003 | -0.010 | -0.008 | 0.001 | 0.001 | 0.003 | 2.61E-03 | 2.61E-13 | 2.21E-02 |
| rs1605750 | A | G | KCNJ2 | 10 | 0.001 | 0.003 | 0.000 | 0.000 | 0.001 | 0.002 | 1.54E-03 | 8.04E-05 | 9.58E-01 |
| rs16845803 | G | A | TNFSF10_NCEH1 | 10 | -0.003 | -0.005 | -0.002 | 0.001 | 0.001 | 0.003 | 1.01E-04 | 5.03E-08 | 3.41E-01 |
| rs16851199 | G | A | GALNT2 | 10 | -0.007 | -0.012 | -0.013 | 0.001 | 0.002 | 0.006 | 1.37E-06 | 2.61E-09 | 2.02E-02 |
| rs17005322 | C | T | LYPLAL1_RNU5F-1 | 10 | -0.001 | -0.003 | -0.001 | 0.001 | 0.001 | 0.002 | 4.73E-02 | 2.45E-04 | 6.91E-01 |
| rs17091113 | T | C | CSGALNACT1_INTS10 | 10 | 0.001 | 0.004 | -0.001 | 0.000 | 0.001 | 0.002 | 5.84E-02 | 1.85E-07 | 6.37E-01 |
| rs17569676 | C | T | BUD13_LINC00900 | 10 | 0.001 | 0.004 | 0.002 | 0.001 | 0.001 | 0.002 | 7.85E-02 | 1.26E-06 | 4.29E-01 |
| rs1801700 | A | G | APOB | 10 | -0.004 | -0.005 | 0.000 | 0.001 | 0.001 | 0.004 | 2.96E-05 | 1.24E-03 | 9.39E-01 |
| rs180358 | T | C | BUD13_LINC00900 | 10 | 0.005 | 0.012 | 0.006 | 0.001 | 0.001 | 0.002 | 3.57E-17 | 2.31E-56 | 6.45E-03 |
| rs186868868 | A | C | LPL_SLC18A1 | 10 | -0.011 | -0.030 | -0.012 | 0.002 | 0.003 | 0.007 | 2.87E-09 | 4.66E-32 | 7.87E-02 |
| rs1984142 | C | G | MACF1 | 10 | 0.001 | 0.003 | -0.002 | 0.000 | 0.001 | 0.002 | 1.06E-01 | 6.52E-06 | 3.68E-01 |
| rs2154593 | G | A | LOC284889_GSTT2B | 10 | 0.001 | 0.003 | 0.001 | 0.000 | 0.001 | 0.002 | 1.51E-02 | 3.86E-06 | 5.95E-01 |
| rs2224198 | G | A | GSTA7P | 10 | 0.001 | 0.004 | 0.004 | 0.000 | 0.001 | 0.002 | 7.07E-03 | 4.18E-08 | 2.44E-02 |
| rs2412710 | A | G | CAPN3 | 10 | -0.007 | -0.013 | 0.003 | 0.002 | 0.002 | 0.006 | 1.14E-05 | 2.54E-08 | 5.96E-01 |
| rs2540953 | A | G | SLC1A4_CEP68 | 10 | 0.001 | 0.002 | -0.001 | 0.000 | 0.001 | 0.002 | 4.81E-02 | 2.60E-03 | 6.69E-01 |
| rs2602381 | C | T | UGT1A10_UGT1A9 | 10 | 0.001 | 0.007 | 0.002 | 0.000 | 0.001 | 0.002 | 2.37E-01 | 2.19E-30 | 1.53E-01 |
| rs2622525 | A | G | SLFN5_SLFN11 | 10 | 0.001 | 0.003 | 0.001 | 0.001 | 0.001 | 0.003 | 6.70E-02 | 1.32E-03 | 6.59E-01 |
| rs2645429 | G | A | FDFT1_NEIL2 | 10 | 0.001 | 0.006 | 0.002 | 0.001 | 0.001 | 0.002 | 7.33E-02 | 2.97E-15 | 4.32E-01 |
| rs2675638 | A | G | ARID5B_C10orf107 | 10 | 0.000 | -0.002 | 0.000 | 0.000 | 0.001 | 0.002 | 5.91E-01 | 5.28E-04 | 9.15E-01 |
| rs268 | G | A | LPL | 10 | -0.025 | -0.050 | -0.015 | 0.002 | 0.002 | 0.007 | 4.17E-45 | 5.44E-92 | 1.92E-02 |
| rs283814 | G | A | PVRL2 | 10 | -0.001 | -0.013 | -0.006 | 0.001 | 0.001 | 0.003 | 1.19E-01 | 4.66E-26 | 5.38E-02 |
| rs284032 | A | G | MAP3K1_ANKRD55 | 10 | -0.001 | -0.003 | 0.000 | 0.000 | 0.001 | 0.002 | 1.75E-02 | 1.49E-06 | 8.03E-01 |
| rs28594657 | A | C | TGM7 | 10 | -0.002 | -0.004 | -0.002 | 0.001 | 0.001 | 0.003 | 1.62E-03 | 1.90E-05 | 5.15E-01 |
| rs28989504 | C | T | BACE1_BACE1-AS | 10 | -0.001 | -0.005 | -0.003 | 0.001 | 0.001 | 0.004 | 2.02E-01 | 4.20E-04 | 4.12E-01 |
| rs295 | C | A | LPL | 10 | 0.010 | 0.025 | 0.009 | 0.001 | 0.001 | 0.002 | 8.80E-74 | 3.66E-238 | 4.66E-06 |
| rs35341095 | C | T | CALN1 | 10 | 0.001 | 0.003 | -0.002 | 0.001 | 0.001 | 0.003 | 2.37E-01 | 6.07E-03 | 5.82E-01 |
| rs35946942 | C | T | LPL_SLC18A1 | 10 | 0.003 | 0.006 | 0.002 | 0.001 | 0.001 | 0.003 | 6.96E-04 | 1.79E-08 | 4.25E-01 |
| rs3729848 | T | C | GATA4 | 10 | 0.001 | 0.005 | 0.001 | 0.001 | 0.001 | 0.002 | 5.39E-02 | 4.42E-07 | 8.20E-01 |
| rs3740690 | C | T | ARFGAP2 | 10 | -0.002 | -0.003 | -0.002 | 0.000 | 0.001 | 0.002 | 2.99E-04 | 1.61E-06 | 2.96E-01 |
| rs3741390 | T | C | SAC3D1 | 10 | 0.000 | -0.002 | 0.003 | 0.001 | 0.001 | 0.002 | 6.69E-01 | 2.06E-03 | 8.92E-02 |
| rs3892710 | T | C | HLA-DQB1_HLA-DQA2 | 10 | -0.001 | -0.003 | 0.005 | 0.001 | 0.001 | 0.002 | 2.76E-01 | 3.28E-03 | 3.27E-02 |
| rs41278045 | G | A | PLA2G12A | 10 | -0.011 | -0.059 | -0.049 | 0.006 | 0.008 | 0.022 | 5.27E-02 | 1.23E-13 | 2.21E-02 |
| rs41290102 | T | C | PVRL2 | 10 | 0.005 | 0.018 | 0.010 | 0.002 | 0.003 | 0.008 | 4.72E-02 | 8.13E-09 | 2.48E-01 |
| rs4342497 | A | G | GIMAP8_GIMAP7 | 10 | 0.001 | 0.002 | 0.000 | 0.000 | 0.001 | 0.002 | 2.37E-02 | 1.38E-03 | 8.82E-01 |
| rs4356265 | T | C | DSCAML1 | 10 | -0.001 | -0.005 | -0.003 | 0.001 | 0.001 | 0.003 | 2.12E-01 | 2.24E-08 | 2.22E-01 |
| rs4434872 | C | T | GATAD2B_SLC27A3 | 10 | -0.002 | -0.004 | -0.004 | 0.001 | 0.001 | 0.002 | 1.09E-03 | 6.20E-06 | 1.10E-01 |
| rs4759364 | T | C | HCAR1_KNTC1 | 10 | 0.001 | 0.005 | 0.002 | 0.001 | 0.001 | 0.002 | 2.71E-02 | 3.68E-08 | 2.87E-01 |
| rs4762753 | T | G | PDE3A | 10 | 0.001 | 0.003 | -0.001 | 0.001 | 0.001 | 0.002 | 2.99E-02 | 1.12E-03 | 6.72E-01 |
| rs4788887 | G | C | GRB2 | 10 | 0.000 | -0.004 | 0.000 | 0.001 | 0.001 | 0.002 | 5.76E-01 | 9.30E-07 | 8.85E-01 |
| rs4865796 | A | G | ARL15 | 10 | -0.001 | -0.003 | 0.004 | 0.000 | 0.001 | 0.002 | 5.88E-02 | 1.95E-05 | 2.80E-02 |
| rs4938399 | T | C | DSCAML1 | 10 | -0.001 | -0.003 | -0.001 | 0.001 | 0.001 | 0.002 | 3.98E-01 | 1.61E-04 | 6.33E-01 |
| rs4980661 | A | G | CCND1_MYEOV | 10 | -0.001 | -0.003 | -0.001 | 0.000 | 0.001 | 0.002 | 1.15E-02 | 1.50E-07 | 4.49E-01 |
| rs5093 | A | G | APOA4 | 10 | 0.003 | 0.011 | 0.000 | 0.002 | 0.002 | 0.006 | 5.62E-02 | 3.36E-06 | 9.63E-01 |
| rs519000 | T | C | BUD13_LINC00900 | 10 | -0.002 | -0.009 | -0.006 | 0.001 | 0.001 | 0.002 | 1.54E-04 | 7.36E-26 | 1.40E-02 |
| rs55682243 | G | C | SLC18A1_LPL | 10 | 0.009 | 0.018 | 0.008 | 0.001 | 0.002 | 0.005 | 2.04E-10 | 1.62E-20 | 1.17E-01 |
| rs593245 | T | C | BACE1 | 10 | 0.001 | 0.004 | 0.001 | 0.000 | 0.001 | 0.002 | 1.65E-02 | 3.42E-08 | 3.91E-01 |
| rs6099674 | T | C | PCK1_ZBP1 | 10 | -0.001 | -0.002 | -0.003 | 0.000 | 0.001 | 0.002 | 3.09E-02 | 2.26E-04 | 9.08E-02 |
| rs612822 | T | C | VPS37C | 10 | 0.000 | -0.002 | 0.000 | 0.000 | 0.001 | 0.002 | 6.23E-01 | 2.42E-03 | 8.68E-01 |
| rs6130848 | C | G | WFDC2_SPINT3 | 10 | 0.001 | 0.001 | -0.002 | 0.000 | 0.001 | 0.002 | 1.76E-03 | 1.57E-01 | 3.19E-01 |
| rs61819230 | T | C | HLX_C1orf140 | 10 | -0.002 | -0.005 | -0.003 | 0.001 | 0.001 | 0.003 | 1.26E-02 | 2.11E-06 | 2.74E-01 |
| rs62188980 | T | C | LOC646736_MIR548AR | 10 | 0.002 | 0.005 | 0.001 | 0.001 | 0.001 | 0.002 | 7.35E-04 | 3.61E-09 | 8.36E-01 |
| rs62498193 | A | C | LPL_SLC18A1 | 10 | -0.001 | -0.005 | 0.003 | 0.001 | 0.001 | 0.002 | 5.27E-02 | 8.04E-08 | 3.12E-01 |
| rs62498230 | C | T | SLC18A1 | 10 | -0.002 | -0.003 | -0.004 | 0.001 | 0.001 | 0.002 | 1.10E-02 | 2.30E-04 | 1.27E-01 |
| rs6510386 | G | A | PEPD | 10 | 0.002 | 0.003 | 0.001 | 0.001 | 0.001 | 0.002 | 2.65E-03 | 2.77E-07 | 6.03E-01 |
| rs6682741 | C | T | GALNT2 | 10 | 0.006 | 0.008 | 0.003 | 0.001 | 0.001 | 0.002 | 2.58E-24 | 5.68E-22 | 1.56E-01 |
| rs6816767 | G | A | PDGFC | 10 | 0.001 | 0.003 | -0.001 | 0.001 | 0.001 | 0.002 | 4.40E-02 | 3.87E-04 | 7.92E-01 |
| rs6861681 | A | G | CPEB4 | 10 | -0.001 | -0.003 | -0.004 | 0.000 | 0.001 | 0.002 | 1.05E-01 | 2.08E-06 | 6.18E-02 |
| rs6987457 | A | C | LPL_SLC18A1 | 10 | -0.003 | -0.005 | -0.003 | 0.000 | 0.001 | 0.002 | 3.35E-08 | 1.30E-13 | 6.18E-02 |
| rs7134375 | A | C | PDE3A_LOC100506393 | 10 | 0.002 | 0.003 | -0.001 | 0.000 | 0.001 | 0.002 | 1.74E-04 | 3.16E-07 | 5.71E-01 |
| rs718314 | G | A | ITPR2_SSPN | 10 | -0.001 | -0.003 | 0.003 | 0.001 | 0.001 | 0.002 | 6.59E-02 | 5.84E-04 | 8.64E-02 |
| rs7248104 | A | G | INSR | 10 | 0.001 | 0.003 | -0.001 | 0.000 | 0.001 | 0.002 | 1.21E-02 | 2.22E-06 | 6.04E-01 |
| rs72801433 | G | A | ZCCHC10 | 10 | 0.003 | 0.006 | 0.000 | 0.001 | 0.001 | 0.004 | 6.00E-04 | 1.48E-05 | 9.82E-01 |
| rs72836344 | T | G | ACOXL | 10 | 0.002 | 0.005 | 0.000 | 0.001 | 0.001 | 0.003 | 1.33E-03 | 4.95E-06 | 9.41E-01 |
| rs72836588 | T | C | MPP2_FAM215A | 10 | 0.001 | 0.004 | 0.002 | 0.001 | 0.001 | 0.002 | 4.45E-02 | 2.70E-06 | 4.25E-01 |
| rs73196880 | C | T | XKR6 | 10 | 0.000 | -0.004 | -0.002 | 0.001 | 0.001 | 0.002 | 9.03E-01 | 8.17E-06 | 3.41E-01 |
| rs73208819 | C | T | LPL_SLC18A1 | 10 | -0.004 | -0.009 | -0.003 | 0.001 | 0.001 | 0.003 | 9.16E-06 | 8.27E-14 | 2.92E-01 |
| rs74345411 | A | C | INTS10_LPL | 10 | 0.006 | 0.014 | 0.008 | 0.002 | 0.003 | 0.007 | 7.67E-04 | 4.91E-07 | 2.89E-01 |
| rs74737417 | G | C | LPL_SLC18A1 | 10 | 0.006 | 0.018 | 0.006 | 0.001 | 0.001 | 0.004 | 1.11E-10 | 1.25E-39 | 8.36E-02 |
| rs75466392 | T | C | CSGALNACT1_INTS10 | 10 | 0.005 | 0.014 | 0.005 | 0.002 | 0.002 | 0.006 | 5.76E-03 | 2.47E-09 | 4.16E-01 |
| rs75688370 | G | C | USP37 | 10 | -0.008 | -0.017 | -0.008 | 0.002 | 0.003 | 0.008 | 8.83E-05 | 4.68E-09 | 3.16E-01 |
| rs75813208 | T | C | DSCAML1 | 10 | -0.002 | -0.006 | -0.002 | 0.001 | 0.002 | 0.006 | 1.56E-01 | 6.53E-03 | 7.09E-01 |
| rs76026343 | A | G | UGT1A6_UGT1A10 | 10 | 0.002 | 0.013 | 0.007 | 0.001 | 0.002 | 0.005 | 2.70E-01 | 5.30E-11 | 1.92E-01 |
| rs76427249 | C | T | XKR6_PINX1 | 10 | 0.000 | -0.006 | 0.003 | 0.001 | 0.001 | 0.004 | 9.84E-01 | 3.32E-05 | 5.14E-01 |
| rs7694379 | A | G | KLHL8_MIR5705 | 10 | 0.000 | -0.006 | -0.003 | 0.000 | 0.001 | 0.002 | 8.29E-01 | 1.27E-18 | 8.53E-02 |
| rs769451 | G | T | APOE | 10 | 0.003 | 0.020 | 0.011 | 0.003 | 0.004 | 0.011 | 3.04E-01 | 4.58E-07 | 2.91E-01 |
| rs77393312 | A | G | CALN1_TYW1B | 10 | 0.004 | 0.010 | -0.001 | 0.001 | 0.002 | 0.005 | 5.53E-03 | 2.82E-07 | 8.82E-01 |
| rs78414336 | A | G | HSD17B4 | 10 | 0.002 | 0.005 | 0.001 | 0.001 | 0.001 | 0.002 | 2.41E-03 | 7.85E-07 | 6.33E-01 |
| rs79416478 | T | C | LPL_SLC18A1 | 10 | -0.003 | -0.009 | -0.002 | 0.001 | 0.001 | 0.004 | 6.67E-03 | 8.88E-13 | 5.86E-01 |
| rs7947951 | G | A | ARNTL | 10 | 0.000 | -0.002 | 0.000 | 0.000 | 0.001 | 0.002 | 6.95E-01 | 2.22E-03 | 9.94E-01 |
| rs79610135 | C | T | BUD13 | 10 | 0.003 | 0.009 | 0.004 | 0.001 | 0.002 | 0.005 | 5.33E-03 | 1.13E-07 | 4.28E-01 |
| rs79634051 | C | G | PSMA1 | 10 | 0.005 | 0.009 | 0.006 | 0.001 | 0.002 | 0.005 | 8.85E-04 | 6.11E-06 | 2.65E-01 |
| rs79670217 | G | T | ZNF652 | 10 | -0.002 | -0.005 | 0.000 | 0.001 | 0.001 | 0.004 | 2.15E-02 | 1.17E-04 | 9.47E-01 |
| rs79821925 | C | T | SLC18A1 | 10 | 0.008 | 0.018 | 0.008 | 0.002 | 0.003 | 0.009 | 9.42E-04 | 6.66E-08 | 3.89E-01 |
| rs799160 | T | C | VPS37D_MLXIPL | 10 | -0.003 | -0.007 | 0.001 | 0.000 | 0.001 | 0.002 | 1.73E-09 | 1.96E-31 | 6.48E-01 |
| rs8077889 | C | A | MPP3_C17orf105 | 10 | -0.002 | -0.003 | 0.000 | 0.001 | 0.001 | 0.002 | 4.13E-03 | 7.99E-06 | 8.93E-01 |
| rs8088721 | A | G | CTAGE1_GATA6 | 10 | -0.001 | -0.002 | 0.000 | 0.000 | 0.001 | 0.002 | 9.19E-03 | 1.11E-02 | 8.29E-01 |
| rs864745 | C | T | JAZF1 | 10 | 0.000 | 0.003 | -0.003 | 0.000 | 0.001 | 0.002 | 3.89E-01 | 7.43E-05 | 4.44E-02 |
| rs900400 | C | T | LEKR1_LINC00880 | 10 | 0.001 | 0.004 | -0.002 | 0.000 | 0.001 | 0.002 | 2.67E-01 | 7.06E-08 | 3.28E-01 |
| rs922485 | C | T | BLK | 10 | 0.000 | 0.005 | 0.000 | 0.001 | 0.001 | 0.003 | 7.78E-01 | 3.14E-07 | 9.29E-01 |
| rs9295474 | G | C | CDKAL1 | 10 | -0.001 | -0.002 | 0.002 | 0.000 | 0.001 | 0.002 | 1.07E-01 | 3.49E-03 | 2.33E-01 |

### **Table S2. Mendelian randomization sensitivity analyses of TRL-C-to-LDL-C ratio cluster in relation to AAA risk**

|  |  |  | **IVW-fixed effects** | | | **IVW-random effects** | | | **Weighted median** | | | **MR-Egger** | | |
| --- | --- | --- | --- | --- | --- | --- | --- | --- | --- | --- | --- | --- | --- | --- |
| **AAA source** | **Exposure** | **nsnp** | **Beta** | **SE** | **Pval** | **Beta** | **SE** | **Pval** | **Beta** | **SE** | **Pval** | **Beta** | **SE** | **Pval** |
| AAAgen | Set 1 | 136 | 0.924 | 0.112 | 1.96E-16 | 0.924 | 0.199 | 3.28E-06 | 0.810 | 0.196 | 3.59E-05 | 0.786 | 0.259 | 2.90E-03 |
| AAAgen | Set 2 | 134 | 1.700 | 0.126 | 9.27E-42 | 1.700 | 0.213 | 1.30E-15 | 1.320 | 0.261 | 4.03E-07 | 1.548 | 0.315 | 2.66E-06 |
| AAAgen | Set 3 | 134 | 2.055 | 0.112 | 1.16E-75 | 2.055 | 0.172 | 9.77E-33 | 1.188 | 0.237 | 5.15E-07 | 2.006 | 0.240 | 8.08E-14 |
| AAAgen | Set 4 | 136 | 1.831 | 0.171 | 8.04E-27 | 1.831 | 0.291 | 3.26E-10 | 2.371 | 0.315 | 5.08E-14 | 2.319 | 0.491 | 5.79E-06 |
| AAAgen | Set 5 | 136 | 1.524 | 0.189 | 7.78E-16 | 1.524 | 0.332 | 4.45E-06 | 1.747 | 0.415 | 2.60E-05 | 1.486 | 0.558 | 8.65E-03 |
| AAAgen | Set 6 | 135 | 1.821 | 0.192 | 2.34E-21 | 1.821 | 0.328 | 2.92E-08 | 1.776 | 0.366 | 1.26E-06 | 2.199 | 0.572 | 1.86E-04 |
| AAAgen | Set 7 | 136 | 1.846 | 0.227 | 3.97E-16 | 1.846 | 0.396 | 3.16E-06 | 2.697 | 0.399 | 1.37E-11 | 1.783 | 0.804 | 2.83E-02 |
| AAAgen | Set 8 | 135 | 2.444 | 0.276 | 8.98E-19 | 2.444 | 0.495 | 7.95E-07 | 2.369 | 0.454 | 1.82E-07 | 2.895 | 1.082 | 8.38E-03 |
| AAAgen | Set 9 | 134 | 4.327 | 0.296 | 1.95E-48 | 4.327 | 0.454 | 1.52E-21 | 5.138 | 0.542 | 2.57E-21 | 4.904 | 0.687 | 5.54E-11 |
| AAAgen | Set 10 | 135 | 6.343 | 0.425 | 2.58E-50 | 6.343 | 0.569 | 7.77E-29 | 6.926 | 0.774 | 3.59E-19 | 7.395 | 0.778 | 1.17E-16 |
| FinnGen | Set 1 | 131 | 1.492 | 0.307 | 1.21E-06 | 1.492 | 0.383 | 1.00E-04 | 1.493 | 0.525 | 4.43E-03 | 1.868 | 0.512 | 3.77E-04 |
| FinnGen | Set 2 | 131 | 2.103 | 0.344 | 9.61E-10 | 2.103 | 0.426 | 8.13E-07 | 1.527 | 0.639 | 1.68E-02 | 1.745 | 0.643 | 7.58E-03 |
| FinnGen | Set 3 | 133 | 2.408 | 0.272 | 7.45E-19 | 2.408 | 0.337 | 8.49E-13 | 2.537 | 0.483 | 1.51E-07 | 2.107 | 0.456 | 8.92E-06 |
| FinnGen | Set 4 | 134 | 2.157 | 0.426 | 4.24E-07 | 2.157 | 0.581 | 2.04E-04 | 2.679 | 0.833 | 1.30E-03 | 2.888 | 0.942 | 2.63E-03 |
| FinnGen | Set 5 | 132 | 2.596 | 0.446 | 5.73E-09 | 2.596 | 0.606 | 1.85E-05 | 3.050 | 0.809 | 1.62E-04 | 2.763 | 0.927 | 3.43E-03 |
| FinnGen | Set 6 | 133 | 1.671 | 0.424 | 8.22E-05 | 1.671 | 0.613 | 6.42E-03 | 0.879 | 0.750 | 2.41E-01 | 1.050 | 0.918 | 2.54E-01 |
| FinnGen | Set 7 | 127 | 2.059 | 0.606 | 6.75E-04 | 2.059 | 0.722 | 4.35E-03 | 2.808 | 0.983 | 4.30E-03 | 2.821 | 1.459 | 5.55E-02 |
| FinnGen | Set 8 | 133 | 3.155 | 0.740 | 2.03E-05 | 3.155 | 0.861 | 2.48E-04 | 3.244 | 1.159 | 5.12E-03 | 4.558 | 1.896 | 1.76E-02 |
| FinnGen | Set 9 | 133 | 4.923 | 0.748 | 4.67E-11 | 4.923 | 0.760 | 9.51E-11 | 5.472 | 1.355 | 5.36E-05 | 5.515 | 1.149 | 4.29E-06 |
| FinnGen | Set 10 | 134 | 7.500 | 1.067 | 2.06E-12 | 7.500 | 1.330 | 1.72E-08 | 9.059 | 1.748 | 2.18E-07 | 8.373 | 1.803 | 8.14E-06 |
|  |  |  |  |  | **MR-PRESSO** | | | | | | |  |  |  |
|  |  |  | **Cochran's Q P value** | **P (Intercept)** | **Beta_raw** | **SE_raw** | **P_raw** | **Beta_outlier** | **SE_outlier** | **P_outlier** | **Outliers** |  |  |  |
| AAAgen | Set 1 | 136 | 2.92E-31 | 0.407 | 0.924 | 0.199 | 7.73E-06 | 0.971 | 0.157 | 7.02E-09 | 5 |  |  |  |
| AAAgen | Set 2 | 134 | 8.00E-26 | 0.513 | 1.700 | 0.213 | 5.53E-13 | 1.844 | 0.195 | 2.58E-16 | 8 |  |  |  |
| AAAgen | Set 3 | 134 | 4.21E-17 | 0.771 | 2.051 | 0.172 | 8.31E-23 | 2.187 | 0.199 | 2.25E-20 | 3 |  |  |  |
| AAAgen | Set 4 | 136 | 5.41E-27 | 0.220 | 1.831 | 0.291 | 4.19E-09 | 1.927 | 0.227 | 4.31E-14 | 6 |  |  |  |
| AAAgen | Set 5 | 136 | 2.04E-30 | 0.933 | 1.524 | 0.332 | 1.01E-05 | 1.658 | 0.264 | 4.74E-09 | 4 |  |  |  |
| AAAgen | Set 6 | 135 | 3.80E-27 | 0.420 | 1.821 | 0.328 | 1.50E-07 | 1.803 | 0.283 | 3.24E-09 | 7 |  |  |  |
| AAAgen | Set 7 | 136 | 8.79E-30 | 0.929 | 1.846 | 0.396 | 7.50E-06 | 2.555 | 0.338 | 6.36E-12 | 3 |  |  |  |
| AAAgen | Set 8 | 135 | 8.59E-33 | 0.640 | 2.444 | 0.495 | 2.32E-06 | 2.223 | 0.357 | 6.65E-09 | 8 |  |  |  |
| AAAgen | Set 9 | 134 | 1.53E-16 | 0.265 | 4.327 | 0.454 | 9.74E-17 | 4.627 | 0.440 | 4.24E-19 | 3 |  |  |  |
| AAAgen | Set 10 | 135 | 5.00E-08 | 0.052 | 6.343 | 0.569 | 8.18E-21 | 6.570 | 0.526 | 3.61E-24 | 1 |  |  |  |
| FinnGen | Set 1 | 131 | 4.96E-05 | 0.269 | 1.492 | 0.383 | 1.59E-04 | 1.525 | 0.333 | 1.06E-05 | 2 |  |  |  |
| FinnGen | Set 2 | 131 | 8.00E-05 | 0.458 | 2.103 | 0.426 | 2.44E-06 |  |  |  | 1 |  |  |  |
| FinnGen | Set 3 | 133 | 7.22E-05 | 0.328 | 2.414 | 0.335 | 4.04E-11 |  |  |  | 1 |  |  |  |
| FinnGen | Set 4 | 134 | 7.87E-09 | 0.326 | 2.157 | 0.581 | 2.99E-04 | 2.285 | 0.549 | 5.63E-05 | 1 |  |  |  |
| FinnGen | Set 5 | 132 | 1.16E-08 | 0.811 | 2.596 | 0.606 | 3.55E-05 | 2.727 | 0.589 | 8.79E-06 | 1 |  |  |  |
| FinnGen | Set 6 | 133 | 3.75E-12 | 0.365 | 1.671 | 0.613 | 7.29E-03 | 1.902 | 0.579 | 1.32E-03 | 3 |  |  |  |
| FinnGen | Set 7 | 127 | 1.34E-03 | 0.549 | 2.061 | 0.721 | 5.01E-03 | 2.640 | 0.714 | 3.24E-04 | 1 |  |  |  |
| FinnGen | Set 8 | 133 | 4.35E-03 | 0.407 | 3.155 | 0.861 | 3.58E-04 |  |  |  | 1 |  |  |  |
| FinnGen | Set 9 | 133 | 3.79E-01 | 0.493 | 4.923 | 0.760 | 1.72E-09 |  |  |  |  |  |  |  |
| FinnGen | Set 10 | 134 | 4.33E-05 | 0.473 | 7.500 | 1.330 | 9.88E-08 | 7.605 | 1.245 | 1.08E-08 | 2 |  |  |  |

**VA Million Veteran Program**

**Core Acknowledgements for Publications**

**October 2025**

**MVP Program Office**

- Sumitra Muralidhar, Ph.D., Program Director

US Department of Veterans Affairs, 810 Vermont Avenue NW, Washington, DC 20420

- Jennifer Moser, Ph.D., Associate Director, Scientific Programs

US Department of Veterans Affairs, 810 Vermont Avenue NW, Washington, DC 20420

- Jennifer E. Deen, B.S., Associate Director, Cohort & Public Relations

US Department of Veterans Affairs, 810 Vermont Avenue NW, Washington, DC 20420

**MVP Steering Committee**

- Co-Chair: Philip S. Tsao, Ph.D.

VA Palo Alto Health Care System, 3801 Miranda Avenue, Palo Alto, CA 94304

- Co-Chair: Sumitra Muralidhar, Ph.D.

US Department of Veterans Affairs, 810 Vermont Avenue NW, Washington, DC 20420

- J. Michael Gaziano, M.D., M.P.H.

VA Boston Healthcare System, 150 S. Huntington Avenue, Boston, MA 02130

- Adriana Hung, M.D., M.P.H.,

VA Tennessee Valley Healthcare System, 1310 24th Avenue, South Nashville, TN 37212

- Dave Oslin, M.D.

Philadelphia VA Medical Center, 3900 Woodland Avenue, Philadelphia, PA 19104

- Deepak Voora, M.D.

Durham VA Medical Center, 508 Fulton Street, Durham, NC 27705

MVP Co-Principal Investigators

- J. Michael Gaziano, M.D., M.P.H.

VA Boston Healthcare System, 150 S. Huntington Avenue, Boston, MA 02130

- Philip S. Tsao, Ph.D.

VA Palo Alto Health Care System, 3801 Miranda Avenue, Palo Alto, CA 94304

**MVP Core Operations**

- Jessica V. Brewer, M.P.H., Director, MVP Cohort Operations

VA Boston Healthcare System, 150 S. Huntington Avenue, Boston, MA 02130

- Mary T. Brophy M.D., M.P.H., Director, VA Central Biorepository

VA Boston Healthcare System, 150 S. Huntington Avenue, Boston, MA 02130

- Kelly Cho, M.P.H, Ph.D., Director, MVP Phenomics

VA Boston Healthcare System, 150 S. Huntington Avenue, Boston, MA 02130

- Lori Churby, B.S., Director, MVP Regulatory Affairs

VA Palo Alto Health Care System, 3801 Miranda Avenue, Palo Alto, CA 94304

- Jacob T. Kean, Ph.D., Acting Director, VA Informatics and Computing Infrastructure (VINCI)

VA Salt Lake City Health Care System, 500 Foothill Drive, Salt Lake City, UT 84148

- Saiju Pyarajan Ph.D., Director, Data and Computational Sciences

VA Boston Healthcare System, 150 S. Huntington Avenue, Boston, MA 02130

- Robert Ringer, Pharm.D., Director, VA Albuquerque Central Biorepository

New Mexico VA Health Care System, 1501 San Pedro Drive SE, Albuquerque, NM 87108

- Luis E. Selva, Ph.D., Director, MVP Biorepository Coordination

VA Boston Healthcare System, 150 S. Huntington Avenue, Boston, MA 02130

- Shahpoor (Alex) Shayan, M.S., Director, MVP PRE Informatics

VA Boston Healthcare System, 150 S. Huntington Avenue, Boston, MA 02130

- Brady Stephens, M.S., Principal Investigator, MVP Information Center

Canandaigua VA Medical Center, 400 Fort Hill Avenue, Canandaigua, NY 14424

- Stacey B. Whitbourne, Ph.D., Director, MVP Cohort Development and Management

VA Boston Healthcare System, 150 S. Huntington Avenue, Boston, MA 02130
